# A haplotype reference panel constructed from 490,319 UK Biobank genomes improves genotype imputation for global populations

**DOI:** 10.64898/2026.06.19.26356054

**Authors:** Lars Wienbrandt, Christoph Prieß, Marcus Schmöhl, Volker Neff, Maria Gretsova, Guillermo Torres, Eike Matthias Wacker, Andre Franke, David Ellinghaus

## Abstract

We constructed a genotype imputation reference panel from whole-genome sequence data of 490,319 *UK Biobank* participants and enabled both standalone and cloud-based phasing and imputation through *EagleImp* and *EagleImp-RAP* within the UK Biobank Research Analysis Platform. The UK Biobank reference panel achieves lower phasing switch error rates than the widely used *TOPMed r3* panel (133,597 individuals) for all global superpopulations except AMR and for 25 of 26 1000 Genomes Project subpopulations. Imputation accuracy, measured by mean absolute error, improved for four of five superpopulations and 12 subpopulations for variants with minor allele frequencies down to 0.01%. Estimated squared correlation further improved across all five superpopulations and 22 subpopulations, with the largest gains in East and South Asian populations. Re-imputation of COVID-19 GWAS datasets from Italy, Germany, Spain and Norway demonstrated that common-variant imputation remains unsaturated, enhancing the discovery of genome-wide significant loci and improving fine-mapping resolution.

## 1 Main text - Letter format

Imputation services have been developed that allow researchers to easily conduct genome-wide phasing and imputation using reference panels from different genome sequencing projects (e.g., the Haplotype Reference Consortium (HRC) [1] and the Trans-Omics for Precision Medicine (TOPMed) Program [2]) without utilizing their own computational resources (**Supplementary Sections 1** and **2**, **Supplementary Figures 1–4**, and **Supplementary Table 1**). However, genome-wide sequencing (WGS) data from the *TOPMed r3* panel (133,597 genomes; the largest imputation panel so far) is only partially available via *The database of Genotypes and Phenotypes (dbGaP)*, and the panel was assembled from more than 80 individual studies (so-called *TOPMed* projects), which prevents its use on standalone servers or cloud-based research analysis platforms such as the UK Biobank Research Analysis Platform (*UKB-RAP*).

In this study, we constructed whole-genome haplotype reference panels from 490,319 UK Biobank genomes comprising more than 361 million genetic variants — to our knowledge the largest imputation panels generated to date (**Table 1**, **Online Methods**, **Supplementary Section 3**, **Supplementary Table 2**). To enable reference-based phasing and imputation with these panels, we extended the *EagleImp* software framework [3] with new process options and with cloud-execution capabilities for running *EagleImp* within the *UKB-RAP* (**Table 2**). These new functionalities operate both on the project’s secure standalone compute server — used here for all UKB-based benchmark analyses — and in an implementation of *EagleImp* inside the *UKB-RAP* (*EagleImp-RAP*), enabling reproducibility directly within the *UKB-RAP* (**Online Methods**). Our goal was to provide the UK Biobank research community with a highly accurate imputation solution for diverse global populations, while enabling the use of the enhanced phasing and imputation features implemented in *EagleImp*.

**Table 1.** Summary of the *UK Biobank* imputation reference panels constructed in this study. Reference panels were generated from *UK Biobank* Graphtyper genotype-level variant data for chromosomes 1-22 and X (490,319 individuals, see **Online Methods**). Four panel versions were created, differing only in their reference panel minor allele frequency (rpMAF) thresholds and genome builds (hg19 and hg38). The two hg38 panels formed the basis of all phasing and imputation analyses presented in this study.

| Name | Description | #Samples | #Variants |
| --- | --- | --- | --- |
| UKB complete | Complete *UK Biobank* reference panel, hg38 | 490,319 | 361,441,468 |
|  | Complete *UK Biobank* reference panel, hg19 | 490,319 | 359,383,785 |
| UKB MAF0.0001 | *UK Biobank* reference panel filtered for reference panel MAF (rpMAF) $\geq 0.01\%$, hg38 | 490,319 | 51,939,464 |
| | *UK Biobank* reference panel filtered for reference panel MAF (rpMAF) $\geq 0.01\%$, hg19 | 490,319 | 51,632,019 |

**Table 2.**
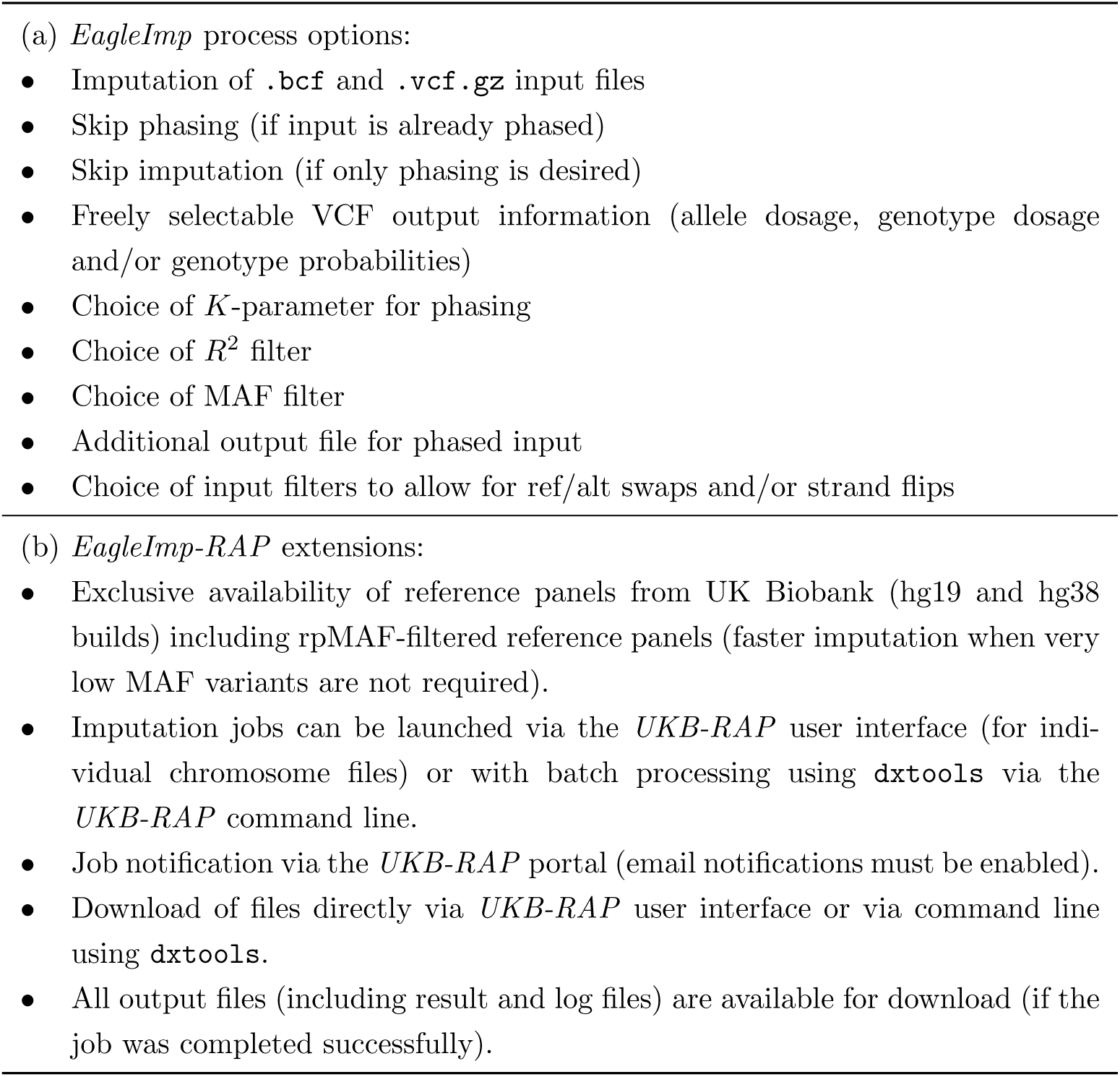
New functionalities added to the *EagleImp* software framework to enable efficient reference-based phasing and imputation with the UK Biobank reference panels. Section (a) lists the *EagleImp* process options. Section (b) summarizes the additional execution capabilities developed for running *EagleImp* within the UK Biobank Research Analysis Platform (*EagleImp-RAP*).

For our first benchmark of imputation quality, we extracted commonly used input variants for imputation available on the *Illumina Global Screening Array (GSA)* [4] from the *1000 Genomes* reference panel data and lifted them to genome build GRCh38 using the *UCSC liftover* tool [5]. We performed imputations separately for the five superpopulation datasets (AFR, AMR, EAS, EUR and SAS) and for the 26 subpopulation datasets (**Supplementary Table 3**), firstly with *EagleImp* and our complete UKB reference panel, secondly with *EagleImp* and our rpMAF-filtered UKB reference panel, and thirdly with the *TOPMed Imputation Server* which uses the *TOPMed r3* reference panel. From each imputation run, we calculated mean imputation estimated *R*^2^ values from reported per-variant *R*^2^ values for three rpMAF categories starting from 0.01%. rpMAF categories are based on the UKB reference panel since per-variant rpMAFs from the *TOPMed r3* panel are not publicly available. For each rpMAF category, we calculated the percentage change in imputation quality of *EagleImp* relative to the *TOPMed Imputation Server* based on variants shared between the panels (**Supplementary Figure 5**). Please refer to **Supplementary Section 4** for details on the conducted benchmarks.

Comparing the first two *EagleImp* imputation results showed that using the rpMAF-filtered UKB reference panel does not lead to a significant reduction in estimated imputation *R*^2^ (**Supplementary Figure 6** and **Supplementary Table 4**). Thus, users who only wish to impute variants with rpMAF *≥* 0.01% (which typically achieve high estimated *R*^2^) may use the rpMAF-filtered UKB panel for faster imputation without noticeable loss of accuracy.

Comparing the estimated *R*^2^ distributions from *EagleImp* (complete UKB panel) with those from the *TOPMed Imputation Server* revealed higher imputation quality for all five superpopulations across the entire allele frequency spectrum, except for a decrease for AMR (-0.76%) for rpMAF *≥* 1%. The largest gain was observed for East Asian samples (EAS; +24.39% for rpMAF *≥* 0.01%). For rpMAF *≥* 0.01%, imputation quality improved in 22 of the 26 subpopulations (max. +15.25% for JPT; min. +0.10% for ASW) and decreased in four (worst -3.33% for GWD). For rpMAF categories *≥* 0.1% and *≥* 1%, 25 and 24 out of 26 populations, respectively, showed improved imputation quality. See **Figure 1**, **Supplementary Figure 7** and **Supplementary Table 5** for details.

**Fig. 1.**
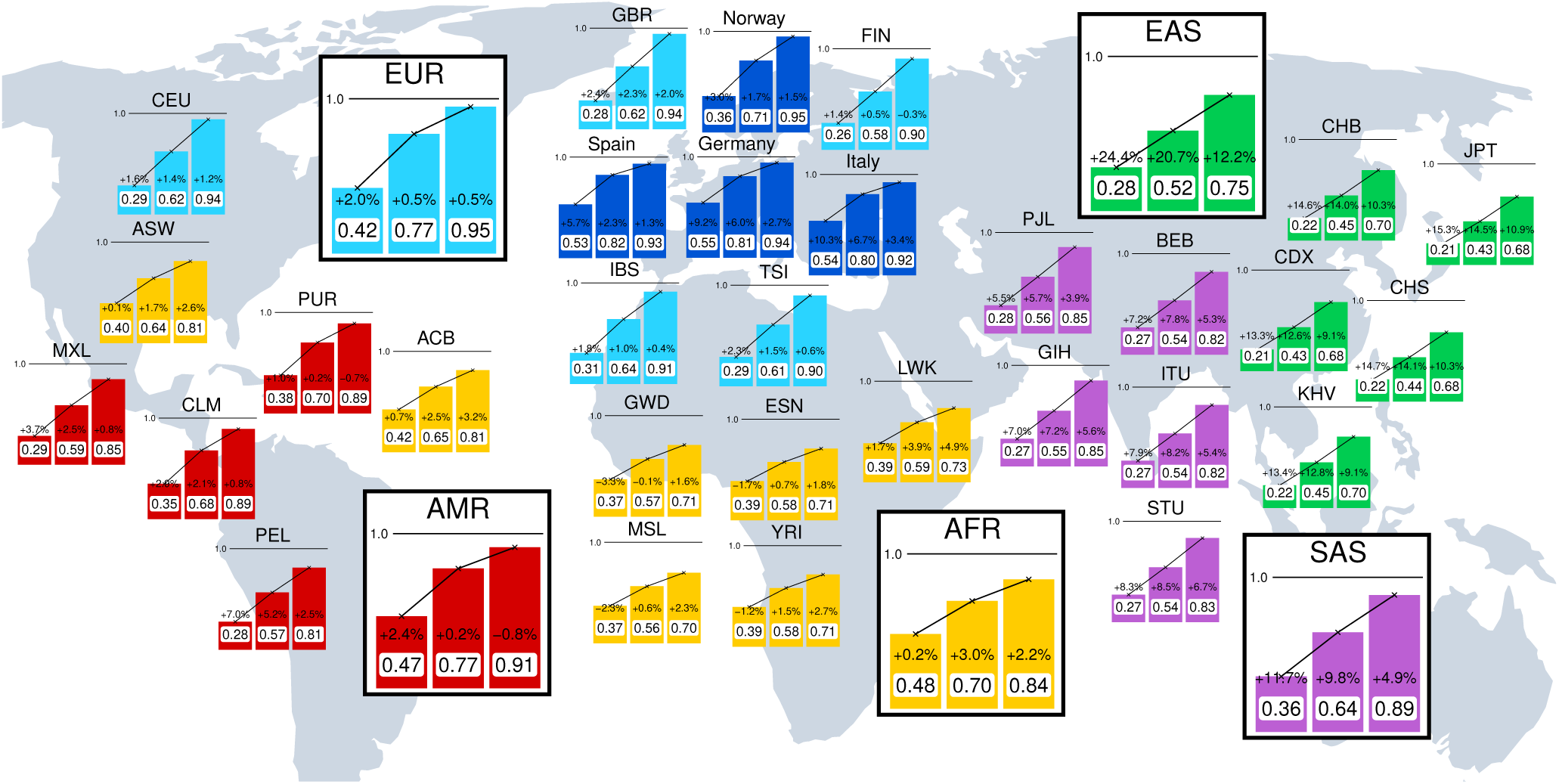
World map illustrating the mean estimated imputation *R*^2^ reported from *EagleImp* and the improvement of *EagleImp* with the complete UK Biobank reference panel (UKB complete) compared to the mean estimated *R*^2^ reported from the *TOPMed Imputation Server* for 35 benchmark datasets from around the world (see **Supplementary Table 3**). Each (super)population shows three bars, where the first (left) bar indicates the mean *R*^2^ for variants with a reference panel MAF (rpMAF) *≥* 0.01%, the second (middle) bar indicates the mean *R*^2^ for rpMAF *≥* 0.1%, and the third (right) bar indicates the mean *R*^2^ for rpMAF *≥* 1% computed across all imputed variants. Percentages indicate the improvement of *EagleImp* in reported estimated *R*^2^ in relation to the *TOPMed Imputation Server* for imputed variants shared between the UK Biobank reference panel and the *TOPMed* reference panel (**Supplementary Figure 5**). Genome-wide SNP data from (super)populations are taken from the 1000 Genomes Project reference data reduced to Illumina GSA-chip variants. The datasets Germany, Italy, Norway and Spain are GSA-typed GWAS case-control datasets (see main text and **Supplementary Section 4** for details).

Estimated *R*^2^ is the key post-imputation quality metric because it is available for each variant from the reference panel and is routinely used to exclude poorly imputed variants in GWAS analyses. The average estimated *R*^2^ for a given MAF category provides the user with an indication of how many variants are expected to pass a chosen minimum *R*^2^ filter, since estimated *R*^2^ serves as a per-variant quality criterion (see *EagleImp* features in **Table 2**). When ground-truth haplotypes with accurate phase information are available (the 1000 Genomes Project haplotypes incorporate trio-based phasing information and are available across diverse populations), direct measures such as switch error rate (SER), mean absolute error (MAE), and correlation *r*^2^ can additionally quantify phasing and imputation performance (see **Supplementary Sections 4.2.2-4.2.4**).

For each imputation run of the five superpopulations and the 26 subpopulations, we calculated SER to assess pre-imputation phasing quality as well as MAE and correlation *r*^2^ for imputed genotypes across four rpMAF range categories. The complete UKB reference panel showed lower SERs than the *TOPMed r3* panel for all superpopulations except AMR. Across the 26 subpopulations, 25 showed lower SERs with the UKB panel, with one subpopulation (PUR) showing no difference in the SER, while the *TOPMed r3* panel yielded lower SERs in two cases (MXL, PUR). See **Supplementary Figures 8-11** and **Supplementary Table 6** for details.

MAE-based analyses showed that the complete UKB reference panel outperformed the *TOPMed r3* panel for all rpMAF range categories in the SAS and EAS superpopulations, for rpMAF range [0.01%, 0.1%) and [0.1%, 1%) in EUR, and for rpMAF range [0.01%, 0.1%) in AMR (with similar performance for [0.1%, 1%)); the *TOPMed r3* panel performed better across all rpMAF range categories in AFR. The rpMAF-filtered UKB reference panel showed the same overall result with slightly improved performance, including lower MAE than the *TOPMed r3* panel for AMR in [0.1%, 1%). Subpopulation-level MAE results varied by reference panel, population, and rpMAF range. Across the 26 subpopulations, the rpMAF-filtered UKB reference panel showed lower MAE across all rpMAF range categories for 12 populations, the *TOPMed r3* panel for six populations, and mixed results across eight populations depending on rpMAF range category. See **Figure 2**, **Supplementary Figures 12-14** and **Supplementary Tables 7 and 8** for details.

**Fig. 2.**
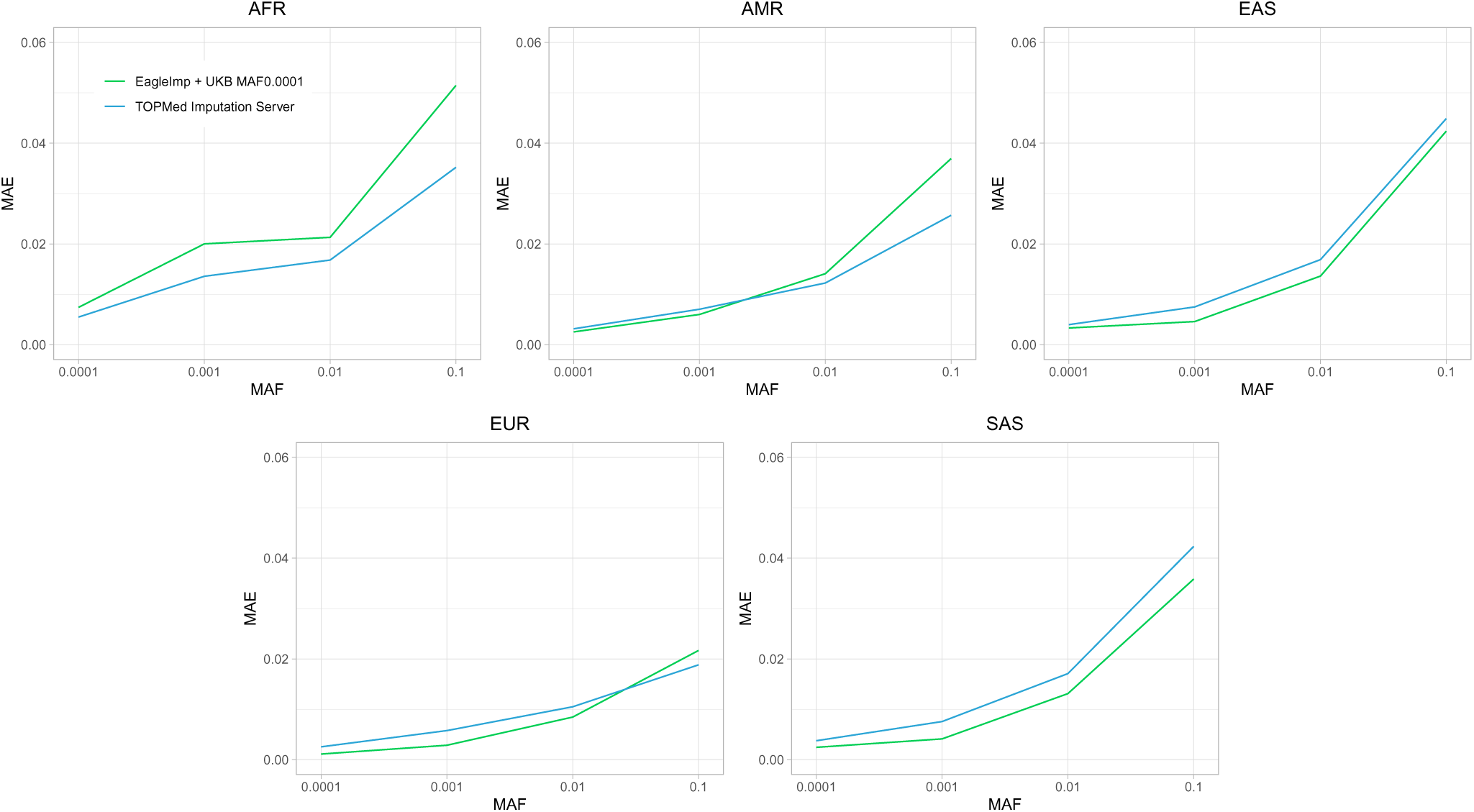
Mean absolute error (MAE) imputation quality results for the five superpopulations from the 1000 Genomes Project based on the 1000 Genomes reference panel ground truth, stratified by four rpMAF range categories [0.01%, 0.1%), [0.1%, 1%), [1%, 10%), [10%, 50%] and imputed by *EagleImp* with the rpMAF-filtered UK Biobank reference panel (UKB MAF0.0001) (green lines) and the *TOPMed Imputation Server* (*TOPMed r3* panel) (blue lines). MAE calculation is based on 30,098,506 variants shared between the UKB MAF0.0001, the *TOPMed r3* panel and the 1000 Genomes Project panel.

A commonly used alternative imputation metric is the squared Pearson correlation coefficient *r*^2^ between imputed allele dosages and masked ground-truth genotypes. However, due to its instability in small cohorts and its strong dependence on the estimation method (**Supplementary Section 5**, **Supplementary Figures 15-22**, **Supplementary Tables 9-12**), we do not interpret small numerical differences in correlation *r*^2^ as meaningful indicators of relative panel performance.

We repeated the benchmark using four real-world GWAS case-control datasets from Germany (3,351 samples), Italy (2,090 samples), Norway (324 samples) and Spain (1,725 samples) from [6] and [7]. As before, using the rpMAF-filtered UKB reference panel did not reduce the estimated imputation *R*^2^ (**Supplementary Figure 23**), and the complete UKB panel improved imputation quality over the *TOPMed r3* panel across the entire allele frequency spectrum (**Supplementary Figure 24**). **Figure 1** summarizes estimated *R*^2^ values and percentage improvements for the five superpopulations, the 26 subpopulations, and the four real-world GWAS case-control datasets across three rpMAF thresholds.

Next, we examined the impact of UKB-based imputation on association testing and the detection of genome-wide significant association loci (*p <* 5*×*10*^−^*^8^). Since the real-world Italy GWAS dataset showed the strongest improvement in imputation quality, we repeated the GWAS study from [6] (835 cases of severe COVID-19 with respiratory failure and 1,255 healthy controls) (**Online Methods**) using imputed genotypes from *EagleImp* (rpMAF-filtered UKB reference panel) and from the *TOPMed Imputation Server*. Genome-wide association results were largely similar (see Manhattan and quantile-quantile (Q-Q) plots in **Supplementary Figures 25** and **26**), except at the well established and most strongly associated COVID-19 susceptibility locus at 3p21.31 (also visible in the tail of the distribution of p-values in the Q-Q plots). First, compared to *TOPMed Imputation Server* imputation (which was also performed in the original study by [6]), only the UKB-based imputation yielded a genome-wide significant association at 3p21.31, demonstrating improved common-variant imputation in this European cohort. Second, the new lead SNP *rs73064425* replicated the signals reported in three independent GWAS on COVID-19 from [8], [9] and [10]. Linkage disequilibrium (LD) and Bayesian fine-mapping analysis further confirmed improved resolution in imputation quality by *EagleImp* compared to the *TOPMed Imputation Server* (**Figure 3**, **Supplementary Tables 13 and 14)**.

**Fig. 3.**
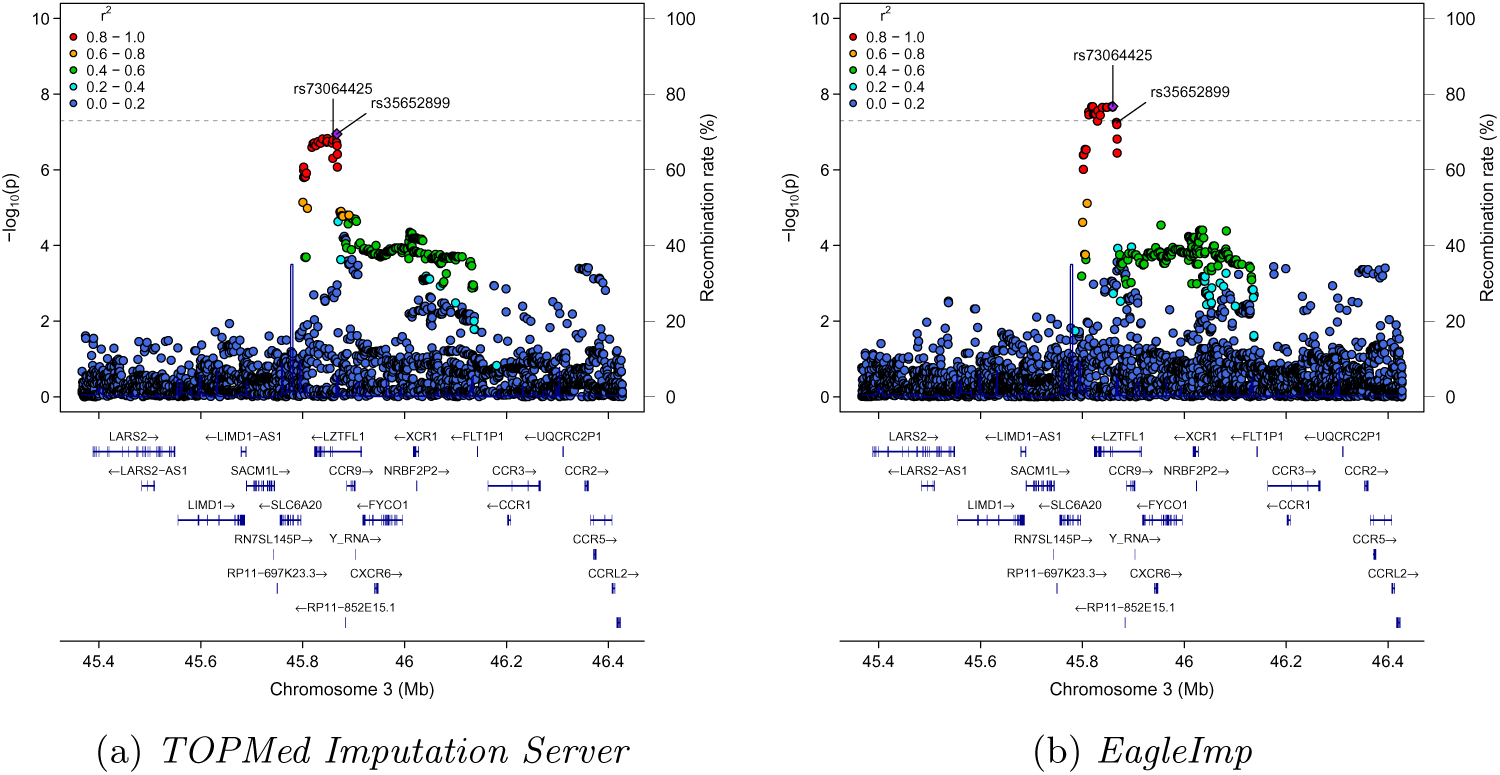
Regional association plots illustrating the improvement of association testing through improved imputation quality for common variants (MAF *≥* 1%) at the most strongly associated COVID-19 risk locus (3p21.31) using the COVID-19 GWAS dataset *Italy* (original data from [6]), indicating that imputation of common variants with the *TOPMed Imputation Server* is still not saturated in European populations. **(a)** Imputation with the *TOPMed Imputation Server* (*TOPMed* reference panel) and **(b)** imputation with *EagleImp* (rpMAF-filtered *UKB* reference panel). Imputed variants were filtered for reported estimated *R*^2^ *≥* 0.6 and MAF *≥* 1% for direct comparison, as performed in [6]. The dotted line represents the genome-wide association significance threshold at *p* = 5 *×* 10*^−^*^8^, with the Italian GWAS dataset showing a genome-wide significant association result for the first time when performing *EagleImp*’s UKB reference panel imputation. Highlighted variants are *rs73064425* (lead SNP of the *EagleImp* imputation) and *rs35652899* (lead SNP of the *TOPMed Imputation Server* imputation). The association signal improvement correlates with the observed improvement in estimated *R*^2^ of the *Italy* dataset shown in Figure 1. The new lead SNP *rs73064425* from the *EagleImp* imputation is also the lead SNP identified in the larger *GenOMICC (Genetics Of Mortality In Critical Care)* GWAS in 2,244 critically ill patients with COVID-19 from 208 UK intensive care units [8], in the *GenOMICC* follow-up whole-genome sequencing study in 7,491 critically ill individuals with COVID-19 versus 48,400 controls [9] and in the GWAS of 2,184 severely ill cases with COVID-19 versus 689,620 COVID-19-negative or unknown population controls [10]. Linkage disequilibrium (LD) analysis showed that the established lead variant *rs73064425* is only in moderate LD with *rs35652899* (LD *r*^2^ = 0.6625; lead SNP from *TOPMed Imputation Server* imputation), further indicating an improvement in imputation quality with *EagleImp* over the *TOPMed Imputation Server*. A Bayesian fine-mapping analysis using the *SuSiE* software [18] (**Online Methods**) for the Italian GWAS results from the *EagleImp* imputation identified *rs73064425* as the most likely causal variant in a credible set that contain the causal variant with a probability of *≥* 95% for locus 3p21.31 for critically ill patients with COVID-19 (**Supplementary Table 13**). On the other hand, the Bayesian fine-mapping analysis for the results of the *TOPMed Imputation Server* listed the established lead variant *rs73064425* on the 4th rank of most likely causal variants, with a larger (less precise) credible set of associated variants (**Supplementary Table 14**).

Since very large reference panels such as the UKB reference panels require additional compute resources, an increase in runtime and memory requirements for phasing and imputation is expected. We therefore measured and compared phasing and imputation runtimes (without queueing times) for the complete and rpMAF-filtered UKB reference panels (both using *EagleImp* and *EagleImp-RAP*), and the *TOPMed r3* panel (*TOPMed Imputation Server*) (**Supplementary 4.3**). The runtime comparison of *EagleImp* and *EagleImp-RAP* did not show significant differences. On average across all 35 benchmark populations (see **Figure 1**), the runtime of the *TOPMed Imputation Server* with the *TOPMed r3* panel was about twice as long as that of *EagleImp* and *EagleImp-RAP* with the complete UKB reference panel, although the UKB reference panel contains more than 3.6 times as many samples as the *TOPMed r3* panel. For the rpMAF-filtered UKB reference panel, we measured an average 4 to 13 times shorter runtime of *EagleImp* and *EagleImp-RAP* compared to the *TOPMed Imputation Server*. See **Table 3** for all measured runtimes.

**Table 3.** Imputation runtimes for phasing and imputation using the UK Biobank reference panels. Runtimes were measured for the complete (UKB complete, hg38) and MAF-filtered (UKB MAF0.0001, hg38) panels using *EagleImp* on the project’s secure standalone compute server and using *EagleImp-RAP* within the UK Biobank Research Analysis Platform (UKB-RAP). For comparison, runtimes of the *TOPMed Imputation Server* (*TOPMed r3* panel) are included. Details of the runtime measurement procedures for all systems are provided in **Supplementary 4.3**. Runtimes exclude queueing times.

| Dataset | *EagleImp* standalone |  | *EagleImp-RAP* |  | *TOPMed* |
| --- | --- | --- | --- | --- | --- |
|  | UKB | UKB | UKB | UKB | *TOPMed r3* |
|  | complete | MAF0.0001 | complete | MAF0.0001 |  |
|  | hh:mm:ss | hh:mm:ss | hh:mm:ss | hh:mm:ss | hh:mm:ss |
| *Datasets from 1000 Genomes Project [19] superpopulations:* |  |  |  |  |  |
| AFR | 01:09:03 | 00:29:49 | 01:27:00 | 00:38:00 | 02:34:55 |
| AMR | 01:01:16 | 00:26:26 | 01:14:00 | 00:34:00 | 02:03:02 |
| EAS | 01:02:31 | 00:26:48 | 01:20:00 | 00:37:00 | 02:25:52 |
| EUR | 01:17:33 | 00:29:53 | 01:25:00 | 00:35:00 | 02:01:16 |
| SAS | 01:06:31 | 00:27:37 | 01:18:00 | 00:34:00 | 02:09:50 |
| *Datasets from 1000 Genomes Project [19] subpopulations:* |  |  |  |  |  |
| ACB | 00:42:16 | 00:21:50 | 00:53:00 | 00:29:00 | 02:26:56 |
| ASW | 00:41:19 | 00:21:19 | 00:52:00 | 00:28:00 | 02:22:39 |
| BEB | 00:44:01 | 00:20:41 | 00:54:00 | 00:29:00 | 01:35:40 |
| CDX | 00:41:20 | 00:23:00 | 00:54:00 | 00:28:00 | 01:21:38 |
| CEU | 00:42:38 | 00:22:41 | 00:54:00 | 00:27:00 | 01:14:54 |
| CHB | 00:43:30 | 00:22:45 | 00:55:00 | 00:28:00 | 01:20:00 |
| CHS | 00:41:51 | 00:23:14 | 00:55:00 | 00:27:00 | 05:17:13 |
| CLM | 00:44:42 | 00:22:07 | 00:56:00 | 00:28:00 | 01:23:36 |
| ESN | 00:43:24 | 00:21:23 | 00:54:00 | 00:28:00 | 01:08:49 |
| FIN | 00:46:08 | 00:22:39 | 00:55:00 | 00:28:00 | 01:26:36 |
| GBR | 00:45:20 | 00:22:52 | 00:54:00 | 00:28:00 | 01:26:53 |
| GIH | 00:42:20 | 00:24:14 | 00:55:00 | 00:27:00 | 01:28:13 |
| GWD | 00:41:46 | 00:21:44 | 00:55:00 | 00:28:00 | 01:28:10 |
| IBS | 00:46:59 | 00:24:24 | 00:57:00 | 00:29:00 | 01:26:44 |
| ITU | 00:43:19 | 00:22:13 | 00:54:00 | 00:28:00 | 01:14:54 |
| JPT | 00:45:44 | 00:21:23 | 00:55:00 | 00:29:00 | 01:17:17 |
| KHV | 00:43:11 | 00:21:13 | 00:55:00 | 00:28:00 | 01:12:48 |
| LWK | 00:41:04 | 00:21:55 | 00:54:00 | 00:28:00 | 01:16:24 |
| MSL | 00:42:56 | 00:21:53 | 00:52:00 | 00:27:00 | 01:13:54 |
| MXL | 00:42:02 | 00:21:58 | 00:53:00 | 00:27:00 | 01:12:15 |
| PEL | 00:42:09 | 00:22:19 | 00:53:00 | 00:27:00 | 01:07:59 |
| PJL | 00:44:51 | 00:22:19 | 00:55:00 | 00:27:00 | 01:27:30 |
| PUR | 00:46:18 | 00:21:55 | 00:57:00 | 00:28:00 | 01:04:24 |
| STU | 00:42:05 | 00:22:48 | 00:53:00 | 00:27:00 | 01:18:30 |
| TSI | 00:47:27 | 00:24:39 | 00:59:00 | 00:28:00 | 01:12:43 |
| YRI | 00:43:13 | 00:21:40 | 00:53:00 | 00:27:00 | 01:10:13 |
| *Datasets from COVID-19 studies [6, 7]:* |  |  |  |  |  |
| Germany | 06:56:03 | 01:35:13 | 05:23:00 | 01:21:00 | 13:12:32 |
| Italy | 04:52:13 | 01:08:34 | 03:36:00 | 01:08:00 | 08:19:25 |
| Norway | 01:14:25 | 00:25:04 | 01:17:00 | 00:31:00 | 05:34:34 |
| Spain | 04:08:37 | 00:58:01 | 03:07:00 | 00:57:00 | 06:26:50 |

Interestingly, Shi et al. recently published a haplotype reference panel of 78,195 individuals from the *GenomicsEngland (GEL)* study used for imputation of GWAS data from UK Biobank [11]. We were unable to include the GEL imputation reference panel in our benchmarks, as the GEL imputation reference panel is not publicly available and there is no publicly accessible GEL imputation server for direct use of the panel. Shi et al. observed an improvement in phasing and imputation quality over the *TOPMed Imputation Server* with the *TOPMed r2* panel for the Great Britain (GBR) subpopulation and the South Asian (SAS) population from the 1000 Genomes Project, but a decrease in phasing and imputation quality for the African (AFR), American (AMR), East Asian (EAS) and non-Finnish European (NFE) populations for variants with MAF *≥* 0.01%. Since our UKB reference panel imputation benchmarks with 35 study populations showed an overall improvement of imputation quality up to 24.39%, we believe that the UKB reference panel is the most suitable reference panel for imputing worldwide populations if no population-specific reference panel is available, as in the example of the GEL reference panel for the majority of the British population. We would like to point out that the *TOPMed r3* panel with a total of 244,838,777 reference panel-specific variants has more reference panel-specific variants (with unknown allele frequencies) than the complete UKB reference panel with 161,298,331 panel-specific variants (with a minor allele count of at least 4), which makes it possible to impute more study population-specific variants from non-European populations with the *TOPMed r3* panel.

Through our re-imputation benchmarks of published COVID-19 GWAS case-control datasets and subsequent association and fine-mapping analyses, we were able to show that imputation of common variants (MAF *≥* 1%) is not saturated in respect to imputation quality for European study populations, in contrast to the results of the imputation benchmarks using the GEL reference panel [11]. For European populations that are not well represented in current imputation reference panels, we therefore recommend imputation with the UKB reference panel.

A limitation of our study is that for many populations worldwide there is still room for improvement in terms of imputation quality, especially for rare variants. The imputation quality benchmark measures MAE and correlation *r*^2^ based on a known ground truth have advantages and disadvantages and may be inaccurate for low MAFs. And the average estimated imputation *R*^2^ for variants with a MAF *≥* 0.01% is still low for most worldwide subpopulations. A global superpanel with millions of haplotypes or the availability of several population-specific reference panels worldwide via imputation servers or cloud-based platforms could help here in the future.

## Online Methods

### Phasing of WGS data from UK Biobank

This section summarizes the process of creating the complete UK Biobank reference panel. The populations represented in the UKB reference panels, as reported in [12], are summarized in **Supplementary Table 2**. Command calls and technical details can be found in **Supplementary Section 3**.

#### Data preparation of WGS data from UK Biobank

For the UKB reference panel we used the GraphTyper population level whole-genome sequencing (WGS) variants (500k release) in pVCF format. The data is available in the *UK Biobank Research Access Platform* (*UKB-RAP*) (https://ukbiobank.dnanexus.com) as packed `.vcf.gz` files in unphased chunks of 20 kbp. The total number of files is 151,561 with an average file size of 9.96 GB which sums up to a total of 1509.29 TB. We created a custom tool *vcffilter* written in *C* to extract variant positions, alleles, the `FILTER` and `INFO` columns, and the genotypes. We used the *UKB-RAP* to process all pVCF files with this tool, resulting in the same number of files but with an average file size of only 27.86 MB and a total size of 4123.70 GB. We then downloaded the generated files from the *UKB-RAP* to our own computing resources for further processing. For restoring a valid VCF from our custom download format, we used a simple pendant of our extraction tool written in C++. Both tools are available in the *Vcffilter* tool set at https://github.com/ikmb/vcffilter. Furthermore, the restoration tool performs custom filtering and conversion of multi-allelic variants to several bi-allelics on-the-fly, which is faster than first restoring a complete VCF-file with subsequent application of *bcftools* [13] for filtering and conversion. In particular, we converted all multi-allelic variants to bi-allelics as if *bcftools* was applied with `bcftools norm -m`, and, as suggested by Delaneau [14] for quality control (QC), we filtered out all variants with (a) a value other than `PASS` in the `FILTER` column, (b) a missingness rate of greater than 10%, and (c) an alternative allele score (*AAScore*) of less than 0.8. Furthermore, we created two batches of files by (i) keeping only variants with a minor allele frequency (MAF) of greater than or equal to 0.001, and (ii) keeping variants with a minor allele count (MAC) of greater than or equal to 4.

#### Phasing of WGS data from UK Biobank

The phasing process using *SHAPEIT5* [15] is divided into two parts. Firstly, a scaffold is created by phasing only common variants with MAF *≥* 0.001, i.e. our first batch, with the tool *phase common*. Secondly, the tool *phase rare* is used to phase the rare variants from our second batch, but using the phased scaffold from the first step.

For phasing the common variants, chunks were created of size around 20-30 Mbp for each chromosome, including an overlap of around 2 Mbp, by simply concatenating the corresponding 20 kbp-chunk files using *bcftools* with the --naive-force option. We used the chunk limits in correspondence with the chunks used by Delaneau in https://github.com/odelaneau/shapeit5/tree/main/tasks/phasingUKB_200k_release/autosomes/chunks.tgz. Phasing is then conducted by calling *phase common* with the default options. As the chunks were phased separately and the phases could switch between chunks, a subsequent *ligation* step is necessary to create the final scaffold. We used the ligation option -l provided by *bcftools* to ligate and merge in one step.

For phasing the rare variants, we used the second batch of our input files with variants with MAC *≥* 4. Now, we created batches with a size of approximately 5 Mbp and without overlaps. We used *phase rare* from *SHAPEIT5* with declared input-region and scaffold to conduct the phasing.

#### Creating Qref reference panel files

We used *bcftools* with the --naive option to concatenate the phased chunks from the previous step. Ligation, as for generating the scaffold, is not required here as the phases are already in concordance to the previously generated scaffold used for phasing the rare variants. Next, we converted the generated phased chromosome files to our *Qref* format used in *EagleImp* [3] for a much faster reference panel loading during imputation (compared to loading a reference panel from .bcf or .vcf.gz format).

#### Special treatment of chromosome X

Male samples are encoded diploid on chromosome X in the UKB Graphtyper data. However, diploid calls are allowed in the *PAR1* and *PAR2* regions (PAR1 and PAR2 of the X and Y chromosome pair recombine during meiosis), but have to be haploid in the *nonPAR* region (chrX:2781480-155701382). To take account for this, we handled these three regions separately. In particular, we firstly phased the *PAR1/2* regions and also the *nonPAR* region separately acording to the procedure described above. This was possible because (haploid) males were encoded as homozygous diploid in the *nonPAR* region. We fixed the ploidy afterwards for the final phased output using the standard *bcftools* plugin *fixploidy* for each chunk. As this step potentially affects allele counts, we applied the *bcftools* plugin *fill-tags* to correct the allele count (`INFO/AC`) and allele number (`INFO/AN`) fields, and finally filtered again for MAC *≥* 4 using *bcftools filter* to stay consistent with our original demand on the panel. Finally, three separate reference panel files in *Qref* format for chromosome X (one for each *PAR1/nonPAR/PAR2* region) were generated as described above.

### The rpMAF-filtered reference panel UKB MAF0.0001

In general, imputation is often conducted as a preliminary step in a GWAS analysis. In order to reduce noise in the association output, usually prefiltering for high-quality imputed variants is done. This is achieved by filtering using an estimated *R*^2^ threshold of, for example, 0.6 or higher. Our benchmarks showed that regardless of the input dataset, the mean estimated *R*^2^ of variants with an rpMAF *<* 0.0001 is far below 0.6, thus it is unlikely to generate *R*^2^-values higher than 0.6 for such variants and their contribution to the GWAS analysis is rather small. Consequently, the use of the complete UK Biobank reference panel in GWAS is only required if an analysis of rare variants is desired and a sufficient GWAS sample size is available (for sufficient statistical power). For a typical GWAS analysis of common variants we therefore decided to provide a second reference panel with variants with MAF *≥* 0.0001 for a much faster imputation. We thus used our previously generated phased chromosome files and filtered for MAF *≥* 0.0001 and converted the output to the *Qref* format again. This second panel can be selected in *EagleImp* to conduct faster imputation for common GWAS analyses without significant loss of imputation quality (see **Results**). We have proven the persistent quality with the runtime advantages in our benchmarks. An exemplary GWAS conducted with the rpMAF-filtered UKB reference panel is presented in the **Results** section and described below.

### Association testing of re-imputed GWAS data

To demonstrate the capabilities of the UKB panel, we repeated a GWAS of severe COVID-19 with respiratory failure with original data from [6] for 835 Italian cases with severe respiratory failure and 1,255 Italian controls. We re-imputed the *Italy* dataset with *EagleImp* using the rpMAF-filtered UKB panel (UKB MAF0.0001) with phasing default parameter *K* = 10, 000. We used the *BIGwas* [16] association software pipeline with *PLINK2* [17] and 10 principal components from principal component analysis (PCA) to generate single-marker summary statistics from imputed data. The summary statistics were further filtered with an estimation *R*^2^-value of 0.6 and a MAF of 0.01 (1%) for comparability with the results of the original study.

### Bayesian fine-mapping analysis of re-imputed GWAS data

Statistical fine-mapping analysis was performed with the software *SuSiE* [18] at locus 3p21.31 for GWAS association results from the re-imputed Italian GWAS dataset. *SuSiE* using the `susie rss` method calculated posterior inclusion probabilities (PIP) for each SNP at locus 3p21.31 to determine the 95% credible set of SNPs assuming one credible set of variants, thus generating the minimum set of variants containing the causal variant with *≥* 95% certainty.

### Implementation of *EagleImp-RAP*

*EagleImp-RAP* was implemented as a DNAnexus applet on the *UKB-RAP*, following the DNAnexus developer guidelines at https://documentation.dnanexus.com/developer/apps/intro-to-building-apps. Using the *dx-app-wizard* from the *dxtools* package, we generated a standardized *UKB-RAP* applet structure containing the execution *bash* script, a `.json` configuration file and a template directory structure for required binaries and libraries. *EagleImp*, *htslib* and *bcftools* were compiled for the *UKB-RAP* target environment. We also added a pre-compiled version of Intel’s *libtbb* (as required by *EagleImp*). The execution script handles argument parsing, retrieval and indexing of input files, and downloading the selected UKB reference panel from a predefined location within *UKB-RAP*. After running *EagleImp* on the acquired virtual machine (VM), the script collects and returns all output files to the user’s project space. Chromosome X input files require special handling, as male samples are haploid in nonPAR regions and diploid in PAR1/2. The applet therefore splits chromosome X into PAR1, nonPAR and PAR2 segments, processes each region separately using the appropriate reference panel, and merges the results into a unified output file.

## Supplementary information

Supplementary information is attached as a separate PDF document.

## Author contributions

L.W. processed the UK Biobank data to create the imputation reference panel, implemented all server backend functionalities including the software *EagleImp* and *EagleImp-RAP*, conducted and analyzed the benchmarks. C.P. and M.S. implemented the benchmark web service frontend functionality. V.N. and C.P. installed and configured the job queuing system *SLURM* for benchmarks. M.G. created the regional association plots and conducted the Bayesian fine-mapping analysis. L.W. and E.M.W. designed the data processing pipeline. G.T., A.F. and D.E. provided the UK Biobank data and enabled the use of the *UKB-RAP* services. D.E. is project leader and conceived, designed and managed the study and benchmark experiments. L.W. and D.E. wrote the manuscript. All authors reviewed and approved the final manuscript.

## Data availability

UKB-based imputation analyses were performed exclusively within secure compute environments approved for UK Biobank Project 139525. The complete UKB reference panel (UKB complete) and the MAF-filtered version (UKB MAF0.0001) were processed and benchmarked on the project’s institutional secure compute server, which corresponds to the standalone backend server of *EagleImp-Web* but is not accessible to external users. To demonstrate reproducibility within UK Biobank’s cloud infrastructure, we additionally implemented *EagleImp-RAP*, a version of *EagleImp* within the UK Biobank Research Analysis Platform (*UKB-RAP*), enabling benchmark runs directly within *UKB-RAP*. The benchmark datasets created from 1000 Genomes data are available for download at https://hybridcomputing.ikmb.uni-kiel.de/imputation-benchmark. These include all super population and subpopulation datasets from the *1000 Genomes Project* [19], which have been reduced to Illumina GSA-chip variants used for our benchmarks.

## Code availability

The *EagleImp* software used for standalone phasing and imputation is freely available under the GPL v3 license at https://github.com/ikmb/eagleimp. The source code for creating the *EagleImp-RAP* applet (under the GPL v3 license) including a usage example is available at https://github.com/ikmb/eagleimp-rap. Our custom C/C++ tools for fast data extraction from VCF files and VCF restoration are available for download in the *Vcffilter* project under the GPL v3 license at https://github.com/ikmb/vcffilter.

## Ethical approval

GWAS data from the COVID-19 GWAS Group: All study participants provided written informed consent, and the study was approved by the ethics boards of the participating institutions in agreement with the Declaration of Helsinki principles.

## Acknowledgments

This research has been conducted using the UK Biobank Resource under application number 139525. The authors would like to thank the COVID-19 GWAS Group for the use of the COVID-19 GWAS data for the benchmark. This project was funded by grants from the *Deutsche Forschungsgemeinschaft (DFG)* for Lars Wienbrandt and David Ellinghaus (grant no. WI 4908/1-1 and EL 831/3-1). The study received infrastructure support from the DFG Cluster of Excellence “Precision Medicine in Chronic Inflammation (PMI)” (DFG Grant: DFG EXC 2167/2) and was funded by the Deutsche Forschungsgemeinschaft (DFG, German Research Foundation) research unit ”miTarget” (project number 426660215; INF (EL 831/5-1; EL 831/5-2)).

