## Supplementary material for "A haplotype reference panel constructed from 490,319 UK Biobank genomes improves genotype imputation for global populations": ---

December 12, 2025

**5 Correlation $r^2$ in small cohorts and its dependence on estimation method 16**

**6 Supplementary Figures 18**

**7 Supplementary Tables 41**

### 1 Established imputation services

Genotype phasing followed by imputation of unmeasured genotypes improves statistical power in genome-wide association studies (GWAS) by massively increasing the genome-wide coverage of genetic variants. Established imputation services are the *Michigan Imputation Server* (MIS; `https://imputationserver.sph.umich.edu`; largest reference panel with 32,470 genomes of predominantly European ancestry from the Haplotype Reference Consortium (HRC) [1]; complete panel of HRC genomes; application for complete HRC data not possible), the *Sanger Imputation Service* (SIS; `https://imputation.sanger.ac.uk`; same reference panel as in the MIS), the *Kiel University Imputation Server* (*EagleImp-Web*; `https://hybridcomputing.ikmb.uni-kiel.de`), the *Helmholtz Imputation Server* (HIS; `https://imputationserver.helmholtz-munich.de` (largest panel is a subset of 27,165 genomes from the HRC panel; application for data possible via *The European Genome-Phenome Archive (EGA)*), and the *TOPMed Imputation Server* (*TOPMed*; `https://imputation.biodatacatalyst.nhlbi.nih.gov`), which has the so far largest imputation reference panel usable via imputation servers constructed from 133,597 genomes (*TOPMed r3* panel [2]).

Phasing of genome-wide study data, the computationally intensive step before genotype imputation, is conducted on the above mentioned imputation servers with the software *Eagle2* [3], which suffers from long runtimes with very large reference panels. This is why we developed the software *EagleImp* [4], in which we optimized *Eagle2* and combined it with a modified version of the imputation software *PBWT* [5] to enable phasing and imputation from very large reference panels with 1 million genomes and to achieve a 2-10 fold speed advantage over the original tools with same or improved phasing and imputation quality. For example, we previously showed that for input datasets from diverse human populations of the 1000 Genomes Project [6] *EagleImp* provides at least equal or better phasing and imputation quality than *Eagle2* and *PBWT* in terms of phasing switch error rate and imputation genotype error rate [4]. For common variants (minor allele frequency (MAF) $> 0.03$ and MAF $> 0.006$ depending on the benchmark dataset used in [4]), we were also able to show that *EagleImp* with the publicly accessible reference panel from the EGA release of the HRC1.1 panel (27,165 genomes) had the same or higher imputation quality (in terms of estimated imputation $R^2$) than the *SIS* (32,470 genomes; complete HRC r1.1 panel), the *MIS* (32,470 genomes; complete HRC r1.1 panel) and the *TOPMed Imputation Server* (133,597 genomes; *TOPMed r2* panel), despite their larger reference panels. *EagleImp-Web* utilizes *EagleImp* and supports several reference panels (e.g., HRC, 1000 Genomes Project), as detailed in **Supplementary Section 2**, **Supplementary Table 1**, and **Supplementary Figures 1–4**.

### 2 Implementation of *EagleImp-Web*

#### 2.1 *EagleImp-Web* server architecture used for benchmarks

An overview of the server architecture of *EagleImp-Web* used for imputation quality and runtime benchmarks of the UKB imputation reference panels (UK Biobank application number 139525) is depicted in **Supplementary Figure 1**. It is divided into two components, referred to as *frontend* and *backend* system, to make our service fast, stable and secure. The frontend hosts the web service including a database with user information, such as login credentials and all data regarding submitted jobs. Uploaded files and result files are also stored on this system. The frontend is equipped with an Intel Xeon Silver 4110 8-core CPU @ 3 GHz and 128 GB RAM. It offers more than 7 TB of redundant storage capacity and is currently running on an Ubuntu 24.04.2 LTS Linux system, which is regularly updated. The web service is mainly written in *PHP* and *JavaScript* and is hosted by an *Apache2 v2.4* [7] server. The database is implemented in *PostgreSQL* [8], version 16.8. For job queuing, we use *SLURM* [9], version 24.11.

With two Intel Xeon Gold 6538Y+ 32-core CPUs running at @ 2.2 GHz and 2048 GB RAM, the *backend* provides the necessary computing power for the actual processing of the submitted jobs. The operating system is Ubuntu Linux 24.04.2 LTS as well. The backend's main task is to run the *EagleImp* tool [4], version 2.0, to process the user's imputation jobs. *EagleImp* is written in *C++* and is freely available at *GitHub* (https://github.com/ikmb/eagleimp). It is based on the popular phasing tool *Eagle2* [3] and imputation tool *PBWT* [5], and combines both steps in a single application. The main advantages of *EagleImp* over the classical two step approach with *Eagle2* and *PBWT* are the increased computation speed of a factor 2 to 10 while the phasing and imputation quality is at least maintained or even improved.

To ensure direct communication without potential interception risks and routing problems, the frontend and backend are connected via a direct Ethernet connection and dedicated network devices. Communication between these systems via this connection is limited to two applications: First, the backend offers access to its *SLURM* daemon over this direct connection from the frontend. Second, the frontend only exports the storage file system via *NFSv4* to the backend via this connection and accepts communication to the *SLURM* process. A firewall on the frontend (implemented with the Linux system tool *iptables*) rejects all incoming traffic from the internet except *https* requests. (In particular, *http* requests are also allowed, but are automatically redirected to *https*.) The backend firewall is configured to completely block all incoming traffic from the internet. Only administrative access via *SSH* is exceptionally allowed only from the local network for both systems.

Since the servers are located in the infrastructure of Kiel University, an additional firewall from the university's router ensures that rules are not violated. The server's certificate (required for secure *https* connections) is issued via the University by the external organization *GEANT Vereniging* and will be verified by any browser's standard certificate chain.

#### 2.2 User interface and functionality

Due to its security and data protection architecture, *EagleImp-Web* helps your research project to become compliant with the European Union (EU) General Data Protection Regulation (GDPR). The conditions of the GDPR are explained transparently on our website.

The service runs on our university computers in Kiel, Germany. All website functions are carried out via encrypted and certified *https* connections. Apart from necessary session cookies on the user's computer, the website does not store any cookies and is free of advertising and does not collect any data for marketing purposes or for passing on to third parties. For the submission of jobs and the administrative functions, a user login with only minimal requirements is necessary, i.e. a valid email address and password. The email address is required for notifications about the user's jobs and is used for password recovery.

File downloads are handled directly on our server via certified *https* and can be started either directly in the protected user area on the website or from the command line from any remote terminal (e.g. via the download tool *wget*) protected with a one-time password. This eliminates the time consuming process of encrypting and decrypting the result files, because we assure authorized and exclusive access at the same time and, in contrast to other imputation web servers, we guarantee that input and output data is securely stored temporarily and exclusively on our own university computers in Kiel, Germany, and will never be passed on to third-party services (such as *Globus* (used by the *SIS*) or *Amazon* cloud servers (used by *MIS* and *TOPMed*)), or transferred to an unknown location. Data access is only possible for the authorized user. Additional security is provided by optional 2-factor authentication with the possibility to register different devices (e.g. passkeys unlocked via smartphone with fingerprint authentication or face recognition, or a USB security key dongle).

An overview of the workflow for running a *EagleImp-Web* job is depicted in **Supplementary Figure 2**.

##### 2.2.1 Registration and login

*EagleImp-Web* requires the registration of a personal user account including a valid email address. The user account enables the protection against unauthorized access by third-parties to user data and test results. The email address is used to inform the user about her/his job events (such as a job completion, because genome-wide imputation processes may take several hours). We explicitly point out that we do not use the provided email address for purposes other than job notifications and account management and do not collect any usage information or statistics of our service in connection with user accounts.

To complete the registration process, the user gets a verification email from our server to the provided email address. (For sending mails the tool *mSMTP* is used.) The email contains a one-time link that finally validates the email address and activates the user account. It is valid for 7 days, after which the account will automatically be deleted if it was not activated before.

The account protection is implemented either via a simple password or optionally for extra security, the user may register passkeys or key devices for 2-factor authentication (see Sect. 2.2.5 below).

After login, the user is able to submit jobs, manage ongoing or completed jobs, download results, and to manage the account. Note, that the web service will logout the user automatically after 30 minutes of inactivity.

In general, we renounce the usage of cookies, but to verify the current login status we need to store a necessary session cookie.

##### 2.2.2 Job submission

New jobs are arranged in a queue to ensure a fair order of execution among users (on a first-come, first-served basis). To run a job within our service, the upload of genotype data in VCF or BCF format (`.vcf.gz`/`.bcf`) is required. A screenshot of the job submission form is depicted in **Supplementary Figure 3**. Uploading files is possible in three different ways. The easiest way is to upload via a browser. Files can simply be selected in a file selection dialog or dragged and dropped in the designated box. Alternatively, the web server can actively fetch files from public URLs (e.g. pointing to a private server) that are provided in the submission form. Or the upload via *Secure File Transfer Protocol (SFTP)* can be chosen, which requires the user to provide the host URL, login credentials and relative paths to the files on that server. Note that we use the login credentials only for the purpose of downloading the files. We never submit them in plain-text, as

SFTP is an encrypted connection, and we delete them immediately after the upload to our server has finished.

During the upload process the files are checked for consistency to improve reliability and ensure stable job runs. In particular, in the case of a browser upload, a JavaScript module executed on the client's device checks selected files already before upload. The user gets an immediate feedback about files not matching the required restrictions:

- Only VCF or BCF files with the suffix `.vcf.gz` or `.bcf` are allowed.
- All files must start with a valid human chromosome number (1-24,X,Y) followed by a dot ".", a prefix `chr` is allowed.
- Only one file per chromosome is allowed.
- The maximum size of a file must not exceed 1,000 MB.
- The number of samples in a file must not exceed 10,000, if you want to use the complete UKB reference panel. For the rpMAF-filtered UKB panel 40,000 samples are allowed, and for the traditional 1000 Genomes or HRC reference panels 100,000 samples are allowed.
- Each file must contain the same number of samples. (We do not check the sample IDs though.)

If the user selects another upload method, the files are checked on our server while uploading and the upload stops immediately if a file is not valid.

When submitting the job, the values in the form are translated by a PHP script in the background to the command line options required for the *EagleImp* software. The relative location of the job folder, that is uniquely created for the input, output and log files, is submitted along with the command-line options to the job queuing system *SLURM* (see Sect. 2.2.3).

The queuing status and progress of the job can be supervised in the *Jobs* section. Once a job has finished, the web service gets notified via a secured notification URL (allowing only connections from the backend system). According to the jobs return state (success or failed), the web service generated download links for the result files and notifies the user per email (see Sect. 2.2.4 for details).

##### 2.2.3 Job queuing system

For job queuing we use the freely available process management software *SLURM* [9], version 24.11. *SLURM* is configured to use the complete backend system exclusively for each *EagleImp-Web* imputation job, i.e. the call to the *EagleImp* launch script is enqueued in the *SLURM* queue and executed on the backend system whenever its resources are free. The parameters applied to the launch script are generated from the user options in the *EagleImp-Web* user interface together with the (relative) location of the user's input data. The script converts the relative location to an absolute path and launches the *EagleImp* executable for the user's input files with added default and user options.

Each *SLURM* job is configured with an epilog command that is executed when the job terminates. It is used to notify the web service about the end of the job processing (either successful or not). This invokes several operations on the frontend, such as changing the job state, notifying the user and preparing the download URLs.

###### 2.2.4 Job management and results download

After submission, the job is queued in the user's job queue and can be managed in the "Jobs" section. Each job is classified into four main categories: *queued*, *running*, *terminated* or *retired*, whereby *terminated* can be one of the subcategories *succeeded*, *failed* or *canceled*. The jobs are listed in chronological order along with a waiting status indicating the position in the job queue. A single user can queue up to three jobs at a time (i.e. no more than three jobs can be queued waiting to be processed by a single user, however, there is no limit to the total number of jobs for a user).

**Supplementary Figure 4** shows a screenshot from an exemplary job progress. Once a job is actively being processed, the user can monitor the current progress and view information about the job, such as the elapsed runtime, warning and error messages. When a job is regularly completed, it is classified as *succeeded* and the result files can be downloaded. A job can also be canceled by the user, in which case it is stopped immediately and classified as *canceled*. If an error occurs, job processing stops and it is classified as *failed*. However, log files of the job execution can still be downloaded even for failed or canceled jobs.

For each successfully finished job the web service generates `.vcf.gz` or `.bcf`-files that contain the results of the phasing and/or imputation steps for each uploaded chromosome file (according to the selected user options). For each file, the MD5 hash is calculated such that downloads can be easily verified. A `.varinfo`-file contains additional information for each input variant, i.e. the mapping to the reference (e.g. if it was excluded, ref/alt swapped, original variant ID vs. reference variant ID, etc.). If phasing is done, the phasing confidences are also provided in a separate file. In addition, the *EagleImp* execution log can be found in a `.log`-file.

The result files are registered in our database with a unique random string for each file. The web service then generates a secure download URL for each file over an encrypted *https*-connection based on this random string. We use the *rewrite engine* by the Apache server to decode this URL to redirect a download request to our download engine, and the download engine queries the database for the associated file.

The user may download each result file separately from the web browser, but an individual script to download all available files at once from a (Linux) command line terminal is also provided to the user. For this purpose, the user receives a notification email for all completed, canceled or otherwise terminated jobs, containing a generated password uniquely associated with that job. The files are locked by default and a direct download via the file URL is only possible when the user is either logged-in to download individual files directly from the website or the received password is entered via the download script on the command line to authenticate the download. Technically, the password has to be sent as a *POST* command after establishing the encrypted *https* connection to the web server to download a file, which is done automatically by the provided script. This is advantageous if the result files have to be analyzed or post-processed on a different system than the user's computer. This way, separate encryption of the result files is not required and saves the user time by not having to decrypt the files before using.

Note that we automatically retire a job after 7 days after the job has terminated. In that case all data related to a job (with the exception of status and log files) is deleted from our server. We keep status and log files for the user to be able to recall information on its previously run jobs. Of course, any job independent of its status can be deleted completely manually by the user. Anyway, retired jobs older than one year are also automatically removed completely.

##### 2.2.5 Account management

As stated above, a valid email address and a password are required to set up a personal user account. The password is not stored in plain text, but as a secure SHA-256 hash on our server. Users are able to change their email address and password at any time in the account management section.

*EagleImp-Web* also provides a password recovery function to set a new password in the case the user has forgotten the login password. The user can click on the link "Forgot your Password?" below the password field and enter the email address and a captcha code displayed as image, which is necessary to prevent abuse by bots. If the captcha code is correct and the email address is registered in our database, a link to reset the password is sent to the user. After clicking the link, a new random password is sent to that email address. The user can now sign in again and change the password.

Optionally, to improve account security, the user can activate 2-factor authentication for her/his account based on the recent standard *Web Authentication API (Webauthn)* [10]. Webauthn enables usage of hardware authenticators with public and private key-based credentials (passkeys) to perform an SSL handshake between server and the client's authenticator as trusted device. A trusted device may be a USB key dongle, fingerprint reader or facial recognition on a phone or any other applicable device. In detail, an account which is protected by 2-factor authentication requires the correct password and the correct authentication of one of the registered trusted devices to verify the user's identity upon login. Note, that after the registration of a trusted device, the login is not possible from clients where not at least one passkey is available, e.g. if a smartphone's integrated fingerprint reader is used as a trusted device, the login is not possible from devices where the user cannot verify the login request on that smartphone. Also note, that once 2-factor authentication has been activated, it is no longer possible to log in without a trusted device. However, 2-factor authentication can be disabled by the user itself after login by removing all trusted keys in its account settings.

The user may also delete its account in the account settings, which results in an immediate deletion of all data associated with this account from our server without exception. Specifically, the login credentials are removed from our database together with all uploaded files and data created for this user as a result from using the service.

### 3 Creation of the UKB reference panel

#### 3.1 Data preparation of WGS data on the *UK Biobank Research Access Platform (RAP)*

The GraphTyper population level whole-genome sequencing (WGS) variants from the 500k release are exclusively available in pVCF format on the *UK Biobank Research Access Platform (UKB RAP)* (https://ukbiobank.dnanexus.com). We created a tool set *Vcffilter* written in C/C++ (https://github.com/ikmb/vcffilter) to extract all necessary information required to build a reference panel from the pVCF files on the UKB RAP, and to quickly apply further filtering and restoration of valid VCF files for phasing on our own compute resources.

*Vcffilter* requires only a valid uncompressed VCF file stream as input to generate the reduced output, which we compressed again for download from the UKB RAP. For download verification we used an MD5 checksum.

```
zcat <input.vcf.gz> | vcffilter | gzip > <reduced_input.gz>
md5sum <reduced_input.gz> > <reduced_input.gz>.md5
```

The call is part of a bash script which launches the extraction on four different pVCF files at once. The script is executed as part of multiple *"Swiss Army Knife"* application launches on low-priority 4-core requests of type *mem2_ssd1_v2_x4* on the UKB RAP. (The `--tag` and `--name` options are only used for organization purposes.)

```
dx run app-swiss-army-knife -icmd="bash <script> <files>" \
    --instance-type mem2_ssd1_v2_x4 --tag <some_tag> --name <some_name> \
    --priority low -y
```

#### 3.2 Restoring and quality control (QC) of Graphtyper VCF data

For further processing, we used our own compute resources, thus we needed to download the reduced data files to our own servers. Downloading from the RAP was conducted using the `dx download` command. Subsequently, we checked the MD5 checksum.

```
dx download -f --no-progress --lightweight <files>
md5sum -c <file>
```

For restoring a valid VCF file with on-the-fly QC-filtering, we used our custom tool *restorevcf* from the *Vcffilter* tool set (see above). We applied options to (a) filter all variants with a value other than `PASS` in the `FILTER` column, (b) a missingness rate of greater than 10%, (c) an alternative allele score (`AAScore`) of less than 0.8, and (d) with unknown alleles named "*". Further, we applied the option to split multi-allelic variants to several bi-allelic ones in the same call as if *bcftools* was applied with `bcftools norm -m`. This option includes the removal of all unnecessary information from the `INFO` column and recalculation of the allele frequency (`AF`), allele count (`AC`) and allele number (`AN`) for all variants. However, we forced to keep the `AAScore` in the `INFO` column. As we need two different batches of files derived from the originals ((i) filtered by MAF $\geq 0.001$ and (ii) filtered by MAC $\geq 4$), we applied the restoration command twice with different MAF/MAC filter options to each reduced file. As restoration to a valid VCF file requires a separate VCF header file, we restored a temporary uncompressed VCF without header first.

```
zcat <reduced_input.gz> | \
    restorevcf --fpass --missfilter 0.1 --aafilter 0.8 --filterunknown \
               --splitma --keepaa --maffilter 0.001 > <uncompressed_maf0.001>
zcat <reduced_input.gz> | \
    restorevcf --fpass --missfilter 0.1 --aafilter 0.8 --filterunknown \
               --splitma --keepaa --macfilter 4 > <uncompressed_mac4>
```

Next, we used an extracted header (`bcftools view -h`) from an arbitrary pVCF file on the UKB RAP (as all pVCF files contain the same header) and applied it to each uncompressed file with subsequent compression to finalize the restoration and QC of valid VCFs. In particular, we used the `.bcf` file format for further processing, generated with *bcftools* and deleted the temporary uncompressed file.

```
cat header.vcf <uncompressed_file> | \
    bcftools convert - -Ob -o <restored.bcf>
rm uncompressed_file
```

##### 3.3 Phasing of common variants

For phasing the common variants in our first batch of 20 kbp-chunk files, we created chunks of size around 20-30 Mbp for each chromosome, including an overlap of around 2 Mbp, by simply concatenating the corresponding files using *bcftools* and chunk limits corresponding to O. Delaneau in `https://github.com/odelaneau/shapeit5/tree/main/tasks/phasingUKB_200k_release/autosomes/chunks.tgz`.

```
bcftools concat --naive-force <list of chunk files> -o <concat file>
bcftools index <concat file>
```

Phasing is then conducted by calling the *SHAPEIT5* [11] tool *phase_common* with the default options. The genetic map file is used as provided together with the *SHAPEIT5* resources.

```
phase_common --input <concat file> --map <genetic map file> \
             --output <phase_common output> --region <chr>
bcftools index <phase_common output>
```

Note, that we also used multi-threading with the `--thread` or `--threads` switches for *SHAPEIT5* or *bcftools* were applicable and our compute resources allowed it.

##### 3.4 Ligation and merging of chunks to create a scaffold

Ligation uses the overlap between chunks to decide which phases need to be switched for which samples in each chunk. We used the ligation option `-l` provided by *bcftools* to ligate and merge the chunks in one step.

```
bcftools concat -l -c <list of concat files> -Ob -o <scaffold file>
bcftools index <scaffold file>
```

##### 3.5 Phasing of rare variants

We used the second batch of our input files with variants with MAC $\geq 4$ for phasing the rare variants and created chunk files of approximately 5 Mbp without overlaps.

```
bcftools concat --naive-force <list of chunk files> -o <concat file>
bcftools index <concat file>
```

The call to phase rare variants requires an accurate declaration of the region to be phased (i.e. the region spanned by the chunk file) and the created scaffold with a declared region with some overlap (approx. 500 kbp) left and right to the region in the chunk file.

```
phase_rare --input <concat file> --map <genetic map file> \
           --output <phase_rare output> \
           --input-region <region of concat file> \
           --scaffold <scaffold file> \
           --scaffold-region <region of concat file +- 500 kbp>
bcftools index <phase_rare output>
```

##### 3.6 Concatenation of chunks and creating *Qref* reference panel files

To create the final phased chromosome file ligation of the phased chunks from the previous step is not required because the phase is chosen according to the scaffold which was the same for each chunk. Thus, a simple concatenation of the 5 Mbp-chunks is sufficient to create the phased chromosome file.

```
bcftools concat --naive <list of chunk files> -o <phased_chromosome>
bcftools index <phased_chromosome>
```

The generated file could now be used directly as a reference panel file for imputation. However, as handling VCF files of this size is extremely slow, we converted the phased chromosome files to our *Qref* format to be used with *EagleImp*.

```
eagleimp --makeQref --ref <phased_chromosome>
```

##### 3.7 Special treatment of chromosome X

To take account for male (haploid) samples in the *nonPAR* region (chrX:2781480-155701382) of chromosome X, we handled the three regions *PAR1*, *nonPAR* and *PAR2* separately. We phased the *PAR1/2* regions as described above, with the exception that both regions could be phased directly in one chunk (common as well as rare variants), such that chunking and ligating or concatenation was not required.

For the *nonPAR* region, we applied chunking as above and left the original encoding for haploid males as homozygous diploid as phasing with *SHAPEIT5* ignores the haploid encoding and again generates a homozygous diploid output anyway. We used the standard *bcftools* plugin *fixploidy* to fix the ploidy in the final phased output for each chunk. Due to a potential removal of alternative alles by this procedure, we needed to correct the reported allele counts (`INFO/AC` field) and allele number (`INFO/AN` field). We used the *bcftools* plugin *fill-tags* for this procedure. Finally, we filtered again to be consistent in keeping only variants with MAC $\geq 4$.

```
bcftools +fixploidy <phased_chromosome> -- -s <list of samples with gender>
         -p <ploidy file> | \
    bcftools +fill-tags -- -t AC,AN | \
    bcftools filter -e 'INFO/AC<4 || INFO/AC>INFO/AN-4' \
         -Ob -o <phased_chromosome_fixedploidy>
bcftools index <phased_chromosome_fixedploidy>
```

The ploidy file used in the previous call simply contains an entry for the *nonPAR* region where male samples have to be haploid:

```
chrX    2781480 155701382       M       1
```

Finally, there are three reference panel files for chromosome X (one for each *PAR1/nonPAR/PAR2* region) which we converted to the *Qref* format as before for imputation with *EagleImp*.

```
eagleimp --makeQref --ref <phased_chromosome>
```

### 4 Supplementary benchmark information

#### 4.1 Benchmark datasets

For the benchmarks we used data from mainly two sources. First, we used publicly available genome data from 2504 individuals from the 1000 Genomes Project [6]. These individuals are distributed over 26 populations (also referred to as subpopulations) and 5 superpopulations. We used the *UCSC liftOver* tool [12] to convert the original genome data from GrCH37 to GrCH38. From this, we extracted only variants that can be typed by Illuminas *Global Screening Array (GSA)* [13]. Next, we created 31 benchmark datasets by (a) extracting the individuals for the five superpopulations, and (b) for the 26 subpopulations.

Second, we used in-house GWAS data sets for COVID-19 studies from four populations: Germany, Italy, Norway and Spain [14, 15], whereby the datasets for Italy and Spain are from the first study in [14].

**Supplementary Table 3** lists all benchmark datasets. The datasets from the 1000 Genomes Project data can be downloaded at https://hybridcomputing.ikmb.uni-kiel.de/imputation-benchmark.

#### 4.2 Imputation quality

For all described benchmark datasets we conducted imputation with *EagleImp* and default parameter $K = 10,000$ using the complete UKB reference panel (UKB complete) and the rpMAF-filtered UKB reference panel (UKB MAF0.0001) which was filtered for reference panel MAF $\geq 0.0001$.

For comparison of our benchmark results to *TOPMed* imputation results, we conducted imputation for the same described datasets with the *TOPMed Imputation Server*.

Note that both, *EagleImp* and the *TOPMed Imputation Server*, generate an imputation output containing dosage values for every variant present in the reference panel. Thus, neither method produced missing imputed values in our benchmark analyses.

##### 4.2.1 Estimated imputation $R^2$

In *EagleImp*, the estimated imputation $R^2$ is calculated as suggested by Das et al. [16]. Correlation can be measured by the squared *Pearson Correlation Coefficient* if the ground truth is known ($X$ defines the true alleles while $Z$ presents the observation (imputation)):

$$r^2 = \frac{(\mathrm{Cov}(X, Z))^2}{\mathrm{Var}(X)\mathrm{Var}(Z)} \tag{1}$$

Under the assumptions of Hardy-Weinberg equilibrium, Das et al. showed that this can be estimated as ($n$ is the number of records, $z_i$ are allele dosages):

$$r^2 \approx R^2 = \frac{n \sum z_i^2 - \left(\sum z_i\right)^2}{n \sum z_i - \left(\sum z_i\right)^2} \tag{2}$$

The computed $R^2$-values are stored in the `INFO/R2` field for each variant in the VCF-file output. The same calculation is used by common tools, i.a. *minimac4* [17] which is also used by the *TOPMed Imputation Server*.

We extracted the estimated imputation $R^2$ values for all variants as reported in the `INFO/R2` field from all resulting VCF-files (separately for *EagleImp* and *TOPMed*) together with the reference panel allele frequency. Unfortunately, in contrast to *EagleImp*, *TOPMed* does not report the reference panel minor allele frequency (rpMAF) of the *TOPMed r3* panel, which is why we used the rpMAF from our UK Biobank reference panel for all analyses involving MAF.

To calculate the mean of the reported $R^2$ values for the UKB reference panels, we summarized the $R^2$ values and divided by their number for (a) all variants with an rpMAF $\geq 0.0001$, (b) all variants with an rpMAF $\geq 0.001$ and (c) all variants with an rpMAF $\geq 0.01$. The results are listed in **Supplementary Table 4**. We observed that the difference in mean $R^2$ between both variants of the UKB panel is negligible.

For visualization of the estimated $R^2$ distribution, we created bins for logarithmically equally distributed rpMAF ranges and calculated the mean $R^2$ for each bin, representing the data points in an $R^2$-plot. We plotted the $R^2$ distribution for the datasets of the 1000 Genomes Project super-populations for all variants in the UKB complete and UKB MAF0.0001 panels in **Supplementary Figure 6**.

For the comparison to the *TOPMed Imputation Server*, we extracted from all reported $R^2$ values (*TOPMed* and UKB complete) only those $R^2$ values that stem from variants shared in both panels (we also considered potential reference/alternative allele swaps). The number of shared variants in the results is 200,233,690. The calculation of the mean $R^2$ is then conducted as before, but based only on the shared variants. As there is no reference panel minor allele frequency (rpMAF) available for the *TOPMed* results, we distributed the results in the three categories based on the rpMAF of the UKB panel. The results are listed in **Supplementary Table 5**.

Corresponding to the $R^2$-plots for the UKB panels, we visualized estimated $R^2$ distribution for the complete UKB panel and the *TOPMed* results based on shared variants in **Supplementary Figure 7**.

We visualized the same $R^2$ distributions for the COVID-19 GWAS datasets (of GSA-typed samples) in **Supplementary Figures 23** (comparison of both UKB panels) and **24** (comparison to *TOPMed*).

###### 4.2.2 Switch error rate (SER)

We compared the phased and imputed samples from our 1000 Genomes Project benchmark datasets to the 1000 Genomes reference, where they originate from (ground truth). The haplotypes from Phase 3 of the 1000 Genomes Project [6], which were generated using a family-based scaffold, are widely used as ground truth in phasing benchmarking studies (e.g. [18]) and in studies that re-phase these data while maintaining consistency with the established scaffold (e.g. [19]). We only focus on the extracted GSA variants which were phased during the imputation preprocessing (referred to as *typed* variants). We define a *phase switch* whenever the current phase at a (heterozygous) typed site differs from the phase at the previous (heterozygous) typed site. We count a *switch error* whenever a phase switch occurs at a typed site in the phased sample, but not in the the same original sample in the reference (with the correct phase known), or vice versa. The *switch error rate (SER)* per sample is then computed by dividing the switch errors by the number of all typed variants. The SER for a complete dataset is then calculated as the average over all samples in the dataset. Please note that switch errors in the reference cannot be completely ruled out, since the publicly available reference data was phased by algorithms themselves.

Switch error rates for our 1000 Genomes Project benchmark datasets phased and imputed with *EagleImp* and the *TOPMed Imputation Server* are listed in **Supplementary Table 6**. An overview is illustrated in **Supplementary Figures 8** and **9** for *EagleImp* imputation of the superpopulations using the complete UKB reference panel (UKB complete) and the rpMAF-filtered UKB reference panel (UKB MAF0.0001) respectively. In **Supplementary Figures 10** and **11** an overview of *EagleImp* imputation for the subpopulations with both panels is shown respectively.

###### 4.2.3 Mean absolute error (MAE)

The *absolute error* of an imputed genotype is its absolute difference to the known genotype from the ground truth. As imputation usually sets a hard called genotype according to the computed allele dosages, we calculate absolute errors based on the imputation allele dosages for a better accuracy. When genotypes are encoded as 0 (homozygous reference), 1 (heterozygous) and 2 (homozygous variant), the absolute error for the homozygous calls can in general simply be computed by the absolute difference to the sum of the allele dosages ($g$ is the genotype, $d^m$ and $d^p$ are the dosages for the maternal and paternal strands respectively):

$$e(g, d^m, d^p) = |g - (d^m + d^p)| \qquad \text{for} \qquad g \in \{0, 2\} \tag{3}$$

However, for heterozygous genotypes the sum of allele dosages does not necessarily reflect a correctly imputed heterozygous genotype. For example, let $d^m = 1.0$ and $d^p = 0.0$. According to Eq. 3 the error would be $e = 0$, which is correct. But for the case $d^m = 0.6$ and $d^p = 0.4$, the error would equally be $e = 0$, which is obviously wrong. In the latter case, imputation clearly produced at least a 0.4 deviation for each strand (as one strand is 1 and the other is 0), depending on the phase, resulting in an error of $e = 0.8$. As we do not consider phase errors for calculating genotype errors, we computed the minimum of the sum of deviations of allele dosages to both strands:

$$e(g, d^m, d^p) = \min\left(\ 1.0 - d^m + d^p\ ,\ 1.0 - d^p + d^m\ \right) \qquad \text{for} \qquad g = 1 \tag{4}$$

The *mean absolute error (MAE)* accordingly is the average absolute error over all called genotypes:

$$MAE = \frac{1}{n} \sum e_i \tag{5}$$

For presentation, we stratified MAE for different rpMAF categories (non-cumulative). The values are listed in **Supplementary Tables 7** and **8** for *EagleImp* imputation using the UKB complete and UKB MAF0.0001 reference panels respectively. Stratified MAE in comparison to *TOPMed* is visualized for each 1000 Genomes super- and subpopulation in **Figure 2** in the main paper (stratified MAE for 1000 Genomes Project superpopulations imputed with *EagleImp* and the UKB MAF0.0001 reference panel) and **Supplementary Figures 12**, **13** and **14** (stratified MAE for 1000 Genomes Project super- and subpopulations imputed with *EagleImp* and the UKB complete panel, and stratified MAE for 1000 Genomes Project subpopulations imputed with *EagleImp* and the UKB MAF0.0001 panel respectively).

###### 4.2.4 True correlation $r^2$

To measure the true correlation between an imputed dataset and the known ground truth, Eq. 1 is used (originally taken from Das et al. [16]). Given $X$ as the set of expected haplotypes from the ground truth and $Z$ as the set of observed (imputed) haplotype dosages, correlation $r^2$ can thus be defined as the squared *Pearson Correlation Coefficient*:

$$r^2 = \frac{(\mathrm{Cov}(X,Z))^2}{\mathrm{Var}(X)\mathrm{Var}(Z)} \tag{6}$$

$$= \left(\frac{\mathrm{Cov}(X,Z)}{\sigma(X)\sigma(Z)}\right)^2 \tag{7}$$

$$= \mathrm{PCC}(X,Z)^2 \tag{8}$$

As for the switch error rate (SER) and the mean absolute error (MAE), we used the 1000 Genomes Project reference, from where we extracted our benchmark datasets, as ground truth.

Let $n$ be the number of sample points (expected and observed values respectively), $x_i$ and $z_i$ the $i$-th sample points ($z_i$ the $i$-th dosage), and $\overline{x}$ and $\overline{z}$ the sample means, then PCC can be generally defined as:

$$r = \frac{\sum (x_i - \overline{x})(z_i - \overline{z})}{\sqrt{\sum (x_i - \overline{x})^2}\sqrt{\sum (z_i - \overline{z})^2}} \tag{9}$$

$$= \frac{n\sum x_i z_i - \sum x_i \sum z_i}{\sqrt{n\sum x_i^2 - (\sum x_i)^2}\sqrt{n\sum z_i^2 - (\sum z_i)^2}} \tag{10}$$

Squaring leads to:

$$r^2 = \frac{(n\sum x_i z_i - \sum x_i \sum z_i)^2}{\left(n\sum x_i^2 - (\sum x_i)^2\right)\left(n\sum z_i^2 - (\sum z_i)^2\right)} \tag{11}$$

We implemented above formula (Eq. 11) to calculate $r^2$ as it provides a convenient single-pass way where the sample means do not have to be known in advance. However, this method may be numerically instable especially in cases where small numbers (e.g. distances) have to be squared and accumulated. We solved this by handling accumulations in batches.

We observed that when calculating correlation $r^2$ on a per variant basis, the sample size is often too small to get an unbiased result and, even more important, in cases of low or no variance, $r^2$ can simply not be calculated (due to a zero denominator). Especially for variants with a very low MAF we lost a significant proportion of $r^2$-values (especially for UKB imputations) even when such variants were perfectly imputed with no errors, but exhibited no variation in the target samples (e.g. because MAF = 0). In these cases, the denominator of the correlation $r^2$ formula is zero, leading to an undefined $r^2$ which we set to zero to be able to calculate a consistent average. We present these results for our 1000 Genomes Project benchmark datasets imputed with *EagleImp* and the *TOPMed Imputation Server* in **Supplementary Figures 15** and **16** for the superpopulations from the 1000 Genomes Project (*EagleImp* results were imputed with the UKB complete and UKB MAF0.0001 panel respectively). Likewise, **Supplementary Figures 17** and **18** show the results for the subpopulations from the 1000 Genomes Project. The computed values can be found in **Supplementary Tables 9** and **10**.

Because of above mentioned problems, we computed correlation $r^2$ also over the total number of observations over all samples and variants (classified in bins of different allele frequencies). The final reported correlation $r^2$ for a bin over a complete dataset is then the average over all corresponding bins from the chromosome files, weighted by the number of variants in each chromosome file. These correlation $r^2$ values for our 1000 Genomes Project benchmark datasets imputed with *EagleImp* and the *TOPMed Imputation Server* are listed in **Supplementary Tables 11** and **12** respectively for *EagleImp* imputation with the UKB complete and the UKB MAF0.0001 panels. Graphical representations are illustrated in **Supplementary Figures 19** and **20** for the superpopulations from the 1000 Genomes Project, and **Supplementary Figures 21** and **22** for the subpopulations each respectively for UKB imputation with the UKB complete and UKB MAF0.0001 panels.

##### 4.3 Runtime

Benchmarks for the UKB panels were performed on the backend system of *EagleImp-Web* (**Supplementary Figure 1**) equipped with two Intel Xeon Gold 6538Y+ CPUs @2.2GHz with a total of 64 cores (128 threads) and 2048 GB of RAM. *EagleImp* was started for each chromosome file set as in *EagleImp* with a 2x8x16 configuration, which means that sixteen runs are executed in parallel, but each two runs exclusively share 16 threads for compute intensive task such as phasing and imputation (file reading and parsing can be conducted by all 16 executions in parallel). The runtime is measured by taking the difference between the start timestamp (at the beginning of processing a dataset, which is the start of executing the *EagleImp* launch script) and the end timestamp (when the last result file of a dataset has been written and the *EagleImp* launch script has terminated).

We conducted the same performance benchmarks with the same datasets on the *UKB-RAP* with *EagleImp-RAP* using the *mem3_ssd1_v2_x16* instance type (as this allows to execute *EagleImp* with the same amount of threads (16) used for compute intensive operations as in above configuration for our benchmark system). For this, we uploaded the data files to the *UKB-RAP* using `dx upload`. Then, for each population dataset, we created a batch input file using

```
dx generate_batch_inputs -i target='(chr.*).*.vcf.gz' -o <batchfile>
```

in the input directory. We used a local installation of *EagleImp-RAP* and started it with:

```
dx run <path-to-applet>/ikmb-eagleimp --batch-tsv <batchfile>
       -ireference=<UKBcomplete/UKBmaf> -ibuild=hg38 -iallowRefAltSwap=true
       -iimputeInfo=agp --instance-type=mem3_ssd1_v2_x16 --priority=high
```

Note, that we had to have the reference panel files installed locally in our project. And as this launches all chromosome file jobs at the same time, we reported the runtime of the longest job (which was chromosome 2 with no exception) as total runtime.

Runtimes for *TOPMed* have been taken as reported by the server (not including queuing times). The execution platform and order of parallelism is unknown.

The runtimes are listed in **Table 3** in the main manuscript.

#### 5 Correlation $r^2$ in small cohorts and its dependence on estimation method

As expected, calculations based on the correlation $r^2$ with $r^2$ values averaged across individually calculated $r^2$ for each variant showed correlation $r^2$ for both UK Biobank reference panels and the *TOPMed r3* panel (**Supplementary Tables 9 and 10**, upper panel; **Supplementary Figures 15 and 16**) consistent with previous correlation $r^2$ benchmarks (also $r^2$ values averaged across variants) based on 500 proxy individuals from the *TOPMed r3* panel assigned to the five 1000 Genomes Project superpopulations [2]. However, the correlation $r^2$ values calculated separately for the 26 subpopulations (**Supplementary Tables 9 and 10**, middle panel; **Supplementary Figures 17 and 18**) showed a weakness of the correlation $r^2$ measure for imputation quality for low allele frequencies, leading to potentially biased results due to small sample sizes especially for rpMAF range categories $[0.1\%, 1\%)$ and $[0.01\%, 0.1\%)$ and because in cases with small or no variance the correlation $r^2$ simply cannot be calculated due to a denominator of zero; thus, setting the correlation $r^2$ to 0 for perfectly imputed genotypes for all benchmark samples leads to (disproportionately) lower average correlation $r^2$ values (see detailed explanations and formulas for correlation $r^2$ in **Supplementary 4.2.4**). From a population genetics perspective, for example, it is also unlikely that the imputation quality at $[0.01\%, 0.1\%)$ for the rpMAF-filtered UKB reference panel (or *TOPMed r3* panel) for GBR is worse than for YRI, with a correlation $r^2$ value of 0.04 (0.04) for GBR compared to YRI 0.40 (0.44) (**Supplementary Tables 9 and 10**, middle panel; **Supplementary Figures 17 and 18**), knowing that 93.85% of UK Biobank participants reported their ancestry as "White British" and only 1.56% of participants reported their ancestry as "Black" or "Black British" (**Supplementary Table 2**). In an attempt to provide enough data points to calculate correlation $r^2$ for the 26 subpopulations individually, we calculated correlation $r^2$ for all three imputation reference panels additionally based on the total number of observations across all samples and variants per rpMAF range category (instead of correlation $r^2$ averaged across variants) showing high correlation $r^2$ values for all 26 subpopulations (**Supplementary Tables 11-12**; **Supplementary Figures 19-22**), regardless of whether the genetic similarity between our three reference panels and the target (input) subpopulation benchmark dataset was high or low. These results from correlation $r^2$ would imply that the imputation of genotypes appears to be much easier for low-frequency variants than for common variants in the case that the study population and individuals from the reference panel are poorly matched. However, since the algorithm most frequently estimates the more frequent allele for low rpMAF categories purely by chance and thus correlation variance is usually low just by chance, this often leads to a biased correlation $r^2$ for low range rpMAF categories (and a false and underestimated correlation $r^2$ set to 0 for perfectly imputed genotypes, see text above). Note that smaller imputation errors for low-frequency variants than for common variants were also observed in calculations based on MAE (**Figure 2** in the main paper; **Supplementary Figures 12-14**), but with a smaller bias because the difference between the estimate and the ground truth was not measured on the basis of variances. For this reason, we recommend the measure of MAE for imputation quality (which is not based on correlation) as a secondary metric for benchmark analyses with small ground truth datasets such as the 1000 Genomes Project data, and in general the measure SER for benchmarks of phasing quality, as well as the estimated $R^2$ (available for all variants of a reference panel) for post-imputation filtering of poorly imputed variants from GWAS input datasets.

### 6 Supplementary Figures

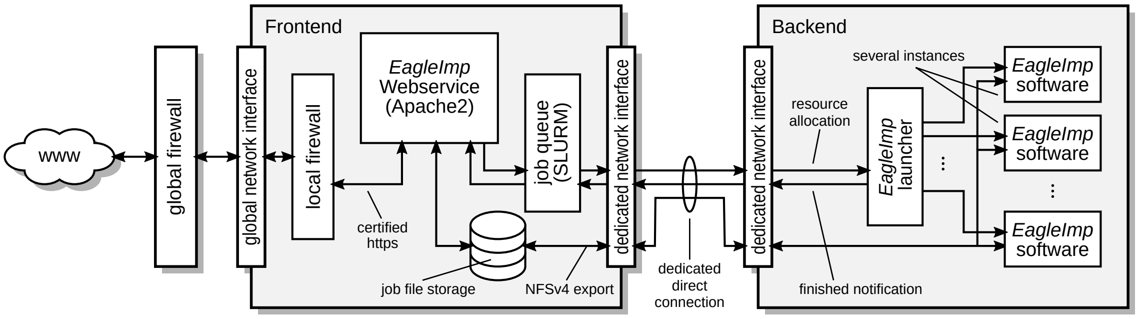

Supplementary Figure 1: The *EagleImp-Web* server architecture consists of two main components: the frontend and backend systems. The web service is hosted by the frontend while jobs are processed on the backend system. The processing order is controlled by the job queuing software *SLURM*. Files are stored on the frontend and accessed via an *NFSv4* network file system.

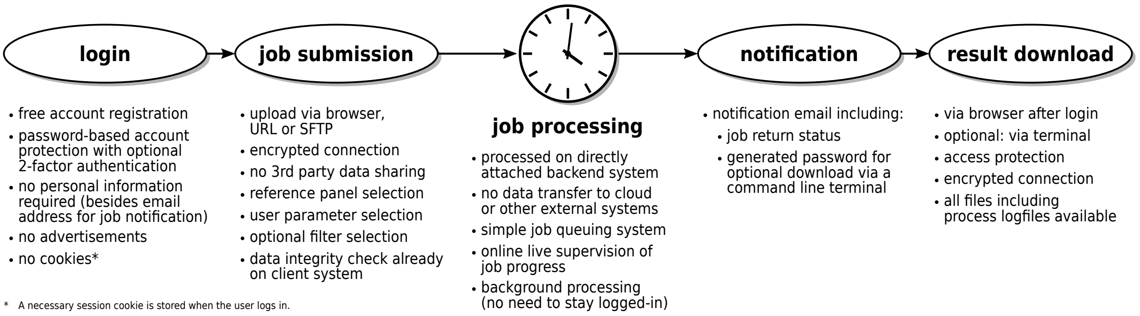

Supplementary Figure 2: Overview of the job processing in *EagleImp-Web*, including key technical details on security and data protection. After registering an account and logging in, the user uploads a VCF/BCF file dataset and completes the job submission form by selecting the desired reference panel, algorithm options and optional filters. The job is then queued and processed on the directly connected backend compute system. Once the process is complete, the user receives an email notification. Result downloads are possible either via the browser or a command line terminal.

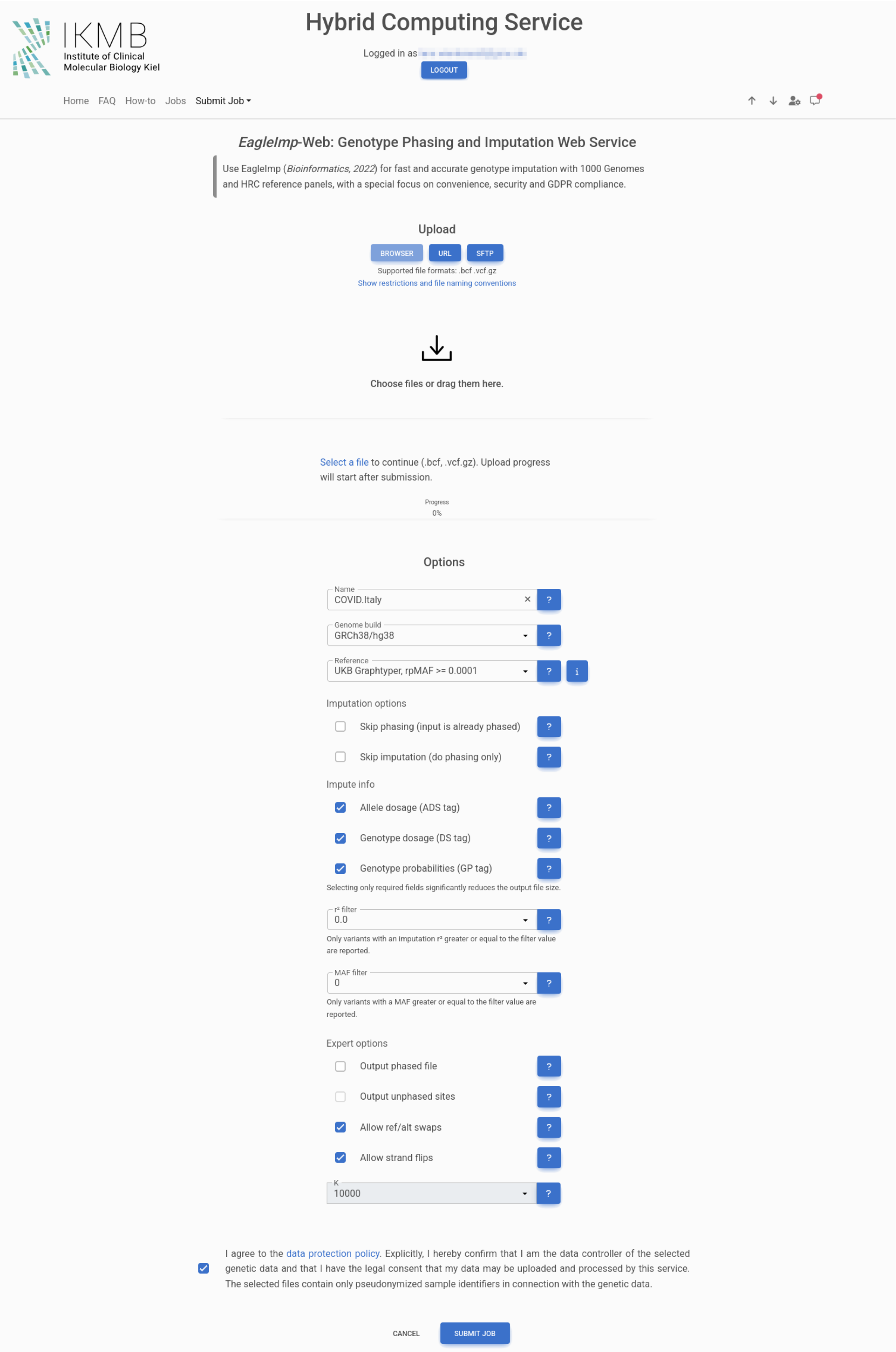

Supplementary Figure 3: Screenshot of *EagleImp-Web*'s job submission page with the available configuration options.

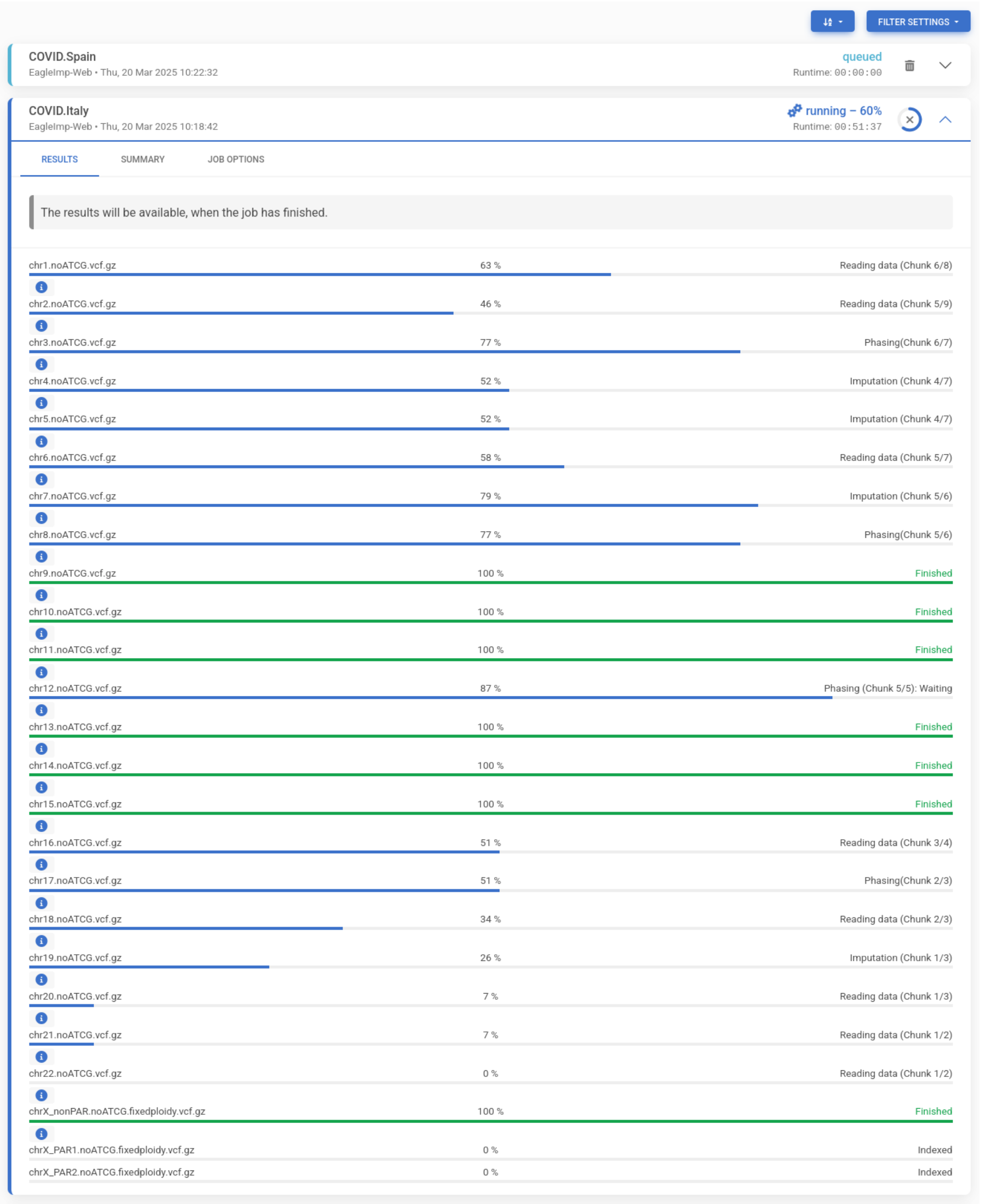

Supplementary Figure 4: Screenshot of an exemplary job progress in *EagleImp-Web*.

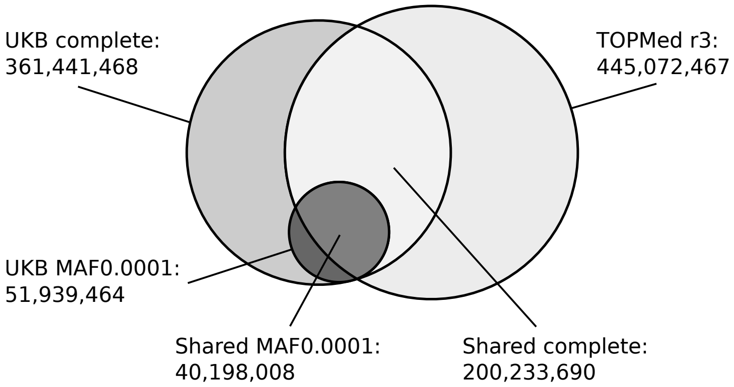

Supplementary Figure 5: Venn diagram comparing the number of variants in the UK Biobank reference panels (hg38 build) and the *TOPMed r3* reference panel. UKB complete shares 200,233,690 variants with *TOPMed* while UKB MAF0.0001 shares 40,198,008 variants.

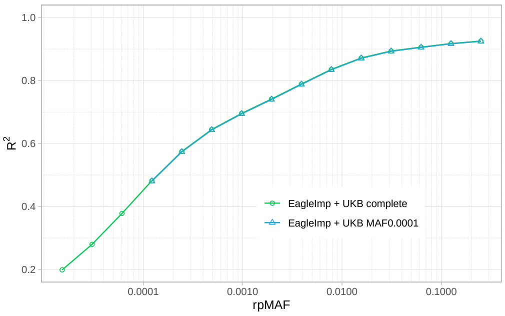

(a) AFR

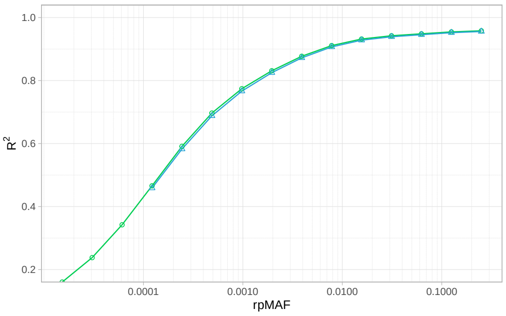

(b) AMR

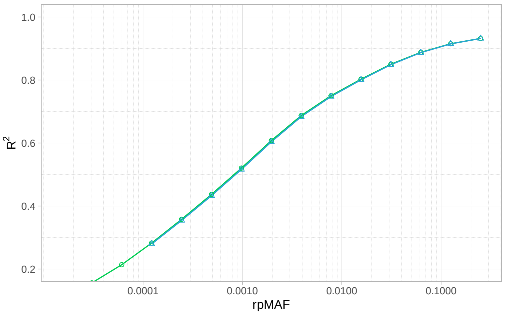

(c) EAS

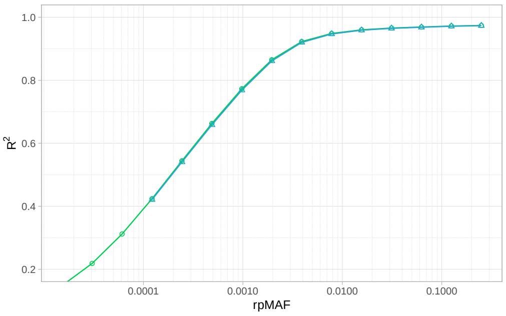

(d) EUR

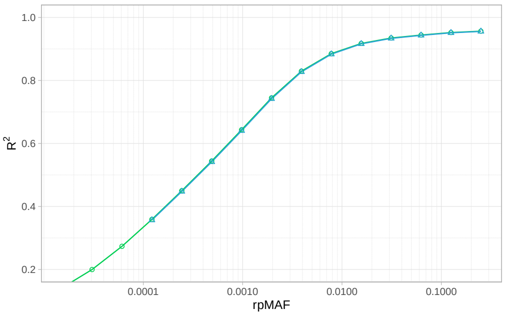

(e) SAS

Supplementary Figure 6: Estimated $R^2$ distribution for *EagleImp* + UKB complete panel and *EagleImp* + UKB MAF0.0001 panel for the benchmark datasets from the five superpopulations of the 1000 Genomes Project data. The results demonstrate no significant difference in mean $R^2$-values for both reference panels. Note that for the UKB MAF0.0001 panel, variants with an rpMAF $< 0.0001$ are not imputed.

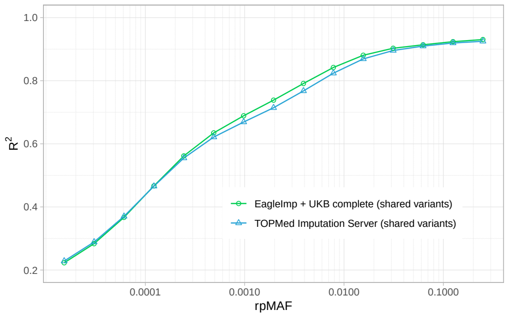

(a) AFR

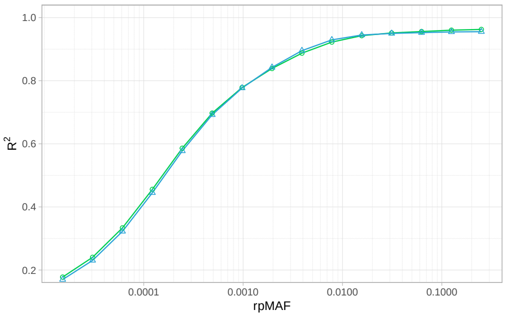

(b) AMR

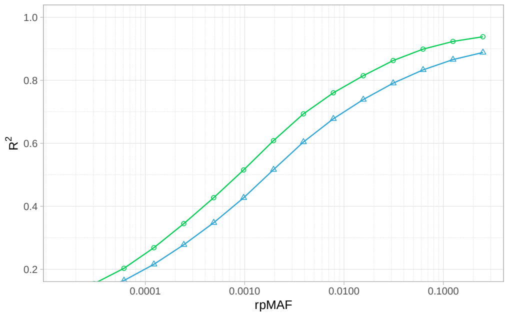

(c) EAS

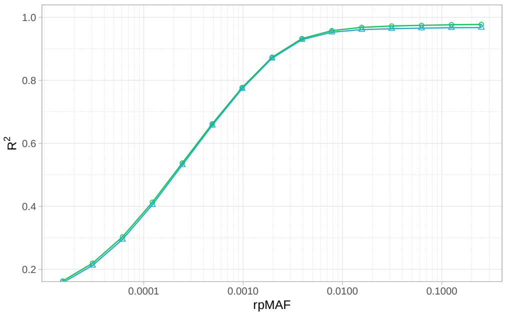

(d) EUR

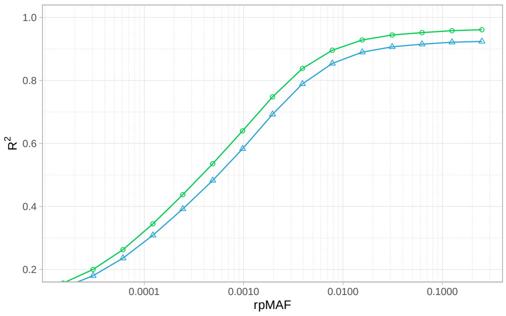

(e) SAS

Supplementary Figure 7: Estimated $R^2$ distribution for *EagleImp* + UKB complete panel in comparison to *TOPMed* for the benchmark datasets from the five superpopulations of the 1000 Genomes Project data. Only variants shared between the UKB panel and *TOPMed* were considered. Reference panel MAF (rpMAF) was taken from the UKB panel.

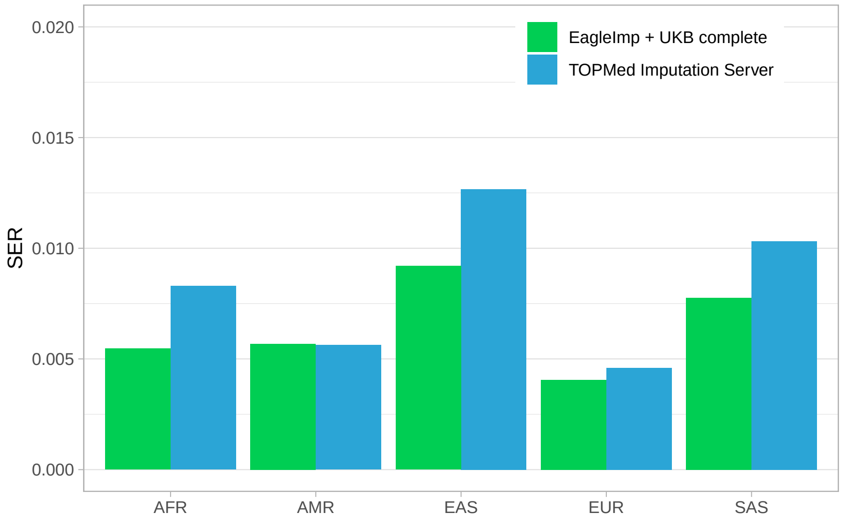

Supplementary Figure 8: Overview of the switch error rate (SER) for the five superpopulation benchmark datasets from the 1000 Genomes Project imputed with *EagleImp* and the complete UK Biobank reference panel (UKB complete) in comparison to *TOPMed*. SER is measured to the 1000 Genome reference (ground truth) and is based on all typed (input) variants shared with the *TOPMed r3* reference and the 1000 Genomes reference. The number of shared variants is 540,537.

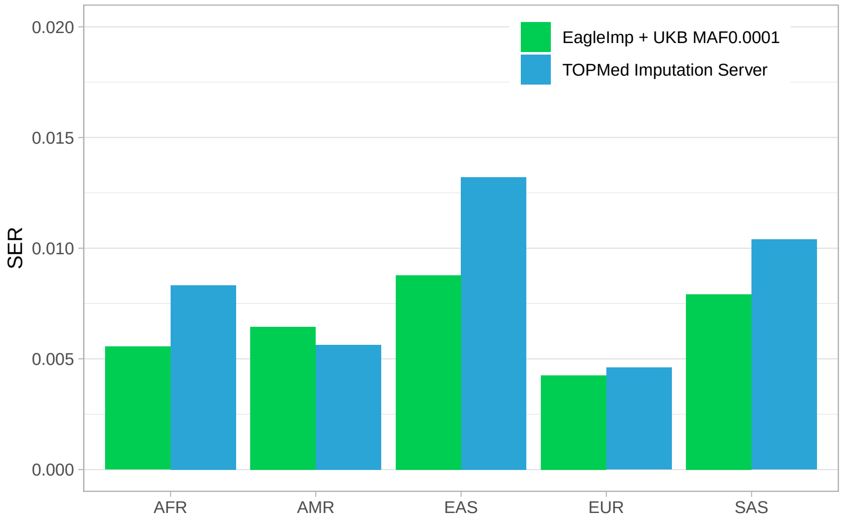

Supplementary Figure 9: Overview of the switch error rate (SER) for the five superpopulation benchmark datasets from the 1000 Genomes Project imputed with *EagleImp* and the rpMAF-filtered UK Biobank reference panel (UKB MAF0.0001) in comparison to *TOPMed*. SER is measured to the 1000 Genome reference (ground truth) and is based on all typed (input) variants shared with the *TOPMed r3* reference and the 1000 Genomes reference. The number of shared variants is 540,537.

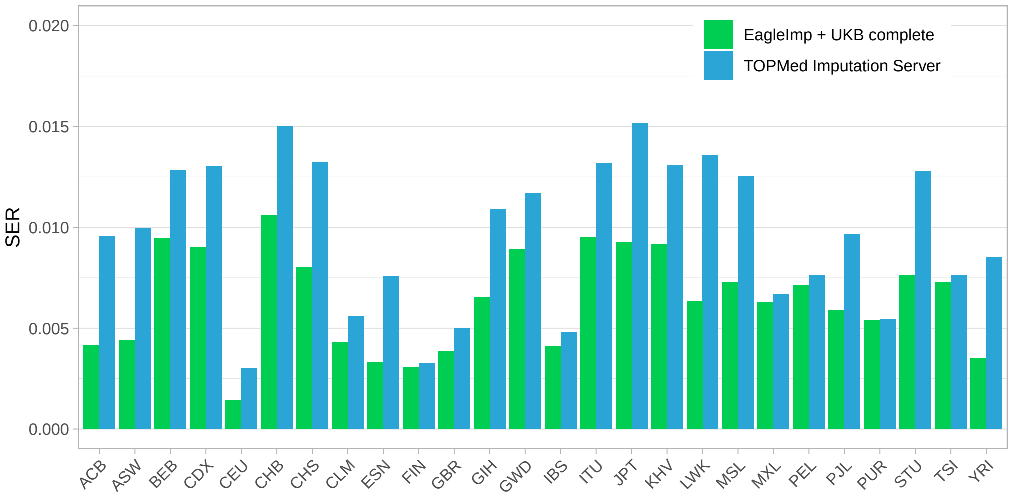

Supplementary Figure 10: Overview of the switch error rate (SER) for the 26 subpopulation benchmark datasets from the 1000 Genomes Project imputed with *EagleImp* and the complete UK Biobank reference panel (UKB complete) in comparison to *TOPMed*. SER is measured to the 1000 Genome reference (ground truth) and is based on all typed (input) variants shared with the *TOPMed r3* reference and the 1000 Genomes reference. The number of shared variants is 540,537.

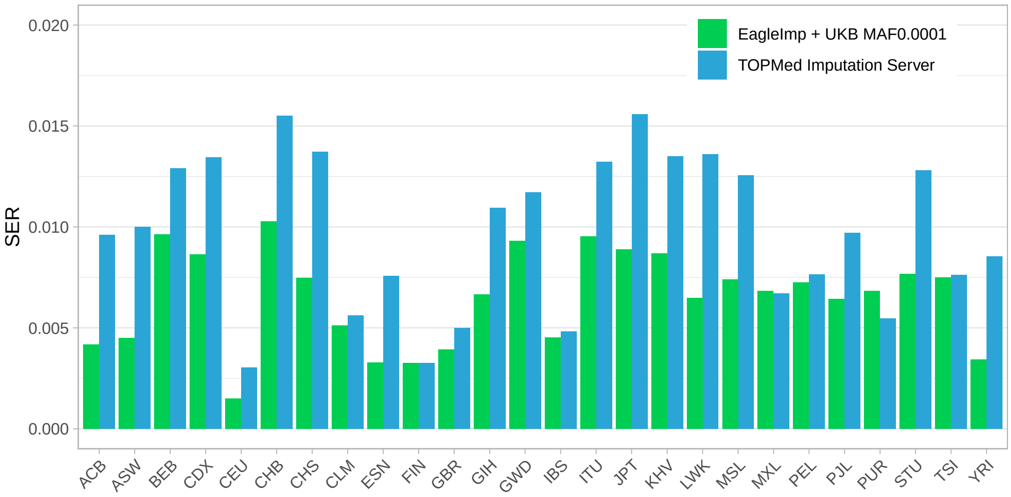

Supplementary Figure 11: Overview of the switch error rate (SER) for the 26 subpopulation benchmark datasets from the 1000 Genomes Project imputed with *EagleImp* and the rpMAF-filtered UK Biobank reference panel (UKB MAF0.0001) in comparison to *TOPMed*. SER is measured to the 1000 Genome reference (ground truth) and is based on all typed (input) variants shared with the *TOPMed r3* reference and the 1000 Genomes reference. The number of shared variants is 540,537.

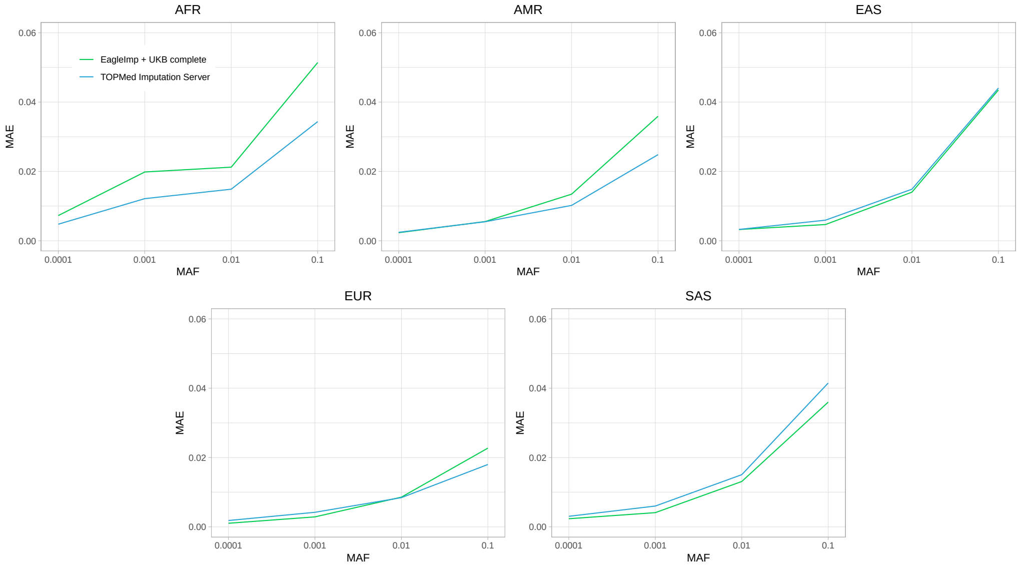

Supplementary Figure 12: Mean absolute error (MAE) imputation quality results for the five super-populations from the 1000 Genomes Project based on the 1000 Genomes reference panel ground truth, stratified by four rpMAF range categories and imputed by *EagleImp* with the complete UK Biobank reference panel (UKB complete) (green lines) and the *TOPMed Imputation Server* (*TOPMed r3* panel) (blue lines). MAE calculation is based on 56,908,240 variants shared between the UKB complete, the *TOPMed r3* panel and the 1000 Genomes Project panel.

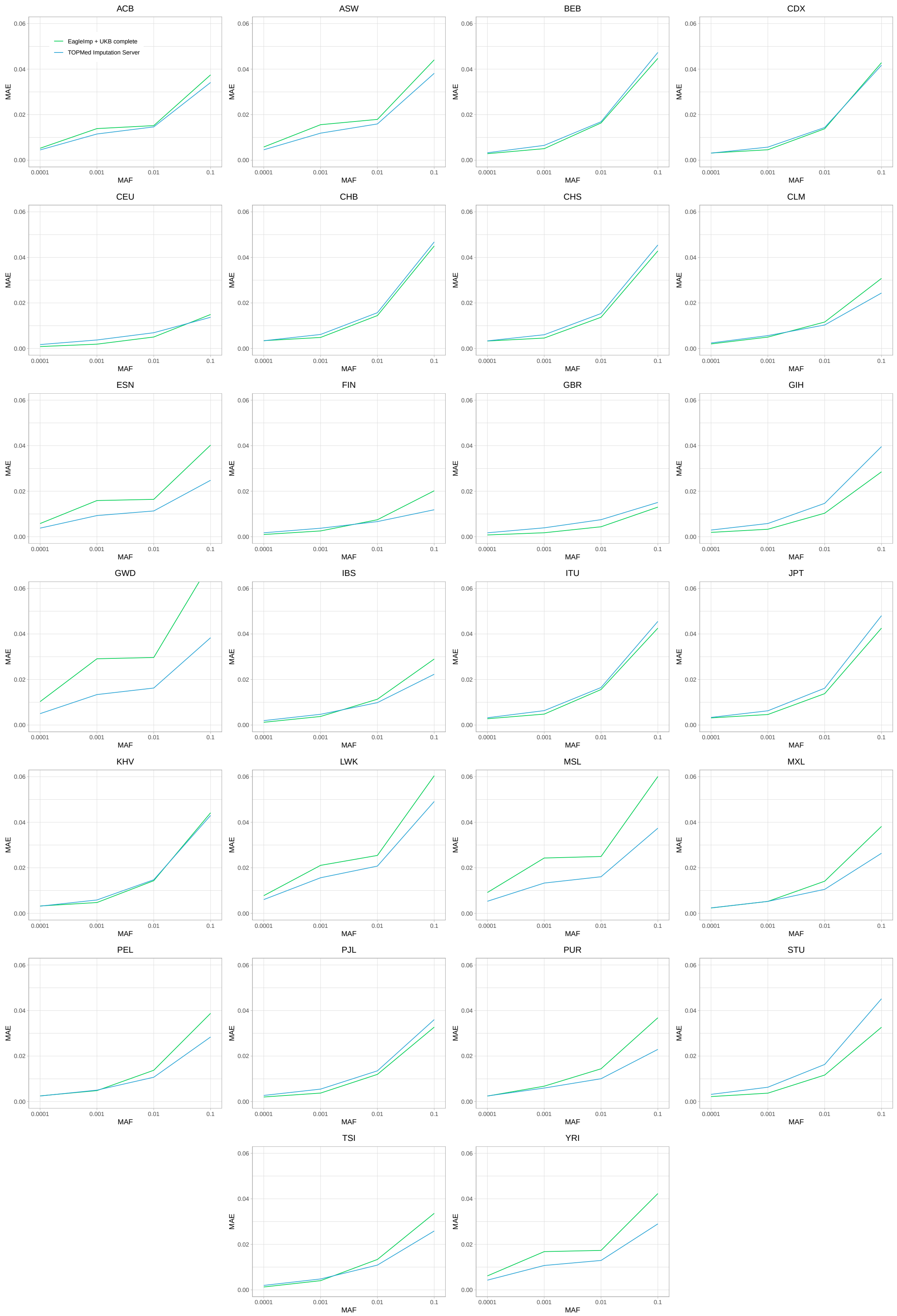

Supplementary Figure 13: Mean absolute error (MAE) imputation quality results for the 26 sub-populations from the 1000 Genomes Project based on the 1000 Genomes reference panel ground truth, stratified by four rpMAF range categories and imputed by *EagleImp* with the complete UK Biobank reference panel (UKB complete) (green lines) and the *TOPMed Imputation Server* (*TOPMed r3* panel) (blue lines). MAE calculation is based on 56,908,240 variants shared between the UKB complete, the *TOPMed r3* panel and the 1000 Genomes Project panel.

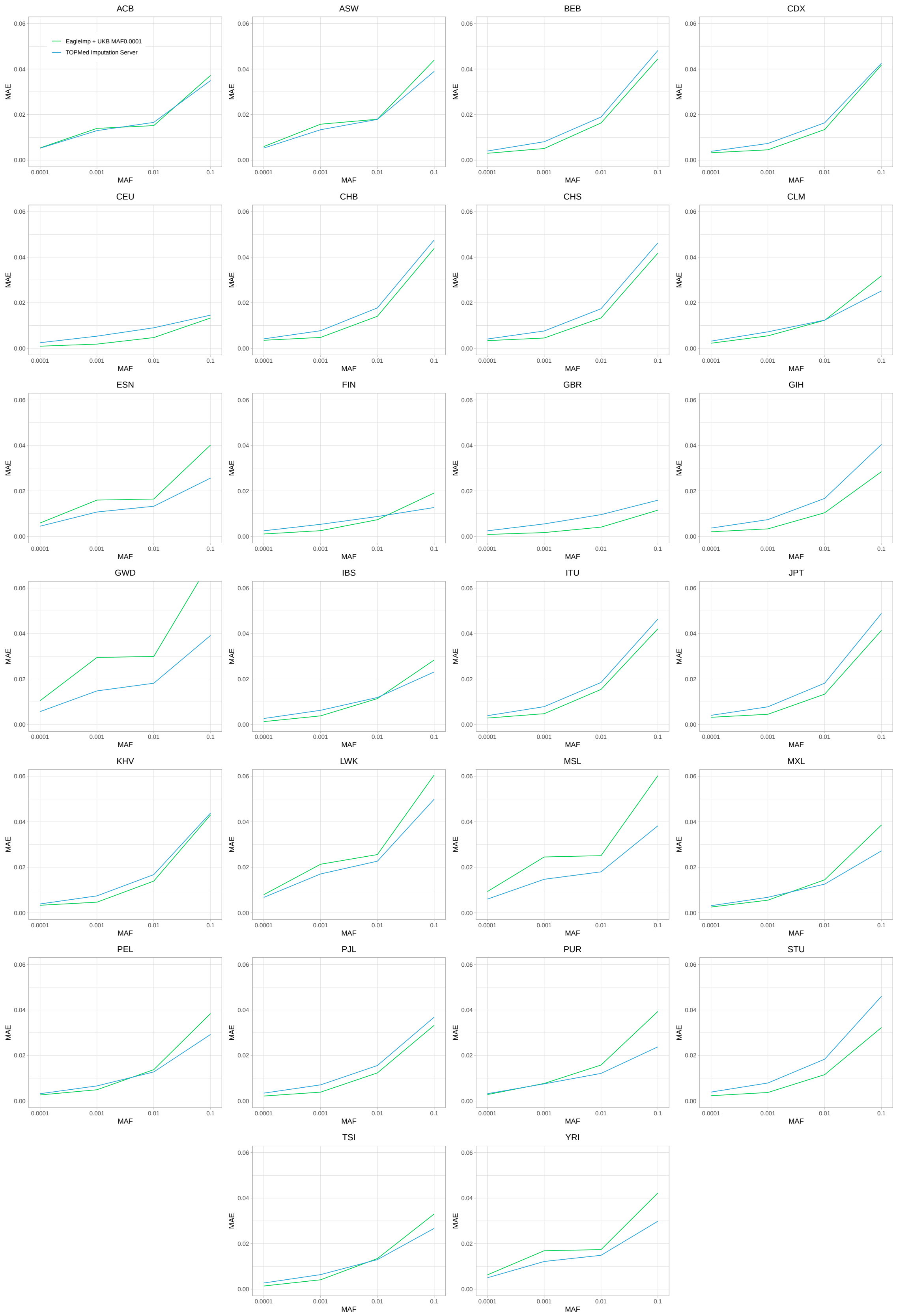

Supplementary Figure 14: Mean absolute error (MAE) imputation quality results for the 26 sub-populations from the 1000 Genomes Project based on the 1000 Genomes reference panel ground truth, stratified by four rpMAF range categories and imputed by *EagleImp* with the rpMAF-filtered UK Biobank reference panel (UKB MAF0.0001) (green lines) and the *TOPMed Imputation Server* (*TOPMed r3* panel) (blue lines). MAE calculation is based on 30,098,506 variants shared between the UKB MAF0.0001, the *TOPMed r3* panel and the 1000 Genomes Project panel.

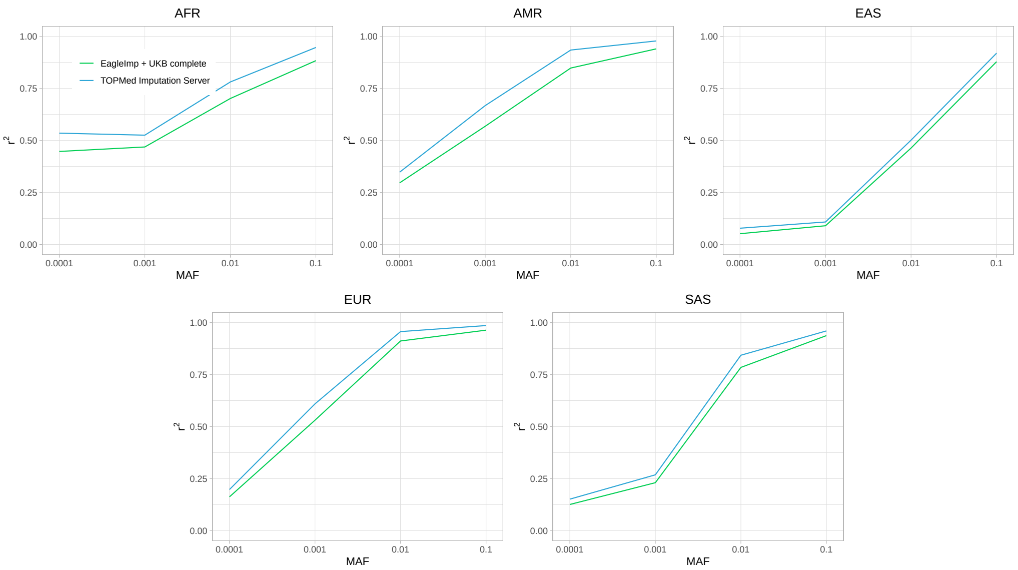

Supplementary Figure 15: Correlation $r^2$ stratified by four rpMAF range categories between the 1000 Genomes reference panel (ground truth) and the five superpopulation benchmark datasets from the 1000 Genomes Project imputed with *EagleImp* using the complete UK Biobank reference panel (UKB complete) and the *TOPMed Imputation Server*. Correlation $r^2$ was calculated separately for each variant. The final data points state the averages over all variants in the corresponding rpMAF category from the complete UK Biobank reference panel shared with *TOPMed r3* and 1000 Genomes. The number of shared variants is 56,908,240. Note, $r^2$ values which could not be calculated (e.g. due to missing variance) were set to $r^2 = 0$ which leads to a strong bias especially in the low rpMAF categories.

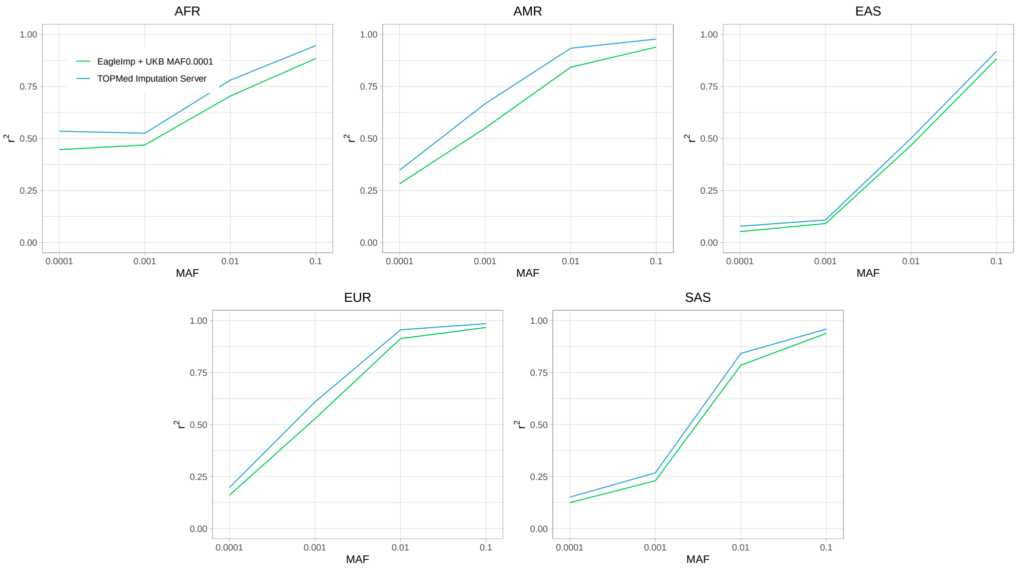

Supplementary Figure 16: Correlation $r^2$ stratified by four rpMAF range categories between the 1000 Genomes reference panel (ground truth) and the five superpopulation benchmark datasets from the 1000 Genomes Project imputed with *EagleImp* using the rpMAF-filtered UK Biobank reference panel (UKB MAF0.0001) and the *TOPMed Imputation Server*. Correlation $r^2$ was calculated separately for each variant. The final data points state the averages over all variants in the corresponding rpMAF category from the rpMAF-filtered UK Biobank reference panel shared with *TOPMed r3* and 1000 Genomes. The number of shared variants is 30,098,506. Note, $r^2$ values which could not be calculated (e.g. due to missing variance) were set to $r^2 = 0$ which leads to a strong bias especially in the low rpMAF categories.

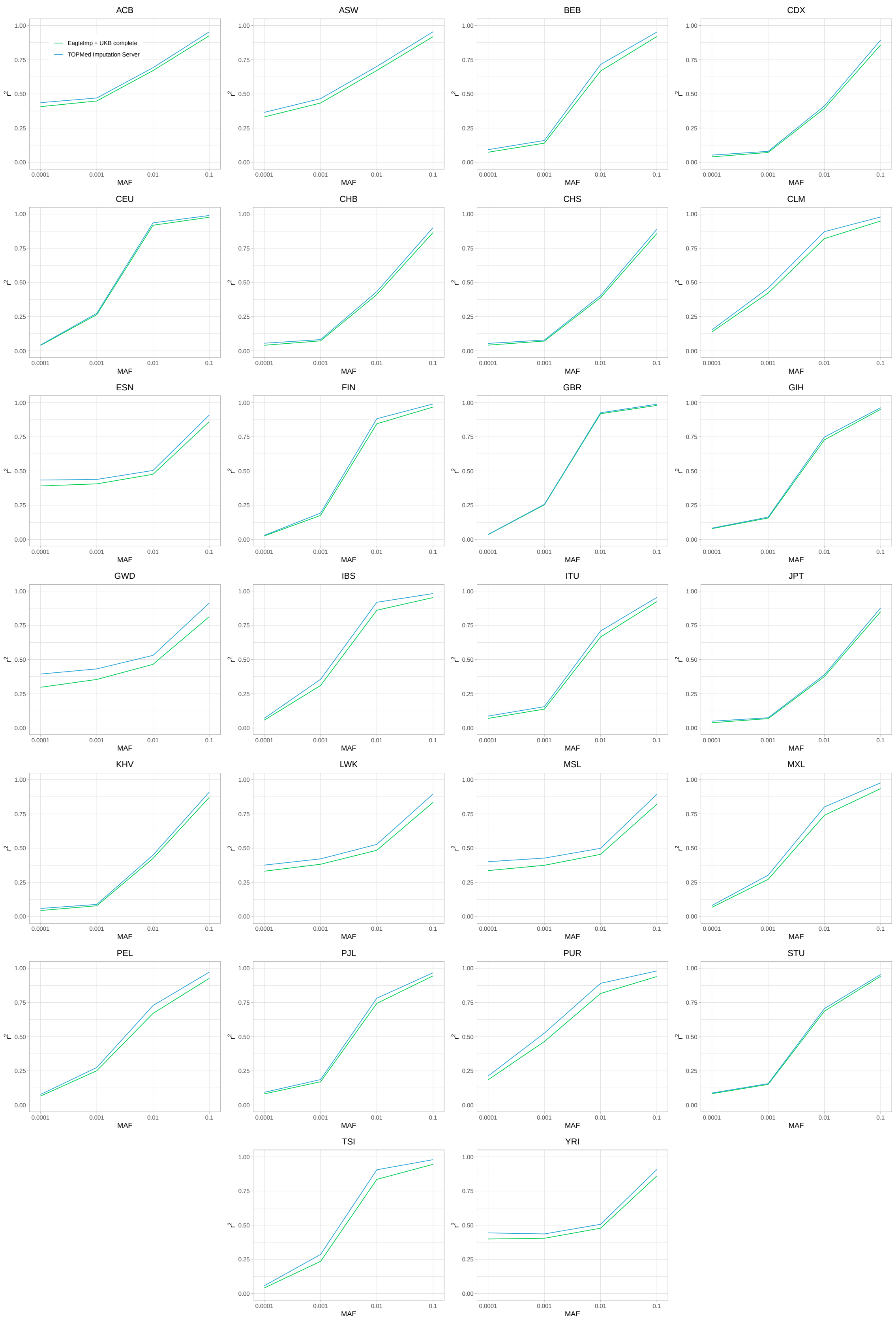

Supplementary Figure 17: Correlation $r^2$ stratified by four rpMAF range categories between the 1000 Genomes reference panel (ground truth) and the 26 subpopulation benchmark datasets from the 1000 Genomes Project imputed with *EagleImp* using the complete UK Biobank reference panel (UKB complete) and the *TOPMed Imputation Server*. Correlation $r^2$ was calculated separately for each variant. The final data points state the averages over all variants in the corresponding rpMAF category from the complete UK Biobank reference panel shared with *TOPMed r3* and 1000 Genomes. The number of shared variants is 56,908,240. Note, $r^2$ values which could not be calculated (e.g. due to missing variance) were set to $r^2 = 0$ which leads to a strong bias especially in the low rpMAF categories.

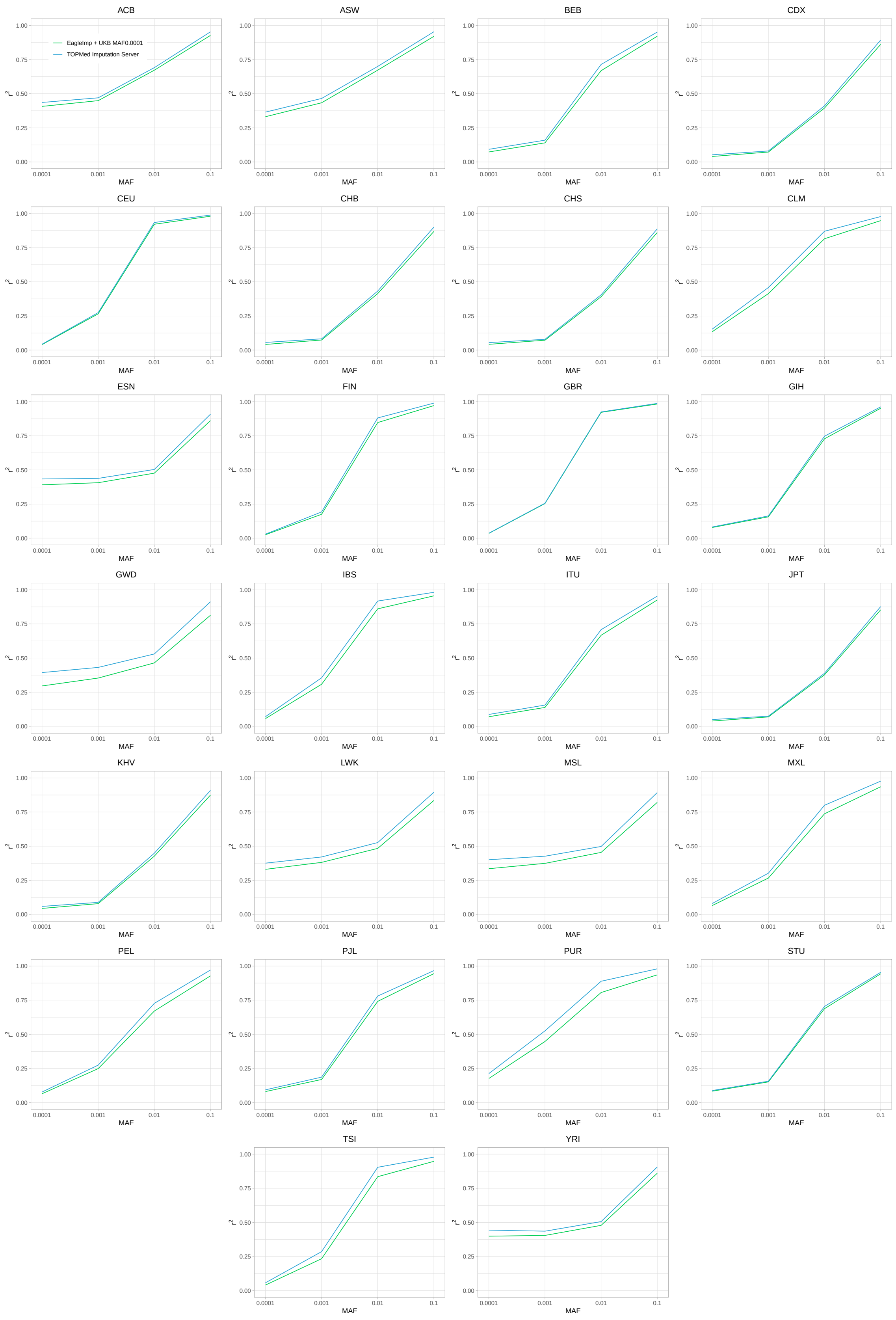

Supplementary Figure 18: Correlation $r^2$ stratified by four rpMAF range categories between the 1000 Genomes reference panel (ground truth) and the 26 subpopulation benchmark datasets from the 1000 Genomes Project imputed with *EagleImp* using the rpMAF-filtered UK Biobank reference panel (UKB MAF0.0001) and the *TOPMed Imputation Server*. Correlation $r^2$ was calculated separately for each variant. The final data points state the averages over all variants in the corresponding rpMAF category from the rpMAF-filtered UK Biobank reference panel shared with *TOPMed r3* and 1000 Genomes. The number of shared variants is 30,098,506. Note, $r^2$ values which could not be calculated (e.g. due to missing variance) were set to $r^2 = 0$ which leads to a strong bias especially in the low rpMAF categories.

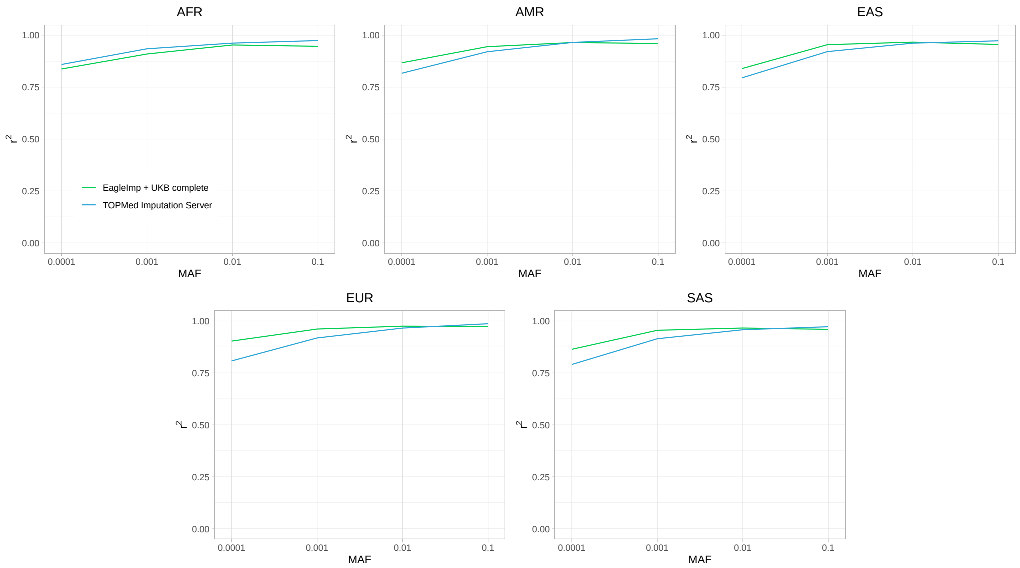

Supplementary Figure 19: Correlation $r^2$ between the 1000 Genomes reference panel (ground truth) and the five benchmark datasets from the superpopulations in the 1000 Genomes Project imputed with *EagleImp* using the complete UK Biobank reference panel (UKB complete) and the *TOPMed Imputation Server*. Correlation $r^2$ was calculated for the complete dataset over all data points from all variants and samples (but separately for each rpMAF category in each chromosome file, but not separately for each variant). Computation is based on 56,908,240 shared variants in the complete UK Biobank reference panel, the *TOPMed r3* panel and the 1000 Genomes reference panel.

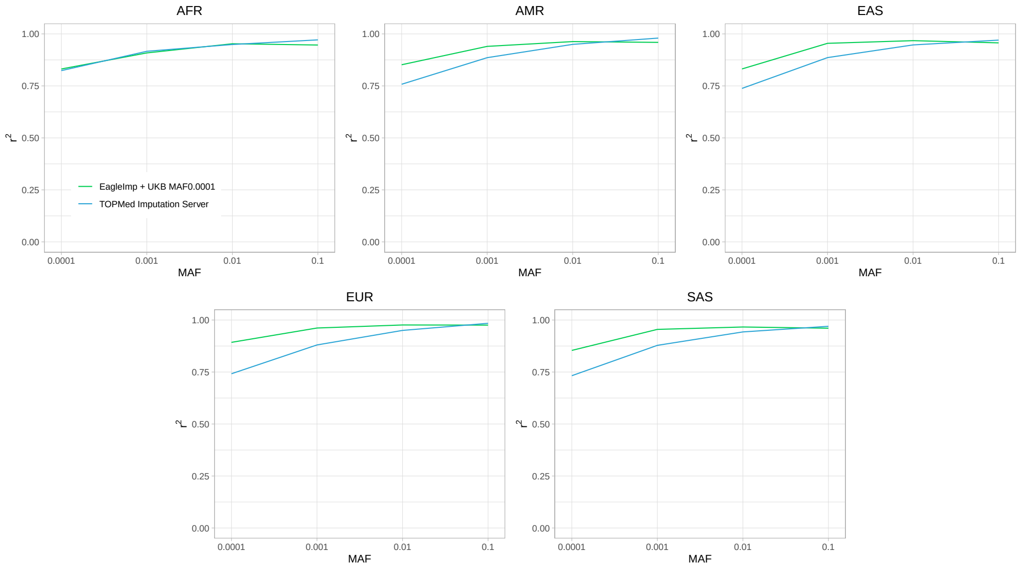

Supplementary Figure 20: Correlation $r^2$ between the 1000 Genomes reference panel (ground truth) and the five benchmark datasets from the superpopulations in the 1000 Genomes Project imputed with *EagleImp* using the rpMAF-filtered UK Biobank reference panel (UKB MAF0.0001) and the *TOPMed Imputation Server*. Correlation $r^2$ was calculated for the complete dataset over all data points from all variants and samples (but separately for each rpMAF category in each chromosome file, but not separately for each variant). Computation is based on 30,098,506 shared variants in the rpMAF-filtered UK Biobank reference panel, the *TOPMed r3* panel and the 1000 Genomes reference panel.

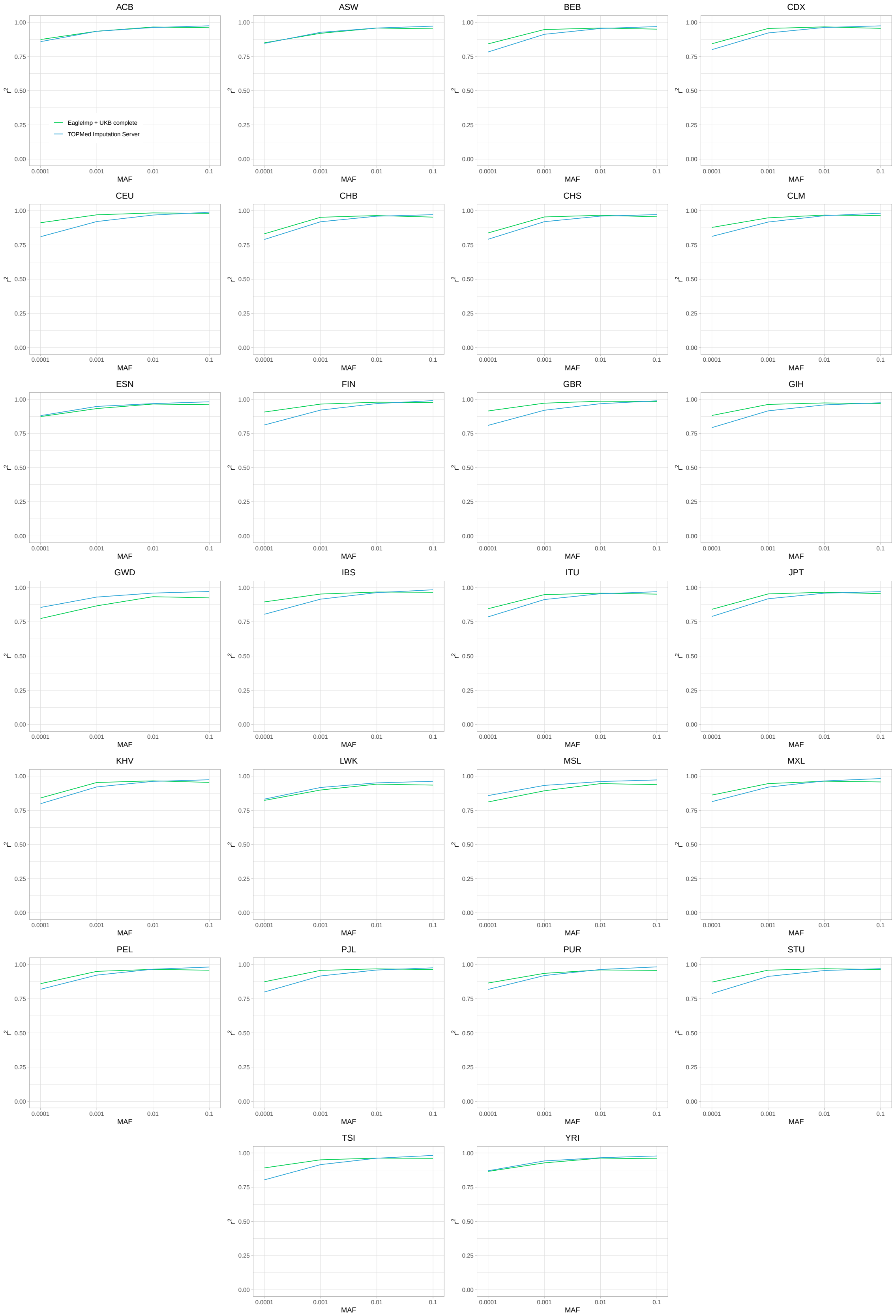

Supplementary Figure 21: Correlation $r^2$ between the 1000 Genomes reference panel (ground truth) and the 26 benchmark datasets from the subpopulations in the 1000 Genomes Project imputed with *EagleImp* using the complete UK Biobank reference panel (UKB complete) and the *TOPMed Imputation Server*. Correlation $r^2$ was calculated for the complete dataset over all data points from all variants and samples (but separately for each rpMAF category in each chromosome file, but not separately for each variant). Computation is based on 56,908,240 shared variants in the complete UK Biobank reference panel, the *TOPMed r3* panel and the 1000 Genomes reference panel.

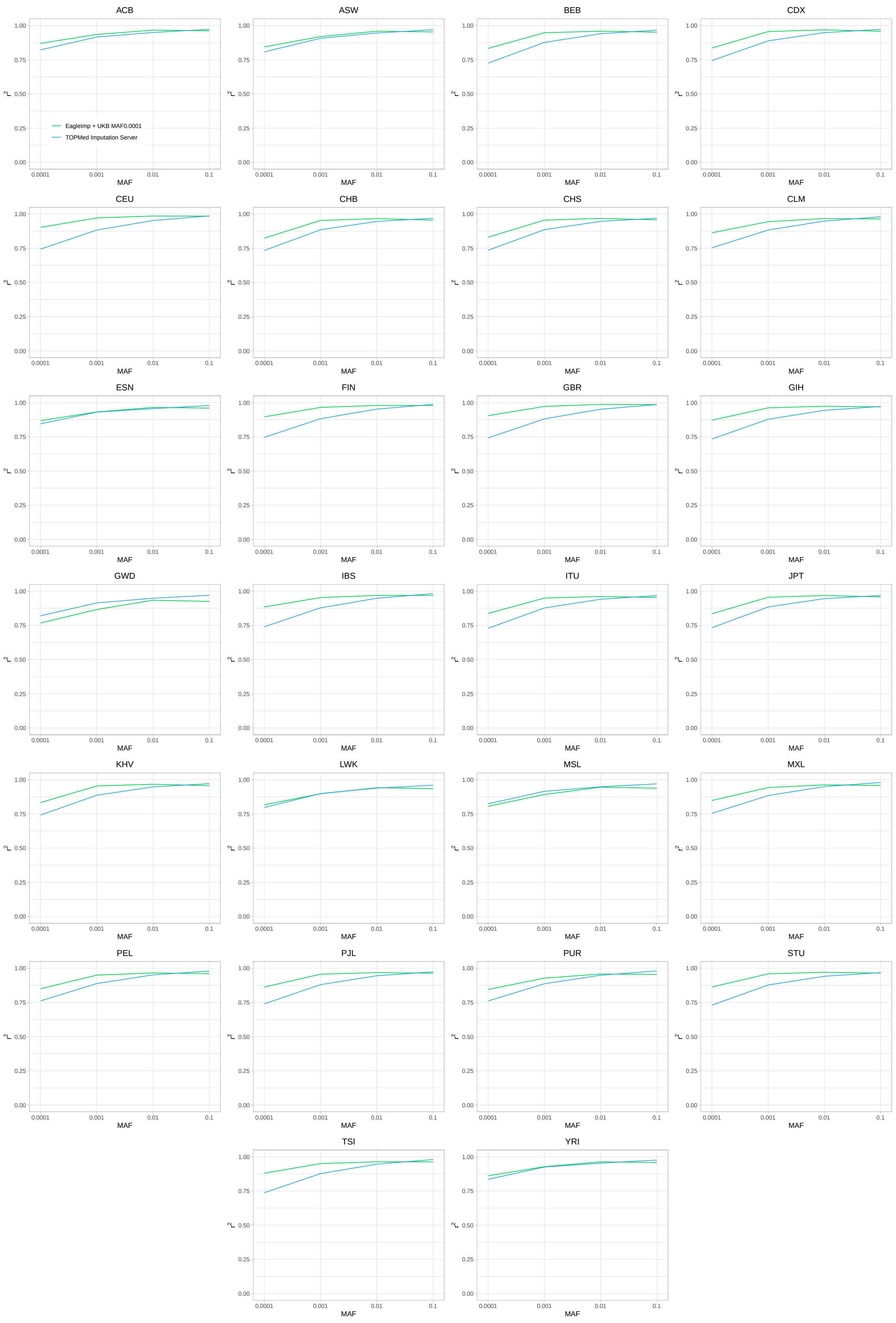

Supplementary Figure 22: Correlation $r^2$ between the 1000 Genomes reference panel (ground truth) and the 26 benchmark datasets from the subpopulations in the 1000 Genomes Project imputed with *EagleImp* using the rpMAF-filtered UK Biobank reference panel (UKB MAF0.0001) and the *TOPMed Imputation Server*. Correlation $r^2$ was calculated for the complete dataset over all data points from all variants and samples (but separately for each rpMAF category in each chromosome file, but not separately for each variant). Computation is based on 30,098,506 shared variants in the rpMAF-filtered UK Biobank reference panel, the *TOPMed r3* panel and the 1000 Genomes reference panel.

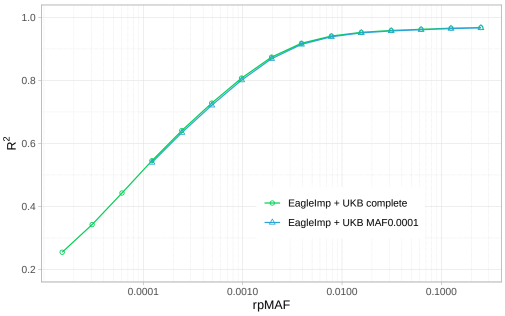

(a) Germany

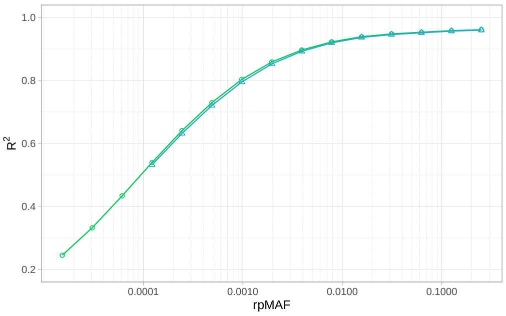

(b) Italy

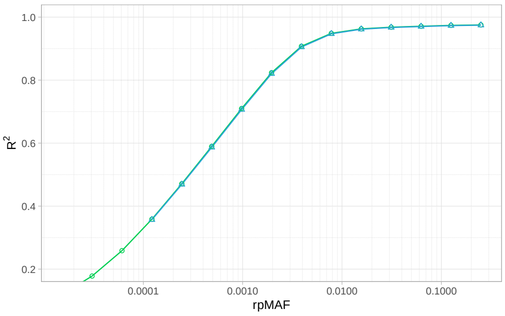

(c) Norway

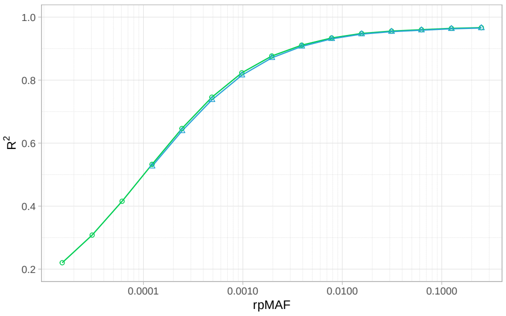

(d) Spain

Supplementary Figure 23: Estimated $R^2$ distribution for *EagleImp* + UKB complete panel and *EagleImp* + UKB MAF0.0001 panel for the COVID-19 GWAS benchmark datasets. The results demonstrate no significant difference in mean $R^2$-values for both reference panels. Note that for the UKB MAF0.0001 panel, variants with an rpMAF $< 0.0001$ are not imputed.

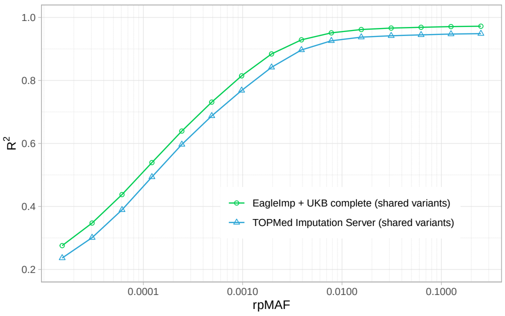

(a) Germany

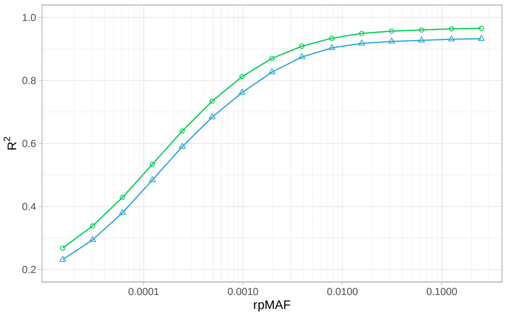

(b) Italy

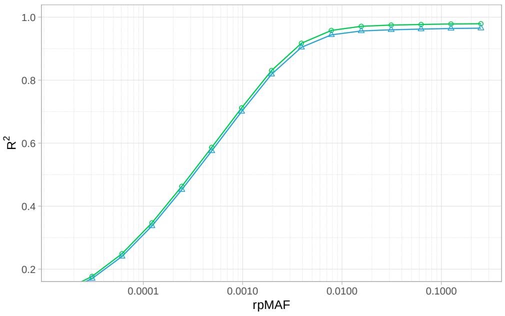

(c) Norway

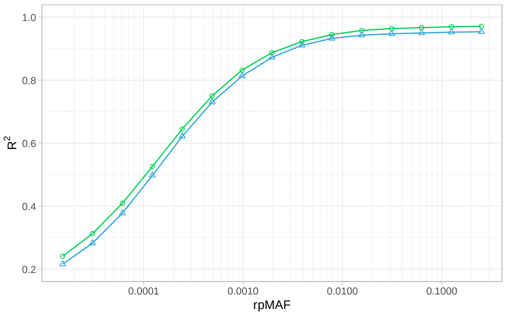

(d) Spain

Supplementary Figure 24: Estimated $R^2$ distribution for *EagleImp* + UKB complete panel in comparison to *TOPMed* for the COVID-19 GWAS benchmark datasets. Only variants shared between the UKB panel and *TOPMed* were considered. Reference panel MAF (rpMAF) was taken from the UKB panel.

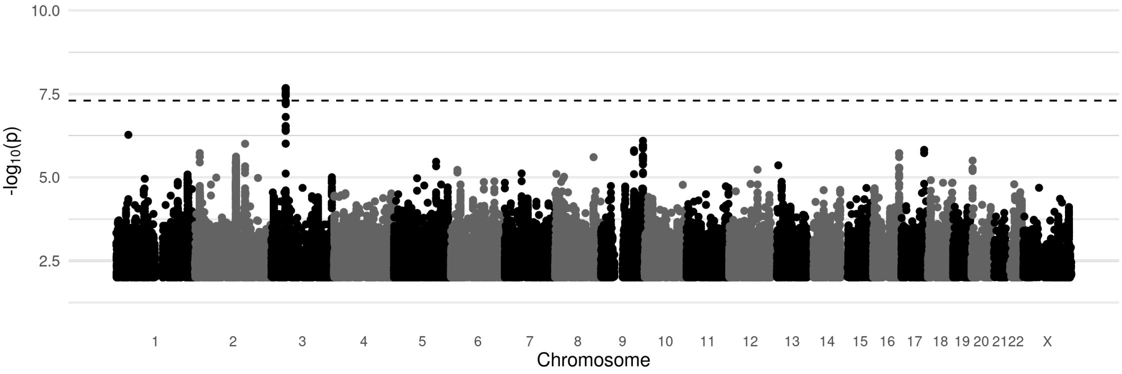

(a) *EagleImp* with UKB MAF0.0001 panel

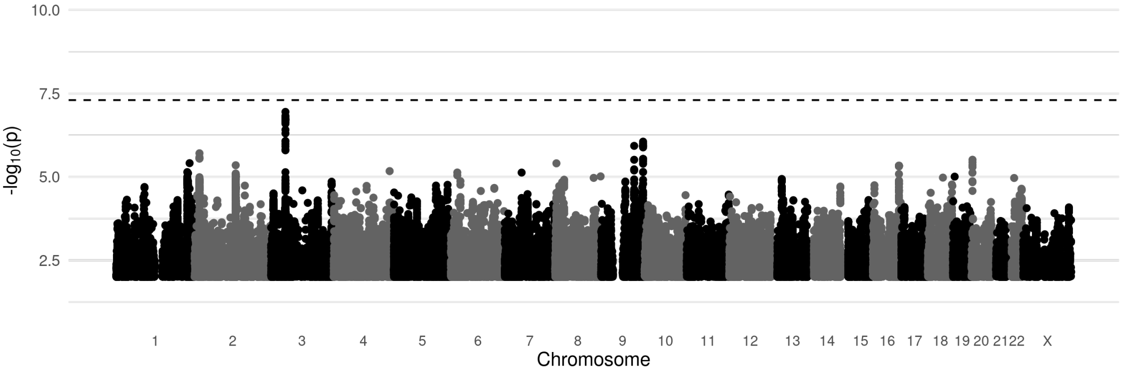

(b) *TOPMed Imputation Server*

Supplementary Figure 25: Manhattan plots of COVID-19 GWAS using 835 Italian cases with severe respiratory failures and 1,255 Italian controls [14]. Imputation was processed (a) using *EagleImp* and the new UKB panel with applied refpanel MAF $\geq 0.0001$ filter, and (b) using the *TOPMed Imputation Server*. All imputation results were filtered by an estimated imputation $R^2$ threshold of 0.6 and a MAF threshold of 0.01. Association analysis was performed using the *BIGwas* pipeline [20] and association test statistics from PLINK2 [21] including 10 principal components from the principal component analysis (PCA) to control for potential population stratification. The dotted line represents the genome-wide significance threshold at $p = 5e$-8. For the plots all results with a $p$-value $> 0.01$ were filtered.

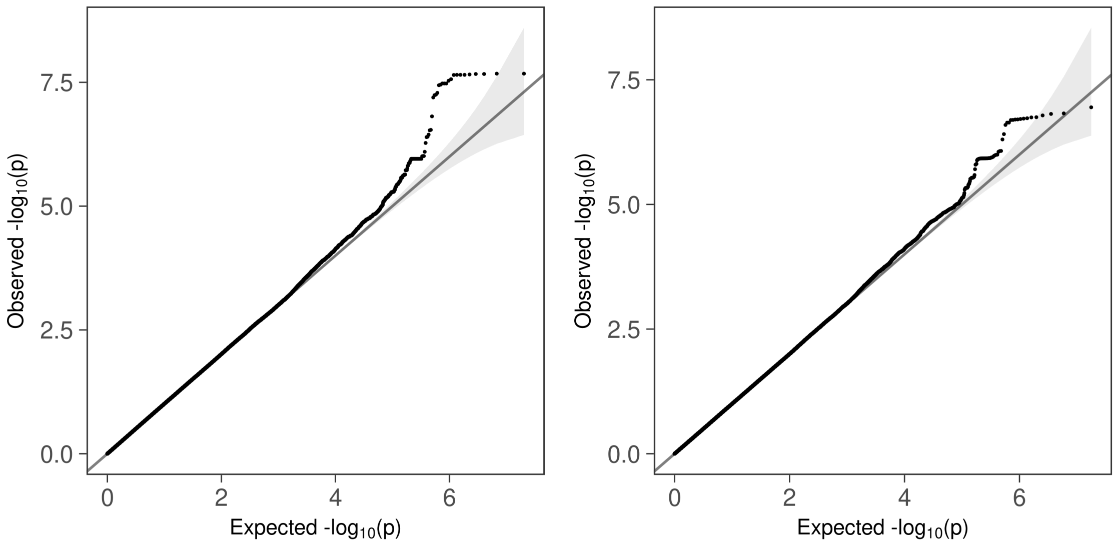

(a) *EagleImp* with UKB MAF0.0001 panel

(b) *TOPMed Imputation Server*

Supplementary Figure 26: QQ plots of expected and observed association $p$-values in a COVID-19 GWAS using 835 Italian cases with severe respiratory failures and 1,255 Italian controls [14]. Imputation was processed (a) using *EagleImp* and the new UKB panel with applied refpanel MAF $\geq 0.0001$ filter, and (b) using the *TOPMed Imputation Server*. All imputation results were filtered by an estimated imputation $R^2$ threshold of 0.6 and a MAF threshold of 0.01. Association analysis was performed using the *BIGwas* pipeline [20] and association test statistics from PLINK2 [21] including 10 principal components from the principal component analysis (PCA) to control for potential population stratification.

### 7 Supplementary Tables

Supplementary Table 1: Features implemented in *EagleImp-Web*.

| |
|---|
| Free and simple account registration:<br>• Only email address and password required<br>• Download of personal account information<br>• Delete account option |
| Data privacy and security:<br>• Data protection policy on website<br>• Optional 2-factor authentication (2FA) via passkeys (for smartphone, iCloud KeyChain, USB key dongle, etc.)<br>• Certified encrypted connection<br>• No 3rd party data sharing or data transfer to commercial cloud or other external systems<br>• No advertisements, no tracking, no cookies (a necessary session cookie is stored as long as the user is logged in) |
| Imputation of `.bcf` and `.vcf.gz` input files |
| Data integrity check already in web browser from client computer before any data upload |
| Reference panels from HRC and 1000 Genomes Project |
| rpMAF-filtered reference panel available (faster imputation when very low MAF variants are not required) |
| *EagleImp* process options available, e.g.:<br>• Skip phasing (if input is already phased)<br>• Skip imputation (if only phasing is desired)<br>• Freely selectable VCF output information (allele dosage, genotype dosage and/or genotype probabilities)<br>• Choice of $K$-parameter for phasing<br>• Choice of $R^2$ filter<br>• Choice of MAF filter<br>• Additional output file for phased input<br>• Choice of input filters to allow for ref/alt swaps and/or strand flips |
| Job control dashboard:<br>• Live online monitoring of job progress<br>• Background processing (no need to stay logged in)<br>• Notification email for finished jobs<br>• All output files (including result and log files) are made available for download, even when the job is cancelled or fails. |
| Download via browser or terminal:<br>• Secure download with access protection and encrypted connection (no need for extra encryption of data files) |
| Message board (similar to that in the UK Biobank AMS Portal) |

Supplementary Table 2: Populations in the UKB panel as reported in Extended Data Table 3 of [22] (participants self-reported their ethnic background in a questionnaire). 2,131 additional individuals present in the Graphtyper data and missing in the table were added to the Other/Unknown group.

| **Population group**<br>(Self reported ethnic group) | **No. of individuals** | **%** |
|---|---:|---:|
| White | 460,186 | 93.85% |
| Asian or Asian British | 9,474 | 1.93% |
| Black or Black British | 7,649 | 1.56% |
| Chinese | 1,504 | 0.31% |
| Mixed | 2,843 | 0.58% |
| Other/Unknown | 8,663 | 1.77% |

Supplementary Table 3: Benchmark datasets used for imputation with the UKB reference panels and for quality comparison with the *TOPMed Imputation Server*. Datasets from *1000 Genomes Project* consist of extracted Illumina GSA variants. The subpopulation datasets are subsets of the corresponding superpopulation (extra column with corresponding superpopulation code). Datasets from COVID-19 studies are GSA-typed and variants were *quality control (QC)* filtered using the *BIGwas* QC pipeline [20].

| **Name** | **Description** | | **#variants** | **#samples** |
|---|---|---|---|---|
| *Datasets from 1000 Genomes [6] superpopulations:* | | | | |
| AFR | African ancestry | | 619,587 | 661 |
| AMR | American ancestry | | 619,587 | 347 |
| EAS | East Asian ancestry | | 619,587 | 504 |
| EUR | European ancestry | | 619,587 | 494 |
| SAS | South Asian ancestry | | 619,587 | 489 |
| *Datasets from 1000 Genomes [6] subpopulations:* | | | | |
| ACB | African Caribbean in Barbados | AFR | 619,587 | 96 |
| ASW | African Ancestry in Southwest US | AFR | 619,587 | 61 |
| BEB | Bengali in Bangladesh | SAS | 619,587 | 86 |
| CDX | Chinese Dai in Xishuangbanna, China | EAS | 619,587 | 93 |
| CEU | Utah residents with European ancestry | EUR | 619,587 | 90 |
| CHB | Han Chinese in Beijing, China | EAS | 619,587 | 103 |
| CHS | Han Chinese South | EAS | 619,587 | 105 |
| CLM | Colombian in Medellin, Colombia | AMR | 619,587 | 94 |
| ESN | Esan in Nigeria | AFR | 619,587 | 99 |
| FIN | Finnish in Finland | EUR | 619,587 | 99 |
| GBR | British in England and Scotland | EUR | 619,587 | 91 |
| GIH | Gujarati Indians in Houston, TX | SAS | 619,587 | 103 |
| GWD | Gambian in Western Division, The Gambia | AFR | 619,587 | 113 |
| IBS | Iberian populations in Spain | EUR | 619,587 | 107 |
| ITU | Indian Telugu in the UK | SAS | 619,587 | 102 |
| JPT | Japanese in Tokyo, Japan | EAS | 619,587 | 104 |
| KHV | Kinh in Ho Chi Minh City, Vietnam | EAS | 619,587 | 99 |
| LWK | Luhya in Webuye, Kenya | AFR | 619,587 | 99 |
| MSL | Mende in Sierra Leone | AFR | 619,587 | 85 |
| MXL | Mexican Ancestry in Los Angeles, California | AMR | 619,587 | 64 |
| PEL | Peruvian in Lima, Peru | AMR | 619,587 | 85 |
| PJL | Punjabi in Lahore, Pakistan | SAS | 619,587 | 96 |
| PUR | Puerto Rican in Puerto Rico | AMR | 619,587 | 104 |
| STU | Sri Lankan Tamil in the UK | SAS | 619,587 | 102 |
| TSI | Toscani in Italy | EUR | 619,587 | 107 |
| YRI | Yoruba in Ibadan, Nigeria | AFR | 619,587 | 108 |
| *Datasets from COVID-19 studies [14, 15]:* | | | | |
| Germany | German incl. 241 COVID-19 cases with severe respiratory failure + 3,110 controls | | 476,770 | 3,351 |
| Italy | Italian incl. 835 COVID-19 cases with severe respiratory failure + 1,255 controls | | 559,519 | 2,090 |
| Norway | Norwegian incl. 62 COVID-19 cases with severe respiratory failure + 262 controls | | 525,216 | 324 |
| Spain | Spanish incl. 775 COVID-19 cases with severe respiratory failure + 950 controls | | 549,696 | 1,725 |

Supplementary Table 4: Mean estimated imputation $R^2$ values reported by *EagleImp* using the complete UKB reference panel (UKB complete) and the rpMAF-filtered UKB reference panel (UKB MAF0.0001) for all benchmark datasets classified by rpMAF.

| Dataset | **UKB complete** refpanel MAF ≥ | | | **UKB MAF0.0001** refpanel MAF ≥ | | | **Difference** refpanel MAF ≥ | | |
|---|---|---|---|---|---|---|---|---|---|
| | 0.0001 | 0.001 | 0.01 | 0.0001 | 0.001 | 0.01 | 0.0001 | 0.001 | 0.01 |
| *Datasets from 1000 Genomes [6] superpopulations:* | | | | | | | | | |
| AFR | 0.4821 | 0.6959 | 0.8356 | 0.4808 | 0.6943 | 0.8342 | 0.0013 | 0.0016 | 0.0014 |
| AMR | 0.4659 | 0.7742 | 0.9111 | 0.4588 | 0.7665 | 0.9068 | 0.0071 | 0.0077 | 0.0043 |
| EAS | 0.2825 | 0.5204 | 0.7505 | 0.2794 | 0.5160 | 0.7476 | 0.0031 | 0.0044 | 0.0029 |
| EUR | 0.4238 | 0.7730 | 0.9486 | 0.4209 | 0.7685 | 0.9468 | 0.0029 | 0.0045 | 0.0018 |
| SAS | 0.3589 | 0.6434 | 0.8851 | 0.3567 | 0.6404 | 0.8833 | 0.0022 | 0.0030 | 0.0018 |
| *Datasets from 1000 Genomes [6] subpopulations:* | | | | | | | | | |
| ACB | 0.4238 | 0.6496 | 0.8131 | 0.4235 | 0.6493 | 0.8127 | 0.0003 | 0.0003 | 0.0004 |
| ASW | 0.3952 | 0.6414 | 0.8107 | 0.3945 | 0.6405 | 0.8100 | 0.0007 | 0.0009 | 0.0007 |
| BEB | 0.2687 | 0.5372 | 0.8199 | 0.2674 | 0.5352 | 0.8182 | 0.0013 | 0.0020 | 0.0017 |
| CDX | 0.2141 | 0.4347 | 0.6835 | 0.2126 | 0.4320 | 0.6810 | 0.0015 | 0.0027 | 0.0025 |
| CEU | 0.2863 | 0.6226 | 0.9405 | 0.2857 | 0.6215 | 0.9401 | 0.0006 | 0.0011 | 0.0004 |
| CHB | 0.2211 | 0.4454 | 0.6965 | 0.2194 | 0.4425 | 0.6939 | 0.0017 | 0.0029 | 0.0026 |
| CHS | 0.2179 | 0.4381 | 0.6850 | 0.2161 | 0.4351 | 0.6823 | 0.0018 | 0.0030 | 0.0027 |
| CLM | 0.3487 | 0.6786 | 0.8917 | 0.3451 | 0.6734 | 0.8882 | 0.0036 | 0.0052 | 0.0035 |
| ESN | 0.3867 | 0.5768 | 0.7096 | 0.3863 | 0.5762 | 0.7088 | 0.0004 | 0.0006 | 0.0008 |
| FIN | 0.2644 | 0.5769 | 0.9015 | 0.2634 | 0.5748 | 0.8996 | 0.0010 | 0.0021 | 0.0019 |
| GBR | 0.2843 | 0.6214 | 0.9431 | 0.2838 | 0.6205 | 0.9428 | 0.0005 | 0.0009 | 0.0003 |
| GIH | 0.2731 | 0.5512 | 0.8493 | 0.2722 | 0.5499 | 0.8481 | 0.0009 | 0.0013 | 0.0012 |
| GWD | 0.3693 | 0.5704 | 0.7101 | 0.3677 | 0.5679 | 0.7072 | 0.0016 | 0.0025 | 0.0029 |
| IBS | 0.3066 | 0.6408 | 0.9124 | 0.3045 | 0.6372 | 0.9099 | 0.0021 | 0.0036 | 0.0025 |
| ITU | 0.2698 | 0.5392 | 0.8214 | 0.2687 | 0.5374 | 0.8200 | 0.0011 | 0.0018 | 0.0014 |
| JPT | 0.2134 | 0.4311 | 0.6759 | 0.2117 | 0.4281 | 0.6731 | 0.0017 | 0.0030 | 0.0028 |
| KHV | 0.2234 | 0.4509 | 0.7042 | 0.2218 | 0.4481 | 0.7017 | 0.0016 | 0.0028 | 0.0025 |
| LWK | 0.3855 | 0.5926 | 0.7336 | 0.3843 | 0.5905 | 0.7311 | 0.0012 | 0.0021 | 0.0025 |
| MSL | 0.3710 | 0.5647 | 0.6988 | 0.3701 | 0.5633 | 0.6971 | 0.0009 | 0.0014 | 0.0017 |
| MXL | 0.2851 | 0.5891 | 0.8462 | 0.2823 | 0.5847 | 0.8434 | 0.0028 | 0.0044 | 0.0028 |
| PEL | 0.2761 | 0.5655 | 0.8116 | 0.2742 | 0.5626 | 0.8098 | 0.0019 | 0.0029 | 0.0018 |
| PJL | 0.2779 | 0.5603 | 0.8548 | 0.2768 | 0.5586 | 0.8526 | 0.0011 | 0.0017 | 0.0022 |
| PUR | 0.3777 | 0.7024 | 0.8907 | 0.3725 | 0.6952 | 0.8853 | 0.0052 | 0.0072 | 0.0054 |
| STU | 0.2726 | 0.5438 | 0.8327 | 0.2718 | 0.5424 | 0.8319 | 0.0008 | 0.0014 | 0.0008 |
| TSI | 0.2891 | 0.6089 | 0.8989 | 0.2875 | 0.6061 | 0.8965 | 0.0016 | 0.0028 | 0.0024 |
| YRI | 0.3921 | 0.5799 | 0.7133 | 0.3917 | 0.5792 | 0.7126 | 0.0004 | 0.0007 | 0.0007 |
| *Datasets from COVID-19 studies [14, 15]:* | | | | | | | | | |
| Germany | 0.5450 | 0.8076 | 0.9410 | 0.5395 | 0.8010 | 0.9386 | 0.0055 | 0.0066 | 0.0024 |
| Italy | 0.5396 | 0.8035 | 0.9223 | 0.5322 | 0.7952 | 0.9193 | 0.0074 | 0.0083 | 0.0030 |
| Norway | 0.3586 | 0.7098 | 0.9486 | 0.3569 | 0.7064 | 0.9470 | 0.0017 | 0.0034 | 0.0016 |
| Spain | 0.5325 | 0.8233 | 0.9339 | 0.5263 | 0.8155 | 0.9310 | 0.0062 | 0.0078 | 0.0029 |

Supplementary Table 5: Mean estimated imputation $R^2$ values reported by *EagleImp* using the UKB complete reference panel and the *TOPMed Imputation Server*, and improvement (in %) for all variants shared with *TOPMed* for all benchmark datasets classified by UKB reference panel MAF. The number of shared variants is 200,233,690. The difference is calculated as *TOPMed* values subtracted from UKB values, i.e. negative values indicate a better estimation $R^2$ for *TOPMed*.

| Dataset | ***EagleImp* + UKB complete** refpanel MAF ≥ | | | ***TOPMed*** refpanel MAF ≥ | | | **Improvement (in %)** refpanel MAF ≥ | | |
|---|---|---|---|---|---|---|---|---|---|
| | 0.0001 | 0.001 | 0.01 | 0.0001 | 0.001 | 0.01 | 0.0001 | 0.001 | 0.01 |
| *Datasets from 1000 Genomes [6] superpopulations:* | | | | | | | | | |
| AFR | 0.4666 | 0.6889 | 0.8419 | 0.4656 | 0.6691 | 0.8238 | 0.20 | 2.96 | 2.20 |
| AMR | 0.4557 | 0.7785 | 0.9224 | 0.4449 | 0.7769 | 0.9294 | 2.42 | 0.21 | −0.76 |
| EAS | 0.2685 | 0.5154 | 0.7602 | 0.2159 | 0.4270 | 0.6778 | 24.39 | 20.69 | 12.15 |
| EUR | 0.4130 | 0.7771 | 0.9576 | 0.4050 | 0.7733 | 0.9527 | 1.97 | 0.49 | 0.52 |
| SAS | 0.3447 | 0.6402 | 0.8960 | 0.3087 | 0.5832 | 0.8541 | 11.67 | 9.78 | 4.91 |
| *Datasets from 1000 Genomes [6] subpopulations:* | | | | | | | | | |
| ACB | 0.4097 | 0.6397 | 0.8151 | 0.4069 | 0.6241 | 0.7900 | 0.69 | 2.50 | 3.19 |
| ASW | 0.3826 | 0.6348 | 0.8150 | 0.3823 | 0.6240 | 0.7945 | 0.10 | 1.73 | 2.57 |
| BEB | 0.2578 | 0.5348 | 0.8310 | 0.2404 | 0.4960 | 0.7888 | 7.20 | 7.83 | 5.35 |
| CDX | 0.2031 | 0.4301 | 0.6923 | 0.1793 | 0.3820 | 0.6345 | 13.26 | 12.58 | 9.10 |
| CEU | 0.2745 | 0.6218 | 0.9477 | 0.2702 | 0.6131 | 0.9362 | 1.60 | 1.42 | 1.23 |
| CHB | 0.2099 | 0.4409 | 0.7058 | 0.1831 | 0.3866 | 0.6400 | 14.64 | 14.04 | 10.28 |
| CHS | 0.2063 | 0.4327 | 0.6933 | 0.1799 | 0.3794 | 0.6284 | 14.68 | 14.05 | 10.32 |
| CLM | 0.3368 | 0.6787 | 0.9006 | 0.3272 | 0.6647 | 0.8936 | 2.93 | 2.11 | 0.78 |
| ESN | 0.3732 | 0.5650 | 0.7080 | 0.3799 | 0.5612 | 0.6953 | −1.74 | 0.67 | 1.83 |
| FIN | 0.2533 | 0.5762 | 0.9107 | 0.2498 | 0.5735 | 0.9132 | 1.41 | 0.47 | −0.27 |
| GBR | 0.2722 | 0.6200 | 0.9495 | 0.2659 | 0.6062 | 0.9306 | 2.36 | 2.28 | 2.02 |
| GIH | 0.2607 | 0.5462 | 0.8574 | 0.2437 | 0.5097 | 0.8119 | 6.99 | 7.16 | 5.61 |
| GWD | 0.3571 | 0.5630 | 0.7138 | 0.3694 | 0.5634 | 0.7027 | −3.33 | −0.07 | 1.58 |
| IBS | 0.2957 | 0.6426 | 0.9233 | 0.2906 | 0.6364 | 0.9194 | 1.77 | 0.97 | 0.42 |
| ITU | 0.2585 | 0.5363 | 0.8321 | 0.2397 | 0.4957 | 0.7892 | 7.86 | 8.21 | 5.44 |
| JPT | 0.2023 | 0.4262 | 0.6842 | 0.1755 | 0.3721 | 0.6170 | 15.25 | 14.53 | 10.89 |
| KHV | 0.2121 | 0.4463 | 0.7133 | 0.1870 | 0.3956 | 0.6536 | 13.43 | 12.80 | 9.14 |
| LWK | 0.3747 | 0.5848 | 0.7358 | 0.3683 | 0.5627 | 0.7013 | 1.72 | 3.94 | 4.93 |
| MSL | 0.3581 | 0.5548 | 0.6994 | 0.3663 | 0.5512 | 0.6837 | −2.26 | 0.65 | 2.30 |
| MXL | 0.2744 | 0.5896 | 0.8572 | 0.2646 | 0.5749 | 0.8499 | 3.68 | 2.55 | 0.85 |
| PEL | 0.2656 | 0.5658 | 0.8229 | 0.2483 | 0.5380 | 0.8027 | 6.95 | 5.18 | 2.51 |
| PJL | 0.2659 | 0.5564 | 0.8640 | 0.2521 | 0.5264 | 0.8315 | 5.50 | 5.70 | 3.90 |
| PUR | 0.3664 | 0.7036 | 0.9005 | 0.3628 | 0.7023 | 0.9070 | 0.97 | 0.19 | −0.71 |
| STU | 0.2598 | 0.5378 | 0.8404 | 0.2399 | 0.4955 | 0.7879 | 8.26 | 8.54 | 6.67 |
| TSI | 0.2786 | 0.6106 | 0.9107 | 0.2723 | 0.6018 | 0.9051 | 2.32 | 1.46 | 0.63 |
| YRI | 0.3786 | 0.5681 | 0.7121 | 0.3832 | 0.5599 | 0.6937 | −1.21 | 1.47 | 2.65 |
| *Datasets from COVID-19 studies [14, 15]:* | | | | | | | | | |
| Germany | 0.5392 | 0.8146 | 0.9511 | 0.4936 | 0.7682 | 0.9259 | 9.24 | 6.03 | 2.72 |
| Italy | 0.5338 | 0.8123 | 0.9340 | 0.4839 | 0.7616 | 0.9033 | 10.32 | 6.65 | 3.40 |
| Norway | 0.3469 | 0.7120 | 0.9578 | 0.3369 | 0.7003 | 0.9437 | 2.96 | 1.66 | 1.49 |
| Spain | 0.5256 | 0.8319 | 0.9445 | 0.4972 | 0.8131 | 0.9322 | 5.71 | 2.30 | 1.32 |

Supplementary Table 6: Switch error rates (SER) to the 1000 Genomes reference panel (ground truth) for all 1000 Genomes benchmark datasets imputed with *EagleImp* and the *TOPMed Imputation Server* for a) *EagleImp* using the complete UKB reference panel (UKB complete) based on all variants shared with *TOPMed* and 1000 Genomes, and b) *EagleImp* using the rpMAF-filtered UKB reference panel (UKB MAF0.0001) based on all variants shared with *TOPMed* and 1000 Genomes. The number of shared variants is 540,537. The difference is calculated as *TOPMed* values subtracted from UKB values, i.e. negative values indicate a better SER for UKB.

| **Dataset** | **UKB** | ***TOPMed*** | **Difference** | **UKB** | ***TOPMed*** | **Difference** |
|---|---|---|---|---|---|---|
| | a) complete UKB panel | | | b) rpMAF-filtered UKB panel | | |
| *Datasets from 1000 Genomes [6] superpopulations:* | | | | | | |
| AFR | 0.0055 | 0.0083 | −0.0028 | 0.0056 | 0.0083 | −0.0028 |
| AMR | 0.0057 | 0.0056 | 0.0001 | 0.0064 | 0.0056 | 0.0008 |
| EAS | 0.0092 | 0.0127 | −0.0035 | 0.0088 | 0.0132 | −0.0044 |
| EUR | 0.0041 | 0.0046 | −0.0005 | 0.0043 | 0.0046 | −0.0004 |
| SAS | 0.0078 | 0.0103 | −0.0026 | 0.0079 | 0.0104 | −0.0025 |
| *Datasets from 1000 Genomes [6] subpopulations:* | | | | | | |
| ACB | 0.0042 | 0.0096 | −0.0054 | 0.0042 | 0.0096 | −0.0054 |
| ASW | 0.0044 | 0.0100 | −0.0056 | 0.0045 | 0.0100 | −0.0055 |
| BEB | 0.0095 | 0.0128 | −0.0033 | 0.0096 | 0.0129 | −0.0033 |
| CDX | 0.0090 | 0.0130 | −0.0040 | 0.0086 | 0.0135 | −0.0048 |
| CEU | 0.0015 | 0.0030 | −0.0016 | 0.0015 | 0.0030 | −0.0015 |
| CHB | 0.0106 | 0.0150 | −0.0044 | 0.0103 | 0.0155 | −0.0052 |
| CHS | 0.0080 | 0.0132 | −0.0052 | 0.0075 | 0.0137 | −0.0062 |
| CLM | 0.0043 | 0.0056 | −0.0013 | 0.0051 | 0.0056 | −0.0005 |
| ESN | 0.0033 | 0.0076 | −0.0042 | 0.0033 | 0.0076 | −0.0043 |
| FIN | 0.0031 | 0.0033 | −0.0002 | 0.0033 | 0.0033 | 0.0000 |
| GBR | 0.0038 | 0.0050 | −0.0012 | 0.0039 | 0.0050 | −0.0011 |
| GIH | 0.0065 | 0.0109 | −0.0044 | 0.0067 | 0.0109 | −0.0043 |
| GWD | 0.0089 | 0.0117 | −0.0028 | 0.0093 | 0.0117 | −0.0024 |
| IBS | 0.0041 | 0.0048 | −0.0007 | 0.0045 | 0.0048 | −0.0003 |
| ITU | 0.0095 | 0.0132 | −0.0037 | 0.0095 | 0.0132 | −0.0037 |
| JPT | 0.0093 | 0.0152 | −0.0059 | 0.0089 | 0.0156 | −0.0067 |
| KHV | 0.0092 | 0.0131 | −0.0039 | 0.0087 | 0.0135 | −0.0048 |
| LWK | 0.0063 | 0.0136 | −0.0072 | 0.0065 | 0.0136 | −0.0071 |
| MSL | 0.0073 | 0.0125 | −0.0053 | 0.0074 | 0.0126 | −0.0051 |
| MXL | 0.0063 | 0.0067 | −0.0004 | 0.0068 | 0.0067 | 0.0001 |
| PEL | 0.0071 | 0.0076 | −0.0005 | 0.0073 | 0.0076 | −0.0004 |
| PJL | 0.0059 | 0.0097 | −0.0038 | 0.0064 | 0.0097 | −0.0033 |
| PUR | 0.0054 | 0.0055 | 0.0000 | 0.0068 | 0.0055 | 0.0014 |
| STU | 0.0076 | 0.0128 | −0.0052 | 0.0077 | 0.0128 | −0.0051 |
| TSI | 0.0073 | 0.0076 | −0.0003 | 0.0075 | 0.0076 | −0.0001 |
| YRI | 0.0035 | 0.0085 | −0.0050 | 0.0034 | 0.0085 | −0.0051 |

Supplementary Table 7: Mean absolute error (MAE) to the 1000 Genomes reference panel (ground truth) stratified by four rpMAF range categories for all 1000 Genomes benchmark datasets imputed with *EagleImp* using the complete UK Biobank reference panel (UKB complete) and the *TOPMed Imputation Server*. MAE was calculated on all variants from the complete UK Biobank reference panel shared with *TOPMed* and 1000 Genomes. The number of shared variants is 56,908,240.

| Dataset | **UKB complete** refpanel MAF range | | | | ***TOPMed*** refpanel MAF range | | | |
|---|---|---|---|---|---|---|---|---|
| | [0.0001, 0.001) | [0.001, 0.01) | [0.01, 0.1) | [0.1, 0.5] | [0.0001, 0.001) | [0.001, 0.01) | [0.01, 0.1) | [0.1, 0.5] |
| *Datasets from 1000 Genomes [6] superpopulations:* | | | | | | | | |
| AFR | 0.0073 | 0.0198 | 0.0212 | 0.0514 | 0.0048 | 0.0121 | 0.0149 | 0.0344 |
| AMR | 0.0023 | 0.0055 | 0.0134 | 0.0359 | 0.0024 | 0.0055 | 0.0102 | 0.0248 |
| EAS | 0.0032 | 0.0047 | 0.0140 | 0.0435 | 0.0033 | 0.0059 | 0.0149 | 0.0440 |
| EUR | 0.0010 | 0.0029 | 0.0086 | 0.0227 | 0.0018 | 0.0042 | 0.0084 | 0.0180 |
| SAS | 0.0023 | 0.0041 | 0.0131 | 0.0360 | 0.0030 | 0.0060 | 0.0150 | 0.0415 |
| *Datasets from 1000 Genomes [6] subpopulations:* | | | | | | | | |
| ACB | 0.0052 | 0.0139 | 0.0152 | 0.0375 | 0.0045 | 0.0115 | 0.0146 | 0.0341 |
| ASW | 0.0058 | 0.0156 | 0.0179 | 0.0441 | 0.0046 | 0.0119 | 0.0159 | 0.0382 |
| BEB | 0.0029 | 0.0051 | 0.0164 | 0.0447 | 0.0033 | 0.0065 | 0.0169 | 0.0473 |
| CDX | 0.0031 | 0.0046 | 0.0138 | 0.0428 | 0.0031 | 0.0057 | 0.0144 | 0.0417 |
| CEU | 0.0008 | 0.0019 | 0.0050 | 0.0149 | 0.0017 | 0.0037 | 0.0069 | 0.0137 |
| CHB | 0.0034 | 0.0048 | 0.0145 | 0.0450 | 0.0034 | 0.0061 | 0.0157 | 0.0468 |
| CHS | 0.0033 | 0.0046 | 0.0137 | 0.0429 | 0.0033 | 0.0060 | 0.0153 | 0.0455 |
| CLM | 0.0020 | 0.0050 | 0.0116 | 0.0308 | 0.0024 | 0.0057 | 0.0103 | 0.0244 |
| ESN | 0.0058 | 0.0159 | 0.0164 | 0.0403 | 0.0038 | 0.0093 | 0.0113 | 0.0248 |
| FIN | 0.0010 | 0.0026 | 0.0075 | 0.0202 | 0.0017 | 0.0037 | 0.0067 | 0.0119 |
| GBR | 0.0008 | 0.0018 | 0.0044 | 0.0130 | 0.0017 | 0.0039 | 0.0075 | 0.0151 |
| GIH | 0.0019 | 0.0033 | 0.0103 | 0.0285 | 0.0029 | 0.0058 | 0.0147 | 0.0396 |
| GWD | 0.0103 | 0.0291 | 0.0297 | 0.0713 | 0.0050 | 0.0134 | 0.0162 | 0.0383 |
| IBS | 0.0012 | 0.0038 | 0.0113 | 0.0290 | 0.0019 | 0.0047 | 0.0098 | 0.0223 |
| ITU | 0.0028 | 0.0048 | 0.0156 | 0.0426 | 0.0032 | 0.0063 | 0.0165 | 0.0455 |
| JPT | 0.0031 | 0.0046 | 0.0138 | 0.0426 | 0.0033 | 0.0063 | 0.0162 | 0.0480 |
| KHV | 0.0032 | 0.0047 | 0.0143 | 0.0441 | 0.0032 | 0.0059 | 0.0147 | 0.0430 |
| LWK | 0.0078 | 0.0211 | 0.0254 | 0.0604 | 0.0060 | 0.0156 | 0.0207 | 0.0491 |
| MSL | 0.0091 | 0.0243 | 0.0250 | 0.0601 | 0.0053 | 0.0133 | 0.0161 | 0.0374 |
| MXL | 0.0023 | 0.0052 | 0.0141 | 0.0382 | 0.0024 | 0.0052 | 0.0105 | 0.0264 |
| PEL | 0.0025 | 0.0048 | 0.0138 | 0.0388 | 0.0025 | 0.0050 | 0.0107 | 0.0284 |
| PJL | 0.0020 | 0.0037 | 0.0120 | 0.0327 | 0.0027 | 0.0055 | 0.0135 | 0.0360 |
| PUR | 0.0024 | 0.0068 | 0.0144 | 0.0368 | 0.0025 | 0.0060 | 0.0100 | 0.0229 |
| STU | 0.0022 | 0.0037 | 0.0117 | 0.0327 | 0.0032 | 0.0063 | 0.0163 | 0.0452 |
| TSI | 0.0013 | 0.0040 | 0.0134 | 0.0336 | 0.0020 | 0.0048 | 0.0109 | 0.0259 |
| YRI | 0.0061 | 0.0168 | 0.0173 | 0.0423 | 0.0043 | 0.0107 | 0.0129 | 0.0290 |

Supplementary Table 8: Mean absolute error (MAE) to the 1000 Genomes reference panel (ground truth) stratified by four rpMAF range categories for all 1000 Genomes benchmark datasets imputed with *EagleImp* using the rpMAF-filtered UK Biobank reference panel (UKB MAF0.0001) and the *TOPMed Imputation Server*. MAE was calculated on all variants from the rpMAF-filtered UK Biobank reference panel shared with *TOPMed* and 1000 Genomes. The number of shared variants is 30,098,506.

| **Dataset** | **UKB MAF0.0001** refpanel MAF range | | | | ***TOPMed*** refpanel MAF range | | | |
|---|---|---|---|---|---|---|---|---|
| | [0.0001, 0.001) | [0.001, 0.01) | [0.01, 0.1) | [0.1, 0.5] | [0.0001, 0.001) | [0.001, 0.01) | [0.01, 0.1) | [0.1, 0.5] |
| *Datasets from 1000 Genomes [6] superpopulations:* | | | | | | | | |
| AFR | 0.0074 | 0.0200 | 0.0213 | 0.0515 | 0.0055 | 0.0136 | 0.0168 | 0.0352 |
| AMR | 0.0025 | 0.0060 | 0.0141 | 0.0369 | 0.0032 | 0.0070 | 0.0122 | 0.0257 |
| EAS | 0.0033 | 0.0046 | 0.0136 | 0.0424 | 0.0040 | 0.0075 | 0.0169 | 0.0449 |
| EUR | 0.0011 | 0.0029 | 0.0085 | 0.0217 | 0.0026 | 0.0058 | 0.0105 | 0.0188 |
| SAS | 0.0025 | 0.0041 | 0.0131 | 0.0359 | 0.0038 | 0.0076 | 0.0171 | 0.0423 |
| *Datasets from 1000 Genomes [6] subpopulations:* | | | | | | | | |
| ACB | 0.0053 | 0.0139 | 0.0152 | 0.0372 | 0.0052 | 0.0130 | 0.0166 | 0.0350 |
| ASW | 0.0060 | 0.0158 | 0.0180 | 0.0440 | 0.0053 | 0.0133 | 0.0179 | 0.0390 |
| BEB | 0.0030 | 0.0051 | 0.0163 | 0.0445 | 0.0040 | 0.0081 | 0.0189 | 0.0482 |
| CDX | 0.0032 | 0.0045 | 0.0134 | 0.0418 | 0.0038 | 0.0073 | 0.0164 | 0.0426 |
| CEU | 0.0009 | 0.0018 | 0.0047 | 0.0133 | 0.0024 | 0.0053 | 0.0090 | 0.0146 |
| CHB | 0.0035 | 0.0048 | 0.0141 | 0.0439 | 0.0041 | 0.0077 | 0.0177 | 0.0476 |
| CHS | 0.0033 | 0.0045 | 0.0133 | 0.0418 | 0.0041 | 0.0076 | 0.0174 | 0.0463 |
| CLM | 0.0022 | 0.0055 | 0.0123 | 0.0319 | 0.0031 | 0.0072 | 0.0124 | 0.0252 |
| ESN | 0.0059 | 0.0160 | 0.0164 | 0.0402 | 0.0045 | 0.0107 | 0.0133 | 0.0256 |
| FIN | 0.0011 | 0.0025 | 0.0073 | 0.0191 | 0.0025 | 0.0053 | 0.0087 | 0.0127 |
| GBR | 0.0009 | 0.0017 | 0.0041 | 0.0115 | 0.0025 | 0.0055 | 0.0096 | 0.0159 |
| GIH | 0.0020 | 0.0033 | 0.0104 | 0.0285 | 0.0037 | 0.0074 | 0.0167 | 0.0404 |
| GWD | 0.0105 | 0.0295 | 0.0299 | 0.0717 | 0.0057 | 0.0148 | 0.0182 | 0.0392 |
| IBS | 0.0013 | 0.0038 | 0.0114 | 0.0284 | 0.0027 | 0.0063 | 0.0119 | 0.0231 |
| ITU | 0.0029 | 0.0048 | 0.0155 | 0.0421 | 0.0039 | 0.0079 | 0.0185 | 0.0464 |
| JPT | 0.0032 | 0.0045 | 0.0134 | 0.0414 | 0.0041 | 0.0078 | 0.0182 | 0.0489 |
| KHV | 0.0033 | 0.0046 | 0.0139 | 0.0430 | 0.0039 | 0.0074 | 0.0168 | 0.0438 |
| LWK | 0.0079 | 0.0213 | 0.0256 | 0.0606 | 0.0067 | 0.0171 | 0.0227 | 0.0499 |
| MSL | 0.0093 | 0.0245 | 0.0251 | 0.0602 | 0.0060 | 0.0147 | 0.0180 | 0.0382 |
| MXL | 0.0025 | 0.0055 | 0.0145 | 0.0386 | 0.0031 | 0.0068 | 0.0126 | 0.0272 |
| PEL | 0.0026 | 0.0049 | 0.0137 | 0.0384 | 0.0032 | 0.0066 | 0.0127 | 0.0292 |
| PJL | 0.0022 | 0.0039 | 0.0123 | 0.0333 | 0.0034 | 0.0071 | 0.0156 | 0.0368 |
| PUR | 0.0028 | 0.0077 | 0.0158 | 0.0393 | 0.0032 | 0.0075 | 0.0121 | 0.0238 |
| STU | 0.0023 | 0.0037 | 0.0116 | 0.0323 | 0.0039 | 0.0079 | 0.0184 | 0.0460 |
| TSI | 0.0014 | 0.0041 | 0.0134 | 0.0330 | 0.0027 | 0.0064 | 0.0130 | 0.0267 |
| YRI | 0.0062 | 0.0169 | 0.0174 | 0.0422 | 0.0050 | 0.0121 | 0.0149 | 0.0298 |

Supplementary Table 9: Correlation $r^2$ between the 1000 Genomes reference panel (ground truth) and all 1000 Genomes benchmark datasets imputed with *EagleImp* using the complete UK Biobank reference panel (UKB complete) and the *TOPMed Imputation Server*. Correlation $r^2$ was calculated separately for each variant. Results are stratified by four rpMAF range categories. The final values state the averages over all variants in the corresponding rpMAF category from the complete UK Biobank reference panel shared with *TOPMed* and 1000 Genomes. The number of shared variants is 56,908,240. Note, $r^2$ values which could not be calculated (e.g. due to missing variance) were set to $r^2 = 0$ which leads to a strong bias especially in the low rpMAF categories.

| **Dataset** | **UKB complete** refpanel MAF range | | | | ***TOPMed*** refpanel MAF range | | | |
|---|---|---|---|---|---|---|---|---|
| | [0.0001, 0.001) | [0.001, 0.01) | [0.01, 0.1) | [0.1, 0.5] | [0.0001, 0.001) | [0.001, 0.01) | [0.01, 0.1) | [0.1, 0.5] |
| *Datasets from 1000 Genomes [6] superpopulations:* | | | | | | | | |
| AFR | 0.4472 | 0.4688 | 0.7020 | 0.8838 | 0.5352 | 0.5255 | 0.7814 | 0.9472 |
| AMR | 0.2962 | 0.5682 | 0.8483 | 0.9405 | 0.3476 | 0.6676 | 0.9349 | 0.9786 |
| EAS | 0.0517 | 0.0900 | 0.4635 | 0.8788 | 0.0781 | 0.1081 | 0.5015 | 0.9199 |
| EUR | 0.1621 | 0.5307 | 0.9116 | 0.9636 | 0.1973 | 0.6092 | 0.9567 | 0.9858 |
| SAS | 0.1254 | 0.2301 | 0.7845 | 0.9374 | 0.1510 | 0.2680 | 0.8430 | 0.9600 |
| *Datasets from 1000 Genomes [6] subpopulations:* | | | | | | | | |
| ACB | 0.4068 | 0.4484 | 0.6704 | 0.9254 | 0.4361 | 0.4701 | 0.6917 | 0.9538 |
| ASW | 0.3318 | 0.4335 | 0.6713 | 0.9187 | 0.3651 | 0.4654 | 0.7006 | 0.9543 |
| BEB | 0.0736 | 0.1399 | 0.6670 | 0.9194 | 0.0924 | 0.1596 | 0.7151 | 0.9522 |
| CDX | 0.0399 | 0.0719 | 0.3934 | 0.8580 | 0.0526 | 0.0802 | 0.4118 | 0.8934 |
| CEU | 0.0398 | 0.2647 | 0.9182 | 0.9766 | 0.0430 | 0.2747 | 0.9355 | 0.9896 |
| CHB | 0.0409 | 0.0731 | 0.4118 | 0.8653 | 0.0559 | 0.0822 | 0.4329 | 0.9010 |
| CHS | 0.0418 | 0.0716 | 0.3892 | 0.8577 | 0.0546 | 0.0790 | 0.4052 | 0.8887 |
| CLM | 0.1385 | 0.4228 | 0.8203 | 0.9492 | 0.1535 | 0.4584 | 0.8715 | 0.9786 |
| ESN | 0.3908 | 0.4059 | 0.4760 | 0.8609 | 0.4340 | 0.4384 | 0.5041 | 0.9088 |
| FIN | 0.0253 | 0.1753 | 0.8452 | 0.9677 | 0.0297 | 0.1924 | 0.8820 | 0.9901 |
| GBR | 0.0349 | 0.2529 | 0.9188 | 0.9793 | 0.0361 | 0.2563 | 0.9261 | 0.9882 |
| GIH | 0.0785 | 0.1565 | 0.7278 | 0.9508 | 0.0821 | 0.1630 | 0.7481 | 0.9626 |
| GWD | 0.2973 | 0.3545 | 0.4651 | 0.8135 | 0.3939 | 0.4318 | 0.5306 | 0.9132 |
| IBS | 0.0572 | 0.3105 | 0.8604 | 0.9532 | 0.0704 | 0.3554 | 0.9187 | 0.9822 |
| ITU | 0.0700 | 0.1377 | 0.6649 | 0.9237 | 0.0869 | 0.1555 | 0.7090 | 0.9543 |
| JPT | 0.0383 | 0.0680 | 0.3757 | 0.8500 | 0.0493 | 0.0744 | 0.3889 | 0.8773 |
| KHV | 0.0437 | 0.0783 | 0.4248 | 0.8704 | 0.0588 | 0.0884 | 0.4489 | 0.9092 |
| LWK | 0.3312 | 0.3818 | 0.4841 | 0.8343 | 0.3757 | 0.4213 | 0.5271 | 0.8955 |
| MSL | 0.3356 | 0.3743 | 0.4544 | 0.8206 | 0.4008 | 0.4269 | 0.4984 | 0.8933 |
| MXL | 0.0671 | 0.2716 | 0.7396 | 0.9351 | 0.0802 | 0.3024 | 0.8007 | 0.9769 |
| PEL | 0.0654 | 0.2507 | 0.6700 | 0.9263 | 0.0772 | 0.2743 | 0.7274 | 0.9716 |
| PJL | 0.0817 | 0.1703 | 0.7431 | 0.9444 | 0.0931 | 0.1862 | 0.7811 | 0.9669 |
| PUR | 0.1848 | 0.4641 | 0.8155 | 0.9391 | 0.2122 | 0.5257 | 0.8896 | 0.9802 |
| STU | 0.0830 | 0.1508 | 0.6862 | 0.9413 | 0.0881 | 0.1564 | 0.7050 | 0.9544 |
| TSI | 0.0416 | 0.2355 | 0.8349 | 0.9449 | 0.0576 | 0.2861 | 0.9051 | 0.9789 |
| YRI | 0.3990 | 0.4043 | 0.4782 | 0.8598 | 0.4436 | 0.4361 | 0.5066 | 0.9069 |

Supplementary Table 10: Correlation $r^2$ between the 1000 Genomes reference panel (ground truth) and all 1000 Genomes benchmark datasets imputed with *EagleImp* using the rpMAF-filtered UK Biobank reference panel (UKB MAF0.0001) and the *TOPMed Imputation Server*. Correlation $r^2$ was calculated separately for each variant. Results are stratified by four rpMAF range categories. The final values state the averages over all variants in the corresponding rpMAF category from the rpMAF-filtered UK Biobank reference panel shared with *TOPMed* and 1000 Genomes. The number of shared variants is 30,098,506. Note, $r^2$ values which could not be calculated (e.g. due to missing variance) were set to $r^2 = 0$ which leads to a strong bias especially in the low rpMAF categories.

| Dataset | **UKB MAF0.0001** refpanel MAF range | | | | ***TOPMed*** refpanel MAF range | | | |
|---|---|---|---|---|---|---|---|---|
| | [0.0001, 0.001) | [0.001, 0.01) | [0.01, 0.1) | [0.1, 0.5] | [0.0001, 0.001) | [0.001, 0.01) | [0.01, 0.1) | [0.1, 0.5] |
| *Datasets from 1000 Genomes [6] superpopulations:* | | | | | | | | |
| AFR | 0.4463 | 0.4688 | 0.7039 | 0.8848 | 0.5350 | 0.5251 | 0.7806 | 0.9467 |
| AMR | 0.2822 | 0.5501 | 0.8425 | 0.9396 | 0.3474 | 0.6671 | 0.9338 | 0.9780 |
| EAS | 0.0522 | 0.0909 | 0.4667 | 0.8830 | 0.0781 | 0.1080 | 0.5010 | 0.9194 |
| EUR | 0.1605 | 0.5282 | 0.9134 | 0.9667 | 0.1972 | 0.6087 | 0.9556 | 0.9852 |
| SAS | 0.1244 | 0.2299 | 0.7856 | 0.9386 | 0.1509 | 0.2678 | 0.8421 | 0.9595 |
| *Datasets from 1000 Genomes [6] subpopulations:* | | | | | | | | |
| ACB | 0.4070 | 0.4490 | 0.6721 | 0.9270 | 0.4359 | 0.4698 | 0.6909 | 0.9533 |
| ASW | 0.3313 | 0.4334 | 0.6725 | 0.9200 | 0.3649 | 0.4650 | 0.6998 | 0.9537 |
| BEB | 0.0731 | 0.1399 | 0.6686 | 0.9210 | 0.0924 | 0.1595 | 0.7143 | 0.9516 |
| CDX | 0.0402 | 0.0724 | 0.3949 | 0.8612 | 0.0525 | 0.0802 | 0.4113 | 0.8929 |
| CEU | 0.0400 | 0.2654 | 0.9217 | 0.9806 | 0.0430 | 0.2744 | 0.9345 | 0.9890 |
| CHB | 0.0412 | 0.0736 | 0.4139 | 0.8690 | 0.0559 | 0.0822 | 0.4324 | 0.9005 |
| CHS | 0.0421 | 0.0721 | 0.3908 | 0.8610 | 0.0545 | 0.0789 | 0.4047 | 0.8882 |
| CLM | 0.1344 | 0.4134 | 0.8157 | 0.9482 | 0.1534 | 0.4580 | 0.8705 | 0.9780 |
| ESN | 0.3910 | 0.4064 | 0.4764 | 0.8615 | 0.4338 | 0.4381 | 0.5035 | 0.9083 |
| FIN | 0.0252 | 0.1750 | 0.8472 | 0.9713 | 0.0297 | 0.1923 | 0.8810 | 0.9895 |
| GBR | 0.0351 | 0.2537 | 0.9220 | 0.9830 | 0.0361 | 0.2561 | 0.9250 | 0.9876 |
| GIH | 0.0781 | 0.1564 | 0.7282 | 0.9517 | 0.0821 | 0.1629 | 0.7473 | 0.9620 |
| GWD | 0.2957 | 0.3535 | 0.4644 | 0.8134 | 0.3938 | 0.4314 | 0.5300 | 0.9127 |
| IBS | 0.0563 | 0.3080 | 0.8606 | 0.9557 | 0.0704 | 0.3552 | 0.9177 | 0.9816 |
| ITU | 0.0701 | 0.1382 | 0.6668 | 0.9257 | 0.0868 | 0.1554 | 0.7082 | 0.9538 |
| JPT | 0.0387 | 0.0686 | 0.3772 | 0.8534 | 0.0493 | 0.0743 | 0.3885 | 0.8768 |
| KHV | 0.0440 | 0.0789 | 0.4269 | 0.8742 | 0.0587 | 0.0883 | 0.4484 | 0.9086 |
| LWK | 0.3303 | 0.3813 | 0.4838 | 0.8348 | 0.3756 | 0.4209 | 0.5265 | 0.8949 |
| MSL | 0.3349 | 0.3741 | 0.4543 | 0.8209 | 0.4006 | 0.4265 | 0.4978 | 0.8928 |
| MXL | 0.0650 | 0.2665 | 0.7372 | 0.9353 | 0.0801 | 0.3021 | 0.7998 | 0.9763 |
| PEL | 0.0644 | 0.2485 | 0.6702 | 0.9281 | 0.0772 | 0.2741 | 0.7266 | 0.9710 |
| PJL | 0.0804 | 0.1689 | 0.7413 | 0.9443 | 0.0931 | 0.1860 | 0.7803 | 0.9664 |
| PUR | 0.1762 | 0.4473 | 0.8060 | 0.9356 | 0.2121 | 0.5252 | 0.8887 | 0.9797 |
| STU | 0.0832 | 0.1511 | 0.6878 | 0.9428 | 0.0880 | 0.1563 | 0.7042 | 0.9539 |
| TSI | 0.0409 | 0.2339 | 0.8351 | 0.9474 | 0.0575 | 0.2859 | 0.9041 | 0.9784 |
| YRI | 0.3992 | 0.4047 | 0.4785 | 0.8604 | 0.4435 | 0.4358 | 0.5060 | 0.9064 |

Supplementary Table 11: Correlation $r^2$ between the 1000 Genomes reference panel (ground truth) and all 1000 Genomes Project benchmark datasets imputed with *EagleImp* using the complete UK Biobank reference panel (UKB complete) and the *TOPMed Imputation Server*. Correlation $r^2$ was calculated for the complete dataset over all data points from all variants and samples (but separately for each rpMAF category in each chromosome file, but not separately for each variant). Results are stratified by four rpMAF range categories. Computation is based on 56,908,240 shared variants in the complete UK Biobank reference panel, the *TOPMed r3* panel and the 1000 Genomes reference panel.

| **Dataset** | **UKB complete** refpanel MAF range | | | | ***TOPMed*** refpanel MAF range | | | |
|---|---|---|---|---|---|---|---|---|
| | [0.0001, 0.001) | [0.001, 0.01) | [0.01, 0.1) | [0.1, 0.5] | [0.0001, 0.001) | [0.001, 0.01) | [0.01, 0.1) | [0.1, 0.5] |
| *Datasets from 1000 Genomes [6] superpopulations:* | | | | | | | | |
| AFR | 0.8371 | 0.9093 | 0.9527 | 0.9463 | 0.8589 | 0.9344 | 0.9616 | 0.9739 |
| AMR | 0.8668 | 0.9444 | 0.9645 | 0.9597 | 0.8165 | 0.9201 | 0.9650 | 0.9826 |
| EAS | 0.8389 | 0.9541 | 0.9663 | 0.9552 | 0.7944 | 0.9207 | 0.9614 | 0.9727 |
| EUR | 0.9033 | 0.9613 | 0.9752 | 0.9730 | 0.8078 | 0.9184 | 0.9660 | 0.9869 |
| SAS | 0.8638 | 0.9553 | 0.9662 | 0.9599 | 0.7908 | 0.9146 | 0.9579 | 0.9725 |
| *Datasets from 1000 Genomes [6] subpopulations:* | | | | | | | | |
| ACB | 0.8744 | 0.9347 | 0.9658 | 0.9606 | 0.8588 | 0.9345 | 0.9619 | 0.9748 |
| ASW | 0.8502 | 0.9197 | 0.9582 | 0.9523 | 0.8460 | 0.9271 | 0.9585 | 0.9719 |
| BEB | 0.8425 | 0.9473 | 0.9582 | 0.9502 | 0.7832 | 0.9124 | 0.9552 | 0.9688 |
| CDX | 0.8434 | 0.9551 | 0.9668 | 0.9559 | 0.8003 | 0.9223 | 0.9624 | 0.9743 |
| CEU | 0.9127 | 0.9705 | 0.9844 | 0.9812 | 0.8104 | 0.9214 | 0.9688 | 0.9894 |
| CHB | 0.8315 | 0.9527 | 0.9653 | 0.9537 | 0.7901 | 0.9198 | 0.9605 | 0.9715 |
| CHS | 0.8381 | 0.9549 | 0.9671 | 0.9560 | 0.7918 | 0.9203 | 0.9609 | 0.9720 |
| CLM | 0.8783 | 0.9485 | 0.9686 | 0.9645 | 0.8128 | 0.9177 | 0.9641 | 0.9825 |
| ESN | 0.8731 | 0.9319 | 0.9648 | 0.9596 | 0.8800 | 0.9474 | 0.9682 | 0.9811 |
| FIN | 0.9064 | 0.9644 | 0.9782 | 0.9760 | 0.8117 | 0.9206 | 0.9683 | 0.9899 |
| GBR | 0.9143 | 0.9712 | 0.9857 | 0.9829 | 0.8089 | 0.9194 | 0.9674 | 0.9885 |
| GIH | 0.8813 | 0.9621 | 0.9730 | 0.9682 | 0.7924 | 0.9151 | 0.9586 | 0.9743 |
| GWD | 0.7743 | 0.8672 | 0.9341 | 0.9254 | 0.8555 | 0.9314 | 0.9606 | 0.9723 |
| IBS | 0.8958 | 0.9535 | 0.9682 | 0.9666 | 0.8055 | 0.9161 | 0.9638 | 0.9846 |
| ITU | 0.8463 | 0.9493 | 0.9600 | 0.9529 | 0.7866 | 0.9133 | 0.9560 | 0.9703 |
| JPT | 0.8418 | 0.9544 | 0.9668 | 0.9562 | 0.7896 | 0.9190 | 0.9599 | 0.9710 |
| KHV | 0.8404 | 0.9536 | 0.9655 | 0.9544 | 0.7985 | 0.9212 | 0.9617 | 0.9734 |
| LWK | 0.8225 | 0.8986 | 0.9414 | 0.9343 | 0.8317 | 0.9171 | 0.9509 | 0.9622 |
| MSL | 0.8113 | 0.8927 | 0.9454 | 0.9379 | 0.8577 | 0.9322 | 0.9605 | 0.9722 |
| MXL | 0.8616 | 0.9453 | 0.9629 | 0.9575 | 0.8135 | 0.9195 | 0.9648 | 0.9823 |
| PEL | 0.8605 | 0.9504 | 0.9651 | 0.9589 | 0.8194 | 0.9231 | 0.9662 | 0.9820 |
| PJL | 0.8744 | 0.9577 | 0.9688 | 0.9633 | 0.7994 | 0.9170 | 0.9604 | 0.9763 |
| PUR | 0.8652 | 0.9359 | 0.9611 | 0.9574 | 0.8184 | 0.9197 | 0.9643 | 0.9831 |
| STU | 0.8720 | 0.9592 | 0.9699 | 0.9635 | 0.7884 | 0.9136 | 0.9565 | 0.9704 |
| TSI | 0.8913 | 0.9505 | 0.9628 | 0.9613 | 0.8038 | 0.9154 | 0.9623 | 0.9826 |
| YRI | 0.8656 | 0.9280 | 0.9629 | 0.9577 | 0.8704 | 0.9423 | 0.9658 | 0.9787 |

Supplementary Table 12: Correlation $r^2$ between the 1000 Genomes reference panel (ground truth) and all 1000 Genomes Project benchmark datasets imputed with *EagleImp* using the rpMAF-filtered UK Biobank reference panel (UKB MAF0.0001) and the *TOPMed Imputation Server*. Correlation $r^2$ was calculated for the complete dataset over all data points from all variants and samples (but separately for each rpMAF category in each chromosome file, but not separately for each variant). Results are stratified by four rpMAF range categories. Computation is based on 30,098,506 shared variants in the rpMAF-filtered UK Biobank reference panel, the *TOPMed r3* panel and the 1000 Genomes reference panel.

| **Dataset** | **UKB MAF0.0001** refpanel MAF range | | | | ***TOPMed*** refpanel MAF range | | | |
|---|---|---|---|---|---|---|---|---|
| | [0.0001, 0.001) | [0.001, 0.01) | [0.01, 0.1) | [0.1, 0.5] | [0.0001, 0.001) | [0.001, 0.01) | [0.01, 0.1) | [0.1, 0.5] |
| *Datasets from 1000 Genomes [6] superpopulations:* | | | | | | | | |
| AFR | 0.8314 | 0.9087 | 0.9527 | 0.9466 | 0.8234 | 0.9165 | 0.9491 | 0.9714 |
| AMR | 0.8517 | 0.9401 | 0.9632 | 0.9593 | 0.7579 | 0.8858 | 0.9498 | 0.9800 |
| EAS | 0.8316 | 0.9548 | 0.9676 | 0.9570 | 0.7379 | 0.8861 | 0.9470 | 0.9702 |
| EUR | 0.8927 | 0.9616 | 0.9762 | 0.9755 | 0.7417 | 0.8802 | 0.9500 | 0.9842 |
| SAS | 0.8539 | 0.9550 | 0.9665 | 0.9607 | 0.7323 | 0.8782 | 0.9428 | 0.9698 |
| *Datasets from 1000 Genomes [6] subpopulations:* | | | | | | | | |
| ACB | 0.8693 | 0.9346 | 0.9661 | 0.9612 | 0.8222 | 0.9159 | 0.9491 | 0.9722 |
| ASW | 0.8437 | 0.9190 | 0.9584 | 0.9530 | 0.8071 | 0.9067 | 0.9452 | 0.9693 |
| BEB | 0.8327 | 0.9472 | 0.9588 | 0.9512 | 0.7250 | 0.8762 | 0.9401 | 0.9661 |
| CDX | 0.8358 | 0.9557 | 0.9680 | 0.9576 | 0.7437 | 0.8877 | 0.9481 | 0.9717 |
| CEU | 0.9027 | 0.9714 | 0.9860 | 0.9844 | 0.7437 | 0.8829 | 0.9528 | 0.9868 |
| CHB | 0.8241 | 0.9533 | 0.9666 | 0.9555 | 0.7339 | 0.8851 | 0.9460 | 0.9690 |
| CHS | 0.8309 | 0.9555 | 0.9683 | 0.9577 | 0.7357 | 0.8857 | 0.9465 | 0.9695 |
| CLM | 0.8630 | 0.9442 | 0.9673 | 0.9641 | 0.7534 | 0.8831 | 0.9487 | 0.9798 |
| ESN | 0.8681 | 0.9318 | 0.9649 | 0.9599 | 0.8451 | 0.9303 | 0.9559 | 0.9786 |
| FIN | 0.8959 | 0.9649 | 0.9794 | 0.9787 | 0.7459 | 0.8824 | 0.9524 | 0.9873 |
| GBR | 0.9043 | 0.9720 | 0.9871 | 0.9858 | 0.7420 | 0.8809 | 0.9514 | 0.9859 |
| GIH | 0.8713 | 0.9617 | 0.9732 | 0.9688 | 0.7336 | 0.8785 | 0.9434 | 0.9716 |
| GWD | 0.7672 | 0.8658 | 0.9337 | 0.9255 | 0.8200 | 0.9139 | 0.9482 | 0.9697 |
| IBS | 0.8845 | 0.9531 | 0.9687 | 0.9685 | 0.7399 | 0.8782 | 0.9479 | 0.9820 |
| ITU | 0.8372 | 0.9495 | 0.9608 | 0.9541 | 0.7286 | 0.8770 | 0.9409 | 0.9676 |
| JPT | 0.8346 | 0.9552 | 0.9681 | 0.9580 | 0.7330 | 0.8844 | 0.9454 | 0.9684 |
| KHV | 0.8330 | 0.9543 | 0.9668 | 0.9562 | 0.7418 | 0.8865 | 0.9474 | 0.9709 |
| LWK | 0.8163 | 0.8976 | 0.9413 | 0.9346 | 0.7967 | 0.8988 | 0.9385 | 0.9597 |
| MSL | 0.8055 | 0.8919 | 0.9452 | 0.9380 | 0.8241 | 0.9150 | 0.9482 | 0.9697 |
| MXL | 0.8483 | 0.9425 | 0.9623 | 0.9578 | 0.7539 | 0.8841 | 0.9496 | 0.9797 |
| PEL | 0.8505 | 0.9496 | 0.9656 | 0.9599 | 0.7614 | 0.8881 | 0.9515 | 0.9794 |
| PJL | 0.8628 | 0.9565 | 0.9682 | 0.9634 | 0.7398 | 0.8803 | 0.9452 | 0.9737 |
| PUR | 0.8450 | 0.9278 | 0.9579 | 0.9553 | 0.7606 | 0.8867 | 0.9489 | 0.9805 |
| STU | 0.8630 | 0.9590 | 0.9704 | 0.9645 | 0.7304 | 0.8774 | 0.9414 | 0.9678 |
| TSI | 0.8802 | 0.9503 | 0.9633 | 0.9631 | 0.7378 | 0.8770 | 0.9464 | 0.9799 |
| YRI | 0.8608 | 0.9278 | 0.9630 | 0.9580 | 0.8354 | 0.9251 | 0.9535 | 0.9761 |

Supplementary Table 13: Results of Bayesian fine-mapping analysis using the *Sum of Single Effects (SuSiE)* R package [23] and summary statistics of the Italian GWAS dataset imputed with *EagleImp* (rpMAF-prefiltered UKB reference panel). A total of 20 variants were identified that belong to the 95% most likely to be causal. *chr3:45859597:C:T* respresents *rs73064425*. *chr3:45867022:C:G* respresents *rs35652899*. Columns are labeled *SNP (single nucleotide polymorphism)* and *PIP (posterior inclusion probability)*.

| SNP | PIP |
|---|---|
| chr3:45859597:C:T | 0.0632594368565981 |
| chr3:45858159:A:G | 0.0632594368565981 |
| chr3:45818159:G:A | 0.0623178473600295 |
| chr3:45820440:G:A | 0.0623178473600295 |
| chr3:45859142:G:C | 0.0610863384044361 |
| chr3:45847198:A:G | 0.0601588521178198 |
| chr3:45848429:A:T | 0.0601588521178198 |
| chr3:45848457:C:T | 0.0601588521178198 |
| chr3:45838989:T:C | 0.0597951831110618 |
| chr3:45830416:G:A | 0.0499762588414211 |
| chr3:45812384:ATTTTTTT:ATTTTTT | 0.0467984942129043 |
| chr3:45821460:T:C | 0.0417175515743682 |
| chr3:45823240:T:C | 0.0417175515743682 |
| chr3:45825948:A:G | 0.0417175515743682 |
| chr3:45812744:CTTTTTT:CTTT | 0.0397141658066035 |
| chr3:45834967:G:GA | 0.0387139529062546 |
| chr3:45829647:GAA:GA | 0.0280028534548443 |
| chr3:45866624:A:T | 0.0260363733864416 |
| chr3:45867022:C:G | 0.0253579710286574 |
| chr3:45867532:A:G | 0.0230662493702976 |

Supplementary Table 14: Results of Bayesian fine-mapping analysis using the *Sum of Single Effects (SuSiE)* R package [23] and summary statistics of the Italian GWAS dataset imputed with the *TOPMed Imputation Server*. A total of 25 variants were identified that belong to the 95% most likely to be causal. *chr3:45859597:C:T* respresents *rs73064425*. *chr3:45867022:C:G* respresents *rs35652899*. Columns are labeled *SNP (single nucleotide polymorphism)* and *PIP (posterior inclusion probability).*

| SNP | PIP |
|---|---|
| chr3:45867022:C:G | 0.0808376813637008 |
| chr3:45848429:A:T | 0.0628811810409609 |
| chr3:45838989:T:C | 0.0613824469908203 |
| chr3:45859597:C:T | 0.0584277827896861 |
| chr3:45848457:C:T | 0.0539823016468194 |
| chr3:45830416:G:A | 0.0530933287499147 |
| chr3:45847198:A:G | 0.0510644391263493 |
| chr3:45866624:A:T | 0.0502285274992595 |
| chr3:45823240:T:C | 0.0502034480773075 |
| chr3:45858159:A:G | 0.0493756613020777 |
| chr3:45834967:G:GA | 0.0482190641158137 |
| chr3:45821460:T:C | 0.0474942508530404 |
| chr3:45820440:G:A | 0.0471190140336258 |
| chr3:45825948:A:G | 0.0425074167185415 |
| chr3:45867532:A:G | 0.0424680345154714 |
| chr3:45818159:G:A | 0.0383782038280491 |
| chr3:45868152:C:A | 0.0265036967610202 |
| chr3:45859142:G:C | 0.0211914994676796 |
| chr3:45801947:G:T | 0.0131102672702105 |
| chr3:45868036:C:T | 0.0130732506830917 |
| chr3:45801823:C:T | 0.0109925898227481 |
| chr3:45807268:G:C | 0.00942629407775131 |
| chr3:45805277:A:G | 0.00763332534098549 |
| chr3:45801750:G:A | 0.00746659406054206 |
| chr3:45802706:A:C | 0.00730356963577561 |

[2] Taliun, D., Harris, D. N., Kessler, M. D., Carlson, J., Szpiech, Z. A., Torres, R., Taliun, S. A. G., Corvelo, A., Gogarten, S. M., Kang, H. M., Pitsillides, A. N., LeFaive, J., Lee, S.-B., Tian, X., Browning, B. L., Das, S., Emde, A.-K., Clarke, W. E., Loesch, D. P., Shetty, A. C., Blackwell, T. W., Smith, A. V., Wong, Q., Liu, X., Conomos, M. P., Bobo, D. M., Aguet, F., Albert, C., Alonso, A., Ardlie, K. G., Arking, D. E., Aslibekyan, S., Auer, P. L., Barnard, J., Barr, R. G., Barwick, L., Becker, L. C., Beer, R. L., Benjamin, E. J., Bielak, L. F., Blangero, J., Boehnke, M., Bowden, D. W., Brody, J. A., Burchard, E. G., Cade, B. E., Casella, J. F., Chalazan, B., Chasman, D. I., Chen, Y.-D. I., Cho, M. H., Choi, S. H., Chung, M. K., Clish, C. B., Correa, A., Curran, J. E., Custer, B., Darbar, D., Daya, M., de Andrade, M., DeMeo, D. L., Dutcher, S. K., Ellinor, P. T., Emery, L. S., Eng, C., Fatkin, D., Fingerlin, T., Forer, L., Fornage, M., Franceschini, N., Fuchsberger, C., Fullerton, S. M., Germer, S., Gladwin, M. T., Gottlieb, D. J., Guo, X., Hall, M. E., He, J., Heard-Costa, N. L., Heckbert, S. R., Irvin, M. R., Johnsen, J. M., Johnson, A. D., Kaplan, R., Kardia, S. L. R., Kelly, T., Kelly, S., Kenny, E. E., Kiel, D. P., Klemmer, R., Konkle, B. A., Kooperberg, C., Köttgen, A., Lange, L. A., Lasky-Su, J., Levy, D., Lin, X., Lin, K.-H., Liu, C., Loos, R. J. F., Garman, L., Gerszten, R., Lubitz, S. A., Lunetta, K. L., Mak, A. C. Y., Manichaikul, A., Manning, A. K., Mathias, R. A., McManus, D. D., McGarvey, S. T., Meigs, J. B., Meyers, D. A., Mikulla, J. L., Minear, M. A., Mitchell, B. D., Mohanty, S., Montasser, M. E., Montgomery, C., Morrison, A. C., Murabito, J. M., Natale, A., Natarajan, P., Nelson, S. C., North, K. E., O'Connell, J. R., Palmer, N. D., Pankratz, N., Peloso, G. M., Peyser, P. A., Pleiness, J., Post, W. S., Psaty, B. M., Rao, D. C., Redline, S., Reiner, A. P., Roden, D., Rotter, J. I., Ruczinski, I., Sarnowski, C., Schoenherr, S., Schwartz, D. A., Seo, J.-S., Seshadri, S., Sheehan, V. A., Sheu, W. H., Shoemaker, M. B., Smith, N. L., Smith, J. A., Sotoodehnia, N., Stilp, A. M., Tang, W., Taylor, K. D., Telen, M., Thornton, T. A., Tracy, R. P., Van Den Berg, D. J., Vasan, R. S., Viaud-Martinez, K. A., Vrieze, S., Weeks, D. E., Weir, B. S., Weiss, S. T., Weng, L.-C., Willer, C. J., Zhang, Y., Zhao, X., Arnett, D. K., Ashley-Koch, A. E., Barnes, K. C., Boerwinkle, E., Gabriel, S., Gibbs, R., Rice, K. M., Rich, S. S., Silverman, E. K., Qasba, P., Gan, W., NHLBI Trans-Omics for Precision Medicine (TOPMed) Consortium, Papanicolaou, G. J., Nickerson, D. A., Browning, S. R., Zody, M. C., Zöllner, S., Wilson, J. G., Cupples, L. A., Laurie, C. C., Jaquish, C. E., Hernandez, R. D., O'Connor, T. D., and Abecasis, G. R. (February, 2021) Sequencing of 53,831 diverse genomes from the NHLBI TOPMed Program. *Nature,* **590**(7845), 290–299.

[3] Loh, P.-R., Danecek, P., Palamara, P. F., Fuchsberger, C., Reshef, Y. A., Finucane, H. K., Schönherr, S., Forer, L., McCarthy, S., Abecasis, G. R., et al. (October, 2016) Reference-based phasing using the Haplotype Reference Consortium panel. *Nature Genetics,* **48**, 1443–1448.

[4] Wienbrandt, L. and Ellinghaus, D. (November, 2022) EagleImp: fast and accurate genome-wide phasing and imputation in a single tool. *Bioinformatics,* **38**, 4999–5006.

[5] Durbin, R. (2014) Efficient haplotype matching and storage using the positional Burrows-Wheeler transform (PBWT). *Bioinformatics,* **30**(9), 1266–1272.

[6] The 1000 Genomes Project Consortium (September, 2015) A global reference for human genetic variation. *Nature,* **526**, 68–74.

[7] Apache HTTP Server Project, https://httpd.apache.org.

[8] PostgreSQL: The World's Most Advanced Open Source Relational Database, https://www.postgresql.org.

[9] Jette, M. A. and Wickberg, T. (2023) Architecture of the Slurm Workload Manager. In Klusáček, D., Corbalán, J., and Rodrigo, G. P., (eds.), *Job Scheduling Strategies for Parallel Processing*, Cham: Springer Nature Switzerland pp. 3–23.

[10] A simple PHP WebAuthn (FIDO2) server library, https://github.com/lbuchs/WebAuthn.

[18] Byrska-Bishop, M., Evani, U. S., Zhao, X., Basile, A. O., Abel, H. J., Regier, A. A., Corvelo, A., Clarke, W. E., Musunuri, R., Nagulapalli, K., Fairley, S., Runnels, A., Winterkorn, L., Lowy, E., Eichler, E. E., Korbel, J. O., Lee, C., Marschall, T., Devine, S. E., Harvey, W. T., Zhou, W., Mills, R. E., Rausch, T., Kumar, S., Alkan, C., Hormozdiari, F., Chong, Z., Chen, Y., Yang, X., Lin, J., Gerstein, M. B., Kai, Y., Zhu, Q., Yilmaz, F., Xiao, C., Paul Flicek,

Germer, S., Brand, H., Hall, I. M., Talkowski, M. E., Narzisi, G., and Zody, M. C. (September, 2022) High-coverage whole-genome sequencing of the expanded 1000 Genomes Project cohort including 602 trios. *Cell,* **185**(18), 3426–3440.e19.
